# Genetic Associations with Age at Dementia Onset in the *PSEN1 E280A* Colombian Kindred

**DOI:** 10.1101/2020.09.23.20198424

**Authors:** J. Nicholas Cochran, Juliana Acosta-Uribe, Bianca T. Esposito, Lucia Madrigal, David Aguillón, Margarita M. Giraldo, Jared W. Taylor, Joseph Bradley, Brian Fulton-Howard, Shea J. Andrews, Natalia Acosta-Baena, Diana Alzate, Gloria P. Garcia, Francisco Piedrahita, Hugo E. Lopez, Ashlyn G. Anderson, Ivan Rodriguez-Nunez, Kevin Roberts, Dominantly Inherited Alzheimer Network, Devin Absher, Richard M. Myers, Gary W. Beecham, Christiane Reitz, Lindsay F. Rizzardi, Maria Victoria Fernandez, Alison M. Goate, Carlos Cruchaga, Alan E. Renton, Francisco Lopera, Kenneth S. Kosik

## Abstract

**INTRODUCTION:** Genetic associations with Alzheimer’s disease (AD) age at onset (AAO) could reveal genetic variants with therapeutic applications. We present a large Colombian kindred with autosomal dominant AD (ADAD) as a unique opportunity to discover AAO genetic associations.

**METHODS:** A genetic association study was conducted for ADAD dementia AAO in 340 individuals with the *PSEN1 E280A* mutation via TOPMed array imputation. Replication was assessed in two ADAD cohorts, one sporadic EOAD study, and four late onset AD studies.

**RESULTS:** 13 variants had *p*<1×10^−7^ or *p*<1×10^−5^ with replication including three independent loci with candidate associations with clusterin including near *CLU*. Other suggestive associations were identified in or near *HS3ST1, HSPG2, ACE, LRP1B, TSPAN10*, and *TSPAN14*.

**DISCUSSION:** Variants with suggestive associations with AAO were associated with biological processes including clusterin, heparin sulfate and amyloid processing. The detection of these effects in the presence of a strong mutation for ADAD reinforce their potentially impactful role.

## Background

Complex genetic, environmental, and lifestyle risk factors confounded by the aging process underlie risk for late onset Alzheimer’s disease (LOAD). Autosomal dominant Alzheimer’s disease (ADAD) closely resembles the clinical and neuropathological features of LOAD, but without the confound of aging, and thus provides a less heterogeneous view of underlying AD-associated processes. ADAD accounts for less than 1% of all cases of AD and mutations in *PSEN1* account for 80% of this monogenic group (reviewed in [1]).

There is a strong correlation between age at onset (AAO) and a particular ADAD mutation (r^2^ = 0.52) [2], but there still remains substantial unexplained variability. Large ADAD families such as the kindred harboring the Colombian *PSEN1* NM_000021:c.839A>C, p.(Glu280Ala) (canonically known as *PSEN1 E280A*) mutation, the world’s largest ADAD founder population with a comprehensive family tree of thousands of individuals [3], provide an opportunity to assess the contribution of genetic variation to unexplained variability in age of dementia onset. *PSEN1 E280A* mutation carriers typically develop mild cognitive impairment (MCI) at a median age of 44 years (95% CI, 43–45) and dementia at age of 49 years (95% CI, 49– 50) [4]. The value of this family for the nomination of genetic variants that delay the onset of AD was recently affirmed by the report of a *PSEN1 E280A* carrier who developed MCI nearly three decades after the kindred’s median age at clinical onset [5] (this individual is also included in this study). This individual was homozygous for the rare *APOE* ε3 Christchurch variant (*APOE* NM_000041:c.460C>A, p.(R154S), rs121918393) and had an exceptionally high amyloid-β plaque burden, but limited neurofibrillary tau burden. In addition to this case report, several studies have explored genetic associations with AAO in *PSEN1 E280A* carriers [6-9], but all with substantially lower numbers of cases (at most 72 individuals) [6]. To expand on the valuable insights gained from these previous studies, we conducted the most comprehensive search to date for genetic variants associated with age at dementia onset in this founder population by assessing 340 individuals, which is the current snapshot of all individuals from this cohort, that currently have high quality genotypic and phenotypic information available.

## Methods

### Patient Recruitment

A cohort of 368 patients was selected from the Neuroscience Group of Antioquia (GNA) database of the *PSEN1 E280A* family. After all quality control steps, 340 individuals remained for analysis. Selection criteria included being a *PSEN1 E280A* carrier with diagnosis of dementia, having adequate medical and neuropsychological evaluations and follow-up for a confident age determination of clinical age at dementia onset, and having a DNA sample. Participants were evaluated following a standard protocol including physical and neurological examination, as well as population-validated neuropsychological assessment [10, 11]. Dementia was diagnosed according to most recent DSM criteria at the time of diagnosis. Collected data were stored in medical records software (SISNE v2.0). Family history was obtained from the patients and their relatives, and genealogical data from baptism and death certificates was gathered from local parishes and was incorporated into the pedigree reconstruction. Blood samples from each individual were obtained through standard phlebotomy and collected in EDTA tubes. Genomic DNA was purified from peripheral blood leukocytes using a modified salting-out technique (Gentra Puregene Blood Kit, Qiagen). All individuals were genotyped for *PSEN1 E280A* using a restriction length fragment polymorphism assay.

### Genotyping Arrays

1,923,394 variants were genotyped using the Illumina Multi-Ethnic Genotyping Array plus Neuro consortium content (catalog #WG-316-1014, beadchip #20028352). Data were annotated with build hg38 and analyzed using PLINK v1.90b5.2, PLINK v2.00aLM [12], and GEMMA [13]. Genetic relatedness was assessed using KING 2.2 [14].

### Genome Sequencing and Annotation

Genome sequencing and preparatory processing of genome data prior to analysis was as previously described [15]. Genome sequencing was conducted for a subset (80) of the individuals in the cohort and was used in this study for quality control only. For intersection of array data, which was annotated with hg38, we lifted over genome sequencing data using CrossMap 0.3.4 [16].

### Variant Measurement, Imputation, and Quality Control

Imputation was conducted using the TOPMed Imputation Panel and Server (version 1.3.3), which includes 97,256 references samples and 308,107,085 variants and uses Minimac4 for imputation. Pre-imputation scripts (version 4.3.0 from William Rayner at the University of Oxford) [17] were run using default settings, which filtered out palindromic single nucleotide variants (SNVs) with minor allele frequency (MAF) > 0.4 or variants with > 0.2 MAF difference from the TOPMed reference panel. For samples with both array and genome sequencing data, imputed genotypes were checked for concordance with genome sequencing data using SnpEff 4.3s.

373 arrays were measured over two batches (299 in batch one and 74 in batch two) representing 368 presumed unique individuals. The five duplicated individuals were from the following: four individuals from batch one with high missingness were re-run, and a new aliquot of one sample from batch one that was identified as a duplicate of another sample was re-run. Of the 373 array measurements, three were excluded because they were identified as mislabeled duplicates of other samples, and 33 were excluded because they had a missingness rate of greater than 5%, heterozygosity >3 standard deviations from the mean, were <95% concordant with genome sequencing data, or lacked age at dementia onset data. 340 unique individuals with high quality array data remained for analysis.

26 genomes were sequenced at HudsonAlpha including seven out of eight of the oldest age at onset individuals along with their affected family members with more typical ages of onset. 55 genomes from additional E280A carriers were sequenced previously with CompleteGenomics technology [6].

We used both array and genome information to generate a high-quality variant set for use in imputation. Array data were further selected (beyond the already mentioned cutoff of a maximum 5% missingness for any sample) to a maximum missingness of 1% for any genotype. A Hardy-Weinberg equilibrium cutoff of 1×10^−6^was used. Variants were quality checked against TOPMed Freeze 5 PASS filter variants using pre-imputation scripts as described above. The pre-imputation quality control pipeline is available at https://github.com/HudsonAlpha/Pre-Imputation-QC-Pipeline and post-imputation at https://github.com/HudsonAlpha/Post-Imputation-Pipeline. Variants with Mendelian inconsistencies were excluded, resulting in 844,970 array-measured variants after QC. For genome sequencing data, we selected PASS filter variants in the HudsonAlpha-sequenced set that also had a 99% call rate across both HudsonAlpha and CompleteGenomics genomes. The intersection of variants from genome sequencing and array measurements was then used to assess concordance between samples with both types of measurements, and one sample was excluded for <95% concordance between measurement types (median concordance for the remaining 80 samples was 99.94%, indicating the high quality of this intersected variant set). 340 total unique individuals with high quality data remained for imputation, and 540,753 variants met all filtering criteria for both arrays and genome sequencing.

The set of 540,753 high quality variants for 340 unique E280A carriers was used as input for imputation using the TOPMed imputation server (version 1.3.3), with an output of 20,707,761 variants passing an r^2^ cutoff of 0.3. Because of the importance of the *APOE* locus, we inspected the imputation results for rs429358 and rs7412, which together define *APOE* ε alleles. All genotypes matched between imputed and separately genotyped values for rs7412, and all but one genotype matched between imputed and separately genotyped values for rs429358. For the discordant genotype, the directly measured genotype was used. In addition, although it was not imputed because of rarity, we manually inserted directly measured genotypes for rs121918393 because of its implicated role in a previous study [5]. As an additional quality control measure, we filtered this imputation set to variants called in HudsonAlpha-sequenced genomes with a VQSLOD score > -3 and again filtered any variants with Mendelian inconsistencies, resulting in a final set of 9,430,010 high quality variants. We considered all variants as a part of the relatedness matrix to obtain the best adjustment for relatedness possible, but only considered variants with an allele count of at least 3 for association testing, yielding 9,012,264 variants.

### Replication sets

Seven cohorts were selected for replication. For ADAD, we used the Dominantly Inherited Alzheimer’s Network (DIAN) cohort, with 116 cases with age of dementia onset as the phenotype as in the main cohort analyzed. The DIAN cohort was analyzed using GEMMA on TOPMed data with an allele frequency cutoff of 1% for all variants considered. Covariates included the parental age at onset, the gene, including considering *PSEN1* before and after codon 200 as separate “genes” given more deleterious effects of *PSEN1* variants after codon 200 [18], and the first three principal components.

As a second dominant AD replication cohort, we used the Alzheimer’s Disease in Adults with Down Syndrome (ADDS) cohort, which was obtained from the Synapse AD Knowledge portal (Synapse ID: syn25871263) and imputed using TOPMed. After quality control for missingness, heterozygosity, and relatedness, 222 individuals remained for analysis. We used the available phenotype if the individuals had converted to MCI or AD (105 not yet converted, 58 MCI, and 59 AD) weighted as 0, 0.5, and 1 respectively for the phenotype. We performed the GEMMA analysis in the same manner as our cohort, with sex and PCs 1-10 included in the model. For this cohort, chromosome 21 was not considered for replication.

Given the limited sample sizes for dominant AD, we also evaluated sporadic AD cohorts. For EOAD, we evaluated the largest sporadic early onset AD cohort aggregated to date, an ADGC EOAD study cohort currently in analysis with 6,282 European ancestry early onset AD cases and 13,386 European ancestry controls (European ancestry is the largest admixture component in our cohort). For this cohort, all Single variant analyses were performed with Plink v2.0 GLM function with the following model: Status∼SNP+SEX+PC1-10. For LOAD, we selected an AD age at onset study (9,162 cases) [19], a study of AD age at onset survival (14,406 cases and 25,849 controls) [20], a genome wide association study (GWAS) meta-analysis for AD (21,982 AD vs. 41,944 controls) [21], and the latest meta-analysis of AD and AD by proxy (111,326 cases and 677,663 controls) [22]. See supplemental methods for discussion of International Genomics of Alzheimer’s Project (IGAP) replication data.

## Results

### Cohort demographics

The final cohort had a mean age of dementia onset of 49.3 years (median: 48, range: 37–75, 10^th^–90^th^ percentile: 43–56). 198 of the patients were genetically female (58.2%). The patients had extensive follow up data; the mean number of medical evaluations was 6.7 (1–27), and 4.8 (1–18) for neuropsychological evaluations. A partial pedigree of enrolled individuals annotated with age at dementia onset is presented in **Supplemental Figure 1**.

### Association analysis

Association analysis was conducted using age at dementia onset as a quantitative outcome for 340 individuals passing QC. We employed GEMMA, a package that performs a likelihood ratio test using a linear mixed model to adjust for relatedness between individuals. For both packages, we adjusted for genetic sex, the first ten principal components (calculated from the set of 540,753 high quality variants used as imputation input using PLINK v2.00aLM) because this was an admixed population, and batch. The chip heritability calculated by GEMMA was 0.74+/-0.14 with a Vg estimate of of 24.6 and Ve estimate of 8.5.

### Top nominally significant loci of interest

To determine if any hits observed statistically deviated from random chance, we generated a QQ plot (**Supplemental Figure 2**). No variants deviated detectably from a uniform distribution’s expected error range except for from modest inflation (genomic inflation factor of 1.05), but this was not surprising given the small size of the cohort, and the modest level of inflation reflects that GEMMA’s kinship matrix adjustment works well for this familial cohort. Because of this, the variants presented throughout should be viewed as speculative. To add evidence for possible biological significance, we relied primarily on replication. First, we compared the number of variants with *p*<1×10^−5^ that exhibit nominal replication (p<0.05) in one of the seven replication cohorts. Second, we used a stricter threshold (*p*<1×10^−7^) where we did not require replication. The result of these filtering conditions is shown in **Table 1** and includes three variants at different loci associated with clusterin biology, rs138295139, rs35980966 and rs4942482. LocusZoom plots and single nucleus multiomics linkages (correlations between single nucleus RNA-seq and ATAC-seq from the same nuclei [23]) for these variants are presented in **Figure 1**. In addition, the age of each individual harboring each variant in **Table 1** is illustrated in **Supplemental Figure 3** by variant to illustrate the spread of the variants across the cohort by zygosity. The Manhattan plot for the study is shown in **Supplemental Figure 4**. Unannotated summary statistics for all variants are provided in **Supplemental Table 1**. Annotated summary statistics including replication information from described cohorts for variants with *p*<1×10^−5^, coding variants with *p*<0.05, APOE coding variants, and variants that are index variants for previous GWAS are provided in **Supplemental Table 2**. Variants with *p*<1×10^−5^ that overlap with a single nucleus multiomics linkage between ATAC-seq and RNA-seq in the same nuclei from a recent study [23] are shown in **Supplemental Table 3** along with more detailed information. LocusZoom plots of all regions with *p*<1×10^−5^ are presented in **Supplemental File 1**.

**Table 1:**
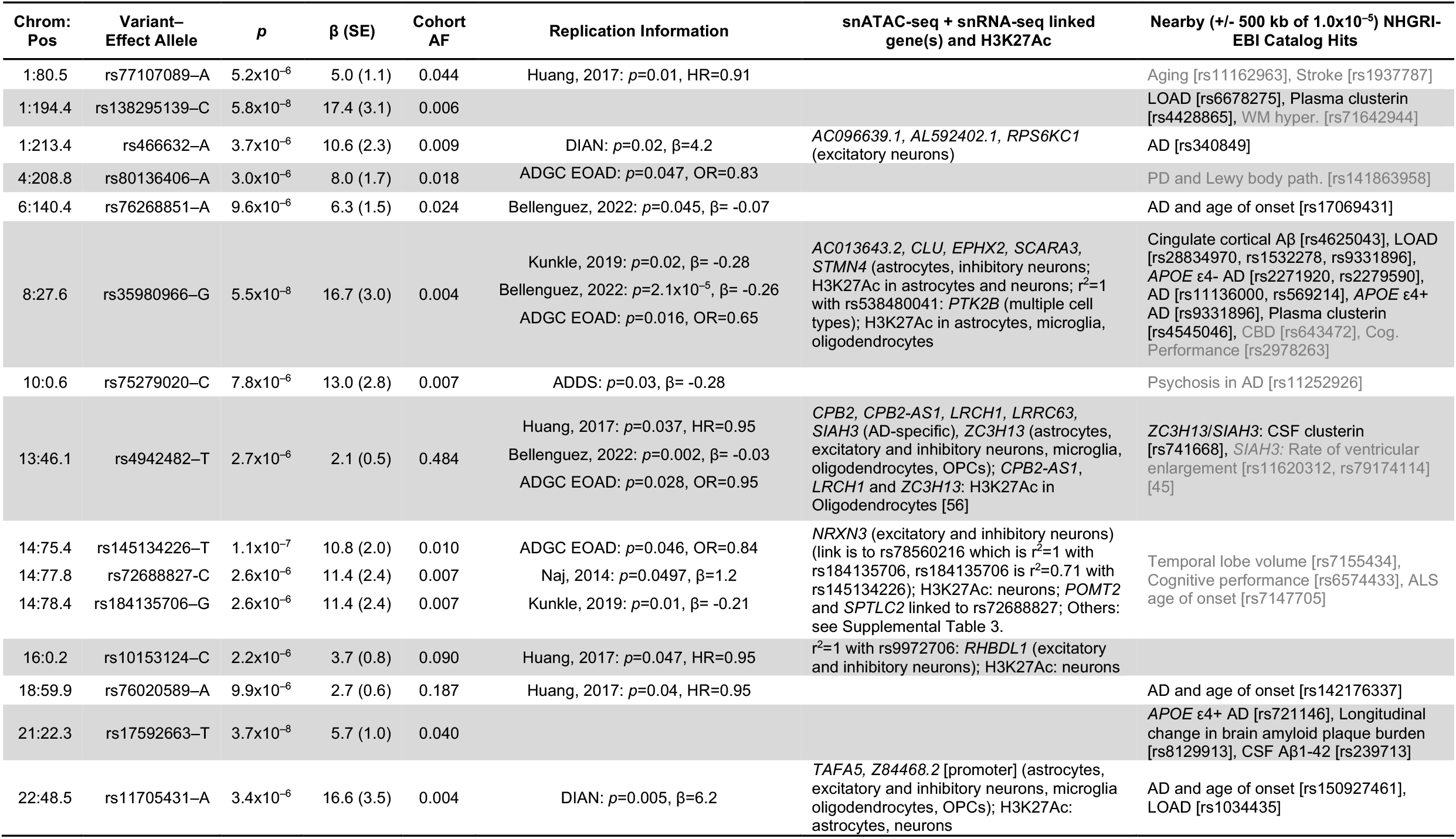
Variants that both met a suggestive threshold (*p*<1×10^−5^) and exhibited one or both of replication at *p*=0.05 and consistent effect direction in at least one cohort and/or presence in a locus within 500 kb of a GWAS variant previously linked to an AD-associated phenotype (less closely related phenotypes are shown in gray). β is the effect in years on age at dementia onset (i.e., positive β values indicate an association with later age of onset). Replication cohorts varied in design but required a consistent effect direction with the matched effect allele. Naj, 2014 is an age of onset study where a positive β indicates later age of onset. Huang, 2017 is an age of onset survival design where a hazard ratio less than 1 indicates a protective correlation. The DIAN cohort was analyzed in the same manner as the main cohort. Bellenguez, 2022, Kunkle, 2019, and ADDS are case/control designs where negative betas indicate a protective correlation. Full summary statistics are presented in Supplemental Table 1 and All variants with *p*<1×10^−5^ along with more detailed information are presented in Supplemental Table 2. **Abbreviations:** Chrom:Pos – Chromosome: build hg38 position, Cohort AF – Cohort allele frequency, DIAN – Dominantly Inherited Alzheimer’s Network cohort, ADDS – Alzheimer’s Disease in Adults with Down Syndrome cohort, LOAD – late onset Alzheimer’s disease, AD – Alzheimer’s disease, Aβ – amyloid beta.

**Figure 1:**
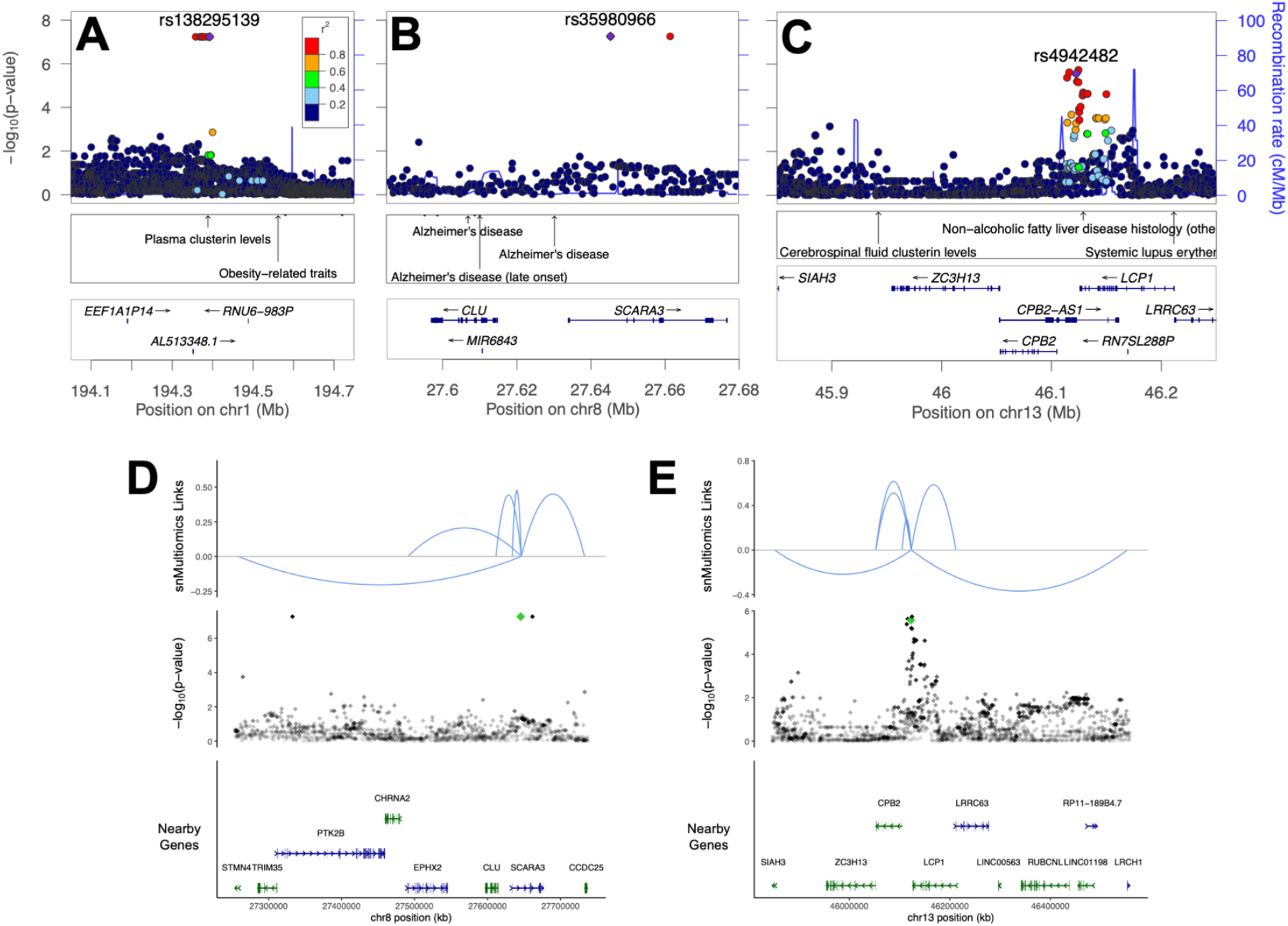
Plots of loci linked to *clusterin* or clusterin related phenotypes meeting criteria in Table 1. (**A–C**) LocusZoom plots of loci with index variants indicated with a purple diamond. Nearby NHGRI-EBI GWAS hits are indicated. (**A**) Note nearby variant previously linked to plasma clusterin levels. (**B**) Note several nearby variants previously linked to AD near *CLU*. (**C**) Note nearby variant previously linked to CSF clusterin levels between *SIAH3* and *ZC3H13*. (**D**,**E**) Single nucleus multiomic (snMultiomics) links (RNA-seq–ATAC-seq correlations from the same nuclei) indicated for hits on chromosomes 8 and 13. Strength of the link is indicated by height, and direction indicates direction of correlation. Index variants are indicated with a green diamond. (**D**) Note link to *CLU*. (**E**) Note links to *SIAH3* and *ZC3H13*.

### Results at key APOE variants

Effects of previously established *APOE* variants important for AD association in LOAD are in the expected direction based on previous studies, but modest in magnitude (**Table 2**). Overall, the observations are consistent with previously reported observations including a protective effect of *APOE* ε2 in the Colombian E280A population (β=8.2, 95% CI=4.5–12.0, *p*=3.8×10^−5^) [24], a deleterious effect of *APOE* ε4 in the Colombian E280A population in one study (hazard ratio 2.1, 95% CI 1.1–4.0, *p*=0.03) [25] but an inability to detect an effect of *APOE* ε4 in three other studies in this population [24, 26, 27], and a non-significant trend towards an *APOE* ε2 > *APOE* ε3 > *APOE* ε4 age-of-onset in dominant AD families with a variety of mutations [2].

**Table 2:** Assessment of previously implicated variants in *APOE*. These variants were directly genotyped by qPCR given their previously implicated role in disease. β is the effect in years on age at dementia onset (i.e., positive β values indicate an association with later age of onset). One other *APOE* coding variant, rs440446–G, was imputed by TOPMed but did not exhibit a meaningful signal (β= -0.57, *p*=0.16, see Supplemental Table 2 for details). TOPMed Bravo Freeze 8 was used for Population Allele Frequency.

| Variant–<br>Effect Allele | Description | $\beta$ (SE) | $p$ | #<br>Homozygous | Cohort<br>Allele<br>Frequency | Population<br>Allele<br>Frequency | Transcript<br>ENST00000252486.9<br>Coding Change | CADD<br>version<br>1.6 |
| --- | --- | --- | --- | --- | --- | --- | --- | --- |
| rs429358–C | *APOE* $\epsilon 4$ (when rs7412 ref.) | -2.0 (0.6) | 0.0025 | 8 | 0.138 | 0.155 | p.(Cys130Arg) | 16.7 |
| rs121918393–A | *APOE* Christchurch | 3.1 (1.6) | 0.0496 | 1 | 0.020 | $1.9 \times 10^{-5}$ | p.(Arg154Ser) | 25.3 |
| rs7412–T | *APOE* $\epsilon 2$ (when rs429358 ref.) | 2.0 (0.9) | 0.0891 | 0 | 0.068 | 0.078 | p.(Arg176Cys) | 26 |

A recent case report implicated the *APOE* Christchurch variant (rs121918393) [5]. That individual was also enrolled in this study, and while we do observe a nominally significant effect on age at onset of this variant, we note that the effect size is modest, which could be because our model does not consider homozygosity effects. No other coding variants in *APOE* beyond those described in **Table 2** were observed in either the imputed set or the subset of cases with genomes available.

### Replication at known AD-associated loci

We evaluated 17 AD GWAS, including the largest case/control studies for AD in European populations [21, 22, 28-30], studies in non-Europeans [31-35], age at onset modifier studies [19, 20], and endophenotype studies [36-40]. These studies identified 108 loci (at least 500kb between unique loci) and 184 index variants within these loci with high confidence associations for AD and endophenotypes (**Supplemental Table 4** (an expansion from a table put forward by [41])). Of these variants, 151 were genotyped in this cohort, nine with *p*<0.05, but only six of these were in a consistent direction. Replication of hits with genome-wide significance for AD-associated phenotypes with nominal significance (*p*<0.05) with consistent effect direction in this cohort are shown in **Table 3**. This table should be interpreted with caution, as it is close to the number of variants that would be expected based on random sampling of this set of GWAS hits (six observed versus ∼four expected), however the variants identified do share some nearby genes or pathways with variants from other nomination approaches (see Discussion).

**Table 3:** Hits with genome-wide significance for AD-associated phenotypes from previous studies with *p*<0.05 and consistent direction of effect in this cohort. β is the effect in years on age at dementia onset (i.e., positive β values indicate an association with later age of onset). For previous studies, all effects are meta-analysis OR (95% CI) except Beecham et al., 2014 and Deming et al., 2017, which are β (SE). **Abbreviations:** Cohort AF indicates minor allele frequency in this cohort. Population AF indicates the population allele frequency (TOPMed Bravo Freeze 8). CADD – Combined Annotation Dependent Depletion score.

| Variant | Locus | Key Nearby Gene(s) | $p$ | $\beta$ (SE) | Cohort AF | Population AF | Previous Study | Previous Study $p$ | Previous Study Effect (Error) | Previous Study Outcome | Other Replication |
| --- | --- | --- | --- | --- | --- | --- | --- | --- | --- | --- | --- |
| rs6448453-G | 4p16.1 | *CLNK/HS3ST1* | 0.010 | 1.4 (0.5) | 0.75 | 0.77 | Jansen, 2019 | $1.9 \times 10^{-09}$ | 0.99 (0.98–0.99) | AD Risk | Huang, 2017<br>Kunkle, 2019<br>Bellenguez, 2022 |
| rs62341097-A | 4q34.1 | *GALNT7* | 0.035 | 2.2 (1.2) | 0.05 | 0.03 | Beecham, 2014 | $6.0 \times 10^{-09}$ | -1.147 (0.198) | Neuritic Plaque | |
| rs316341-A | 6p25 | *SERPINB1* | 0.015 | -1.3 (0.5) | 0.69 | 0.71 | Deming, 2017 | $1.8 \times 10^{-08}$ | -0.025 (0.004) | CSF Aβ42 | |
| rs6586028-T | 10q23.1 | *TSPAN14* | 0.035 | -1.4 (0.7) | 0.86 | 0.87 | Bellenguez, 2022 | $2.0 \times 10^{-19}$ | 1.08 (1.06–1.10) | AD Risk | Huang, 2017<br>Kunkle, 2019<br>DIAN |
| rs1140239-T | 16p11.2 | *DOC2A* | 0.026 | 1.0 (0.5) | 0.39 | 0.31 | Bellenguez, 2022 | $2.6 \times 10^{-13}$ | 0.94 (0.93–0.96) | AD Risk | Huang, 2017<br>Kunkle, 2019 |
| rs138190086-A | 17q23.3 | *ACE* | 0.021 | -3.0 (1.3) | 0.03 | 0.01 | Marioni, 2018<br>Kunkle, 2019 | $1.9 \times 10^{-09}$<br>$5.3 \times 10^{-09}$ | 1.25 (1.16–1.35)<br>1.30 (1.19–1.42) | AD Risk<br>AD Risk | Bellenguez, 2022 |

In addition to testing known LOAD risk loci individually, we also evaluated the effect of LOAD variants combined using a LOAD polygenic risk score (**Figure 2**). Polygenic risk score both without (**Figure 2A**) and with *APOE* ε allele–defining variants rs429358 and rs7412 (**Figure 2B**) exhibited a significant correlation with age at dementia onset in the expected direction (later age of onset associated with a lower polygenic risk score).

**Figure 2:**
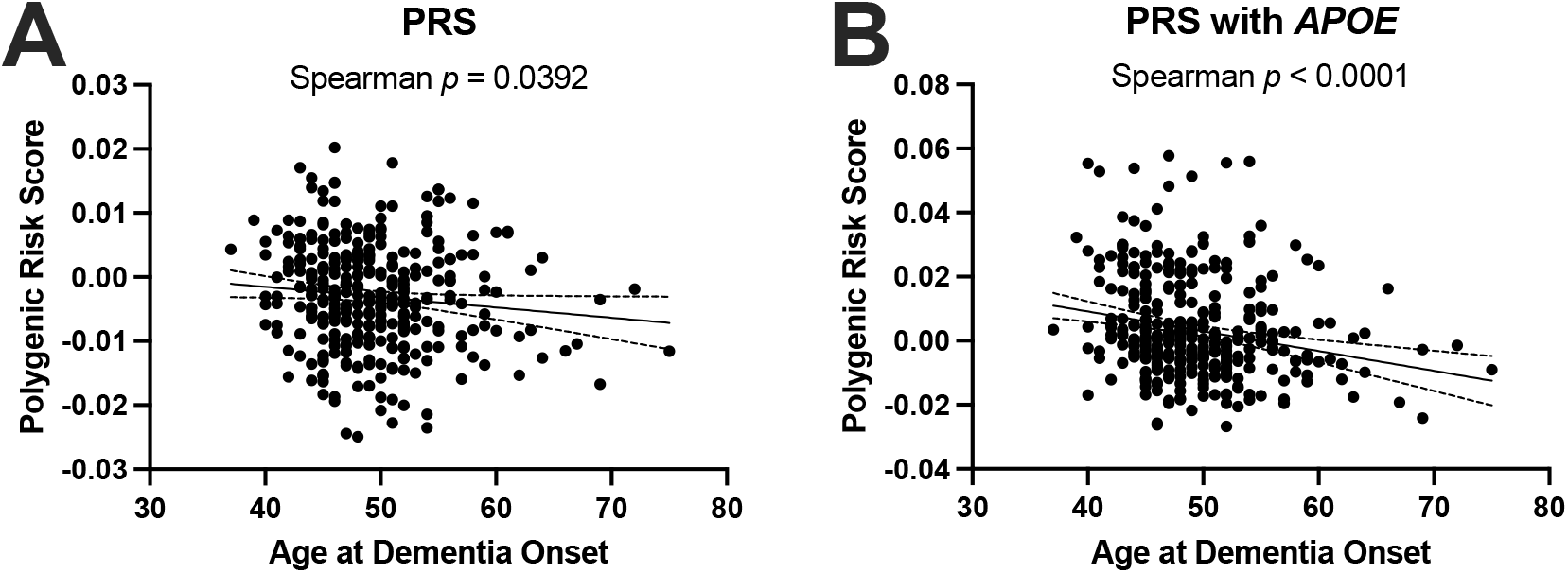
Late onset AD polygenic risk score applied to the *PSEN1 E280A* cohort. **(A)** LOAD polygenic risk score with APOE excluded (Spearman *p* = 0.0392). **(B)** LOAD polygenic risk score with APOE included (Spearman *p* < 0.00001).

### Coding variants of interest

We next asked if any coding variants speculatively associate with age of dementia onset (**Table 4**). We chose four conditions: *p*<1×10^−5^; *p*<0.01, Combined Annotation Dependent Depletion (CADD) phred score [42] >20 and replication in more than 1 study; *p*<0.01, population allele frequency < 2%, CADD>20, and replication in at least 1 study; and coding variants in high priority AD genes with *p*<0.05 including *APP, PSEN1, PSEN2, MAPT, APOE* (not shown because in **Table 2**), *ABCA7, SORL1, TREM2*, and recently implicated GWAS loci with signal for coding variation in a recent exome meta-analysis [43] including *ATP8B4, ABCA1, ADAM10, CLU, ZCWPW1*, and *ACE*.

**Table 4:**
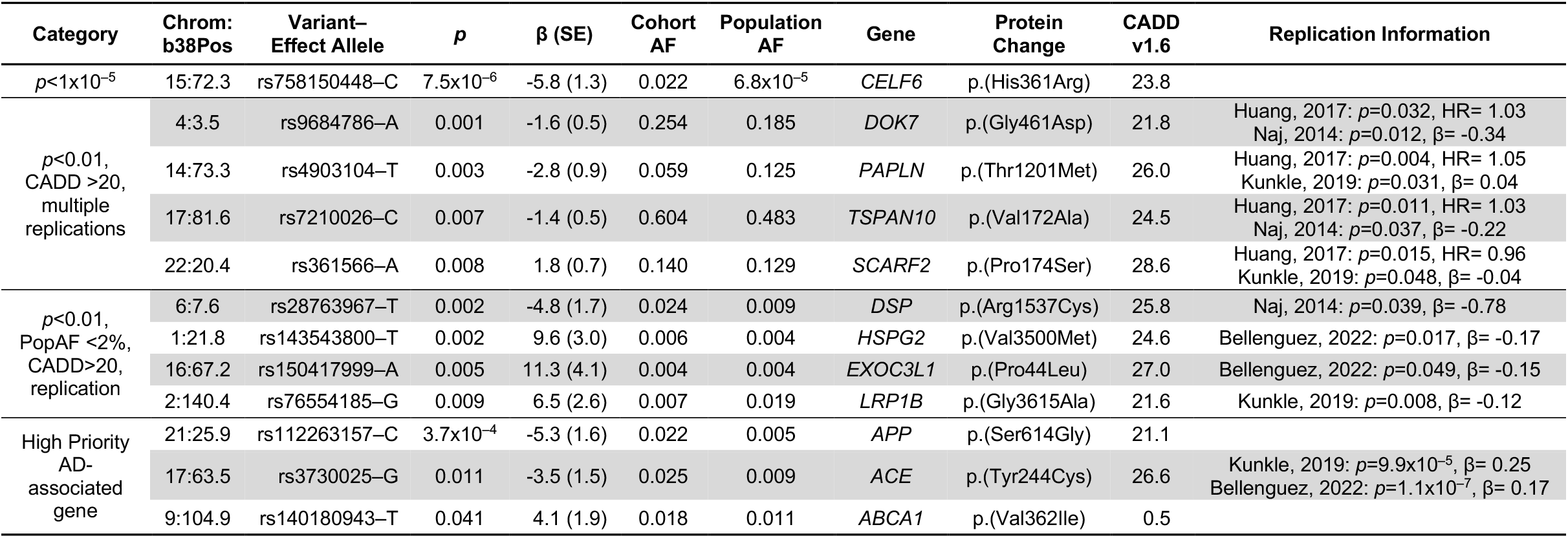
Coding variants of speculative interest. Conditions used to filter to variant categories highlighted in this table are noted. β is the effect in years on age at dementia onset (i.e., positive β values indicate an association with later age of onset). Replication cohorts varied in design but required a consistent effect direction with the matched effect allele. Huang, 2017 is an age of onset survival design where a hazard ratio less than 1 indicates a protective correlation. Naj, 2014 is an age of onset design with the same directional effect as this cohort. Kunkle, 2019 and Bellenguez, 2022 are case/control designs where negative betas indicate a protective correlation. All coding variants with *p*<0.05 along with more detailed information (including transcript for coding changes) are presented in Supplemental Table 2. **Abbreviations:** Chrom: b38 Pos – Chromosome: build hg38 position, Cohort AF – Cohort allele frequency, Population AF indicates the population allele frequency (TOPMed Bravo Freeze 8), CADD v1.6 – Combined Annotation Dependent Depletion score version 1.6.

| Category | Chrom: b38Pos | Variant–Effect Allele | $p$ | $\beta$ (SE) | Cohort AF | Population AF | Gene | Protein Change | CADD v1.6 | Replication Information |
| --- | --- | --- | --- | --- | --- | --- | --- | --- | --- | --- |
| $p < 1 \times 10^{-5}$ | 15:72.3 | rs758150448–C | $7.5 \times 10^{-6}$ | -5.8 (1.3) | 0.022 | $6.8 \times 10^{-5}$ | *CELF6* | p.(His361Arg) | 23.8 | |
| $p < 0.01$,<br>CADD >20,<br>multiple<br>replications | 4:3.5 | rs9684786–A | 0.001 | -1.6 (0.5) | 0.254 | 0.185 | *DOK7* | p.(Gly461Asp) | 21.8 | Huang, 2017: $p=0.032$, HR= 1.03<br>Naj, 2014: $p=0.012$, $\beta= -0.34$ |
| | 14:73.3 | rs4903104–T | 0.003 | -2.8 (0.9) | 0.059 | 0.125 | *PAPLN* | p.(Thr1201Met) | 26.0 | Huang, 2017: $p=0.004$, HR= 1.05<br>Kunkle, 2019: $p=0.031$, $\beta= 0.04$ |
| | 17:81.6 | rs7210026–C | 0.007 | -1.4 (0.5) | 0.604 | 0.483 | *TSPAN10* | p.(Val172Ala) | 24.5 | Huang, 2017: $p=0.011$, HR= 1.03<br>Naj, 2014: $p=0.037$, $\beta= -0.22$ |
| | 22:20.4 | rs361566–A | 0.008 | 1.8 (0.7) | 0.140 | 0.129 | *SCARF2* | p.(Pro174Ser) | 28.6 | Huang, 2017: $p=0.015$, HR= 0.96<br>Kunkle, 2019: $p=0.048$, $\beta= -0.04$ |
| $p < 0.01$,<br>PopAF <2%,<br>CADD>20,<br>replication | 6:7.6 | rs28763967–T | 0.002 | -4.8 (1.7) | 0.024 | 0.009 | *DSP* | p.(Arg1537Cys) | 25.8 | Naj, 2014: $p=0.039$, $\beta= -0.78$ |
| | 1:21.8 | rs143543800–T | 0.002 | 9.6 (3.0) | 0.006 | 0.004 | *HSPG2* | p.(Val3500Met) | 24.6 | Bellenguez, 2022: $p=0.017$, $\beta= -0.17$ |
| | 16:67.2 | rs150417999–A | 0.005 | 11.3 (4.1) | 0.004 | 0.004 | *EXOC3L1* | p.(Pro44Leu) | 27.0 | Bellenguez, 2022: $p=0.049$, $\beta= -0.15$ |
| | 2:140.4 | rs76554185–G | 0.009 | 6.5 (2.6) | 0.007 | 0.019 | *LRP1B* | p.(Gly3615Ala) | 21.6 | Kunkle, 2019: $p=0.008$, $\beta= -0.12$ |
| High Priority<br>AD-associated<br>gene | 21:25.9 | rs112263157–C | $3.7 \times 10^{-4}$ | -5.3 (1.6) | 0.022 | 0.005 | *APP* | p.(Ser614Gly) | 21.1 | |
| | 17:63.5 | rs3730025–G | 0.011 | -3.5 (1.5) | 0.025 | 0.009 | *ACE* | p.(Tyr244Cys) | 26.6 | Kunkle, 2019: $p=9.9 \times 10^{-5}$, $\beta= 0.25$<br>Bellenguez, 2022: $p=1.1 \times 10^{-7}$, $\beta= 0.17$ |
|  | 9:104.9 | rs140180943–T | 0.041 | 4.1 (1.9) | 0.018 | 0.011 | *ABCA1* | p.(Val362Ile) | 0.5 |  |

### Shared pathways between previous GWAS and coding variants of interest

Several pathways emerged with variants in both the previous GWAS replication set and the coding variants of interest set. First, *TSPAN14* and *TSPAN10* are involved in scaffolding ADAM10 and had GWAS and coding variants respectively. Second, *ACE* had a GWAS and coding variant. Third, *HS3ST1* had a GWAS variant, and *HSPG2* had a coding variant, with both involved in heparin sulfate biology.

## Discussion

Genetic association studies for LOAD are limited by heterogeneity of cases and unknown levels of contribution from environmental sources. This study addresses these limitations by employing a well-described phenotype in a geographically isolated population with a monogenic form of AD [3]. While environmental influences will always be present, this population has a relatively homogeneous set of environmental influences.

We identified 13 loci with *p*<1×10^−5^ and replication or *p*<1×10^−7^ with a nearby GWAS hit associated with AD phenotypes as well as more speculative signals when considering replication of previous GWAS in this cohort or important coding variants. This study nominates several important biological processes and pathways for consideration including clusterin, heparin sulfate and amyloid processing.

One of the most significant variants was rs35980966 (p=5.5×10^−8^), which is a rare variant (gnomAD v3.1.2 MAF=0.35%) that tags the *CLU* locus on chromosome 8 and exhibits replication in three studies [21, 22] and ADGC EOAD study in progress. The variant falls within a single nucleus multiomics linkage [23] to *CLU*. In addition, rs138295139 on chromosome 1 is only 4.4kb from a variant previously associated with plasma clusterin, rs4428865 [44], though these variants are not in LD, which could be explained by the rarity of rs138295139. Finally, rs4942482 on chromosome 13 is near a variant previously associated with CSF clusterin [44] and replicates in three studies [20, 22] and ADGC EOAD study in progress. This variant is linked via single nucleus multiomics measurements to nearby genes including *ZC3H13* and *SIAH3* (the linkage to *SIAH3* is particularly interesting as it is AD-specific). The variant previously associated with CSF clusterin levels [44] falls between these genes. In addition, *SIAH3* has been associated through another GWAS to rate of ventricular enlargement in the ADNI cohort [45], an association that has also been separately observed with variants near *CLU* [46]. Taken together, these observations, along with evidence for diverse contributions of clusterin in LOAD (recently review in [47]), suggest that further investigation of the role of clusterin and processes that may influence the effects of clusterin in ADAD is warranted.

Two variants were identified in or near heparin sulfate associated genes including rs6448453, a common variant near *HS3ST1*, and rs143543800, a rare variant in *HSPG2*. Heparin sulfate has been implicated in cell-to-cell spread of tau [48] as well as other AD-associated processes [49], pointing to potential importance of this pathway for dominant AD.

Variants in genes associated with amyloid processing were also identified in this study. A common variant in *TSPAN14*, rs6586028 (recently newly implicated in LOAD [22]), replicated in this cohort, and we also identified a coding variant in *TSPAN10* (**Table 4**). These two genes code for tetraspanins that are a part of the TspanC8 subgroup of tetraspanins which promote ADAM10 maturation [50]. Given ADAM10’s established role as an α-secretase promoting non-amyloidogenic processing of Aβ [51] as well as its ability to cleave TREM2 (reviewed in [52]) and the recent association of genetic variation in or near *ADAM10* with AD risk by GWAS [21, 28, 30] along with a candidate study of mutations [53], the basis of the observed association between age of dementia onset and these variants in *TSPAN14* and *TSPAN10* (both with a deleterious correlation) may result from disruption of a protective role of ADAM10.

As the largest age at onset modifier study in ADAD to date (to our knowledge), this study has nominated several new candidate genetic associations with age of dementia onset in ADAD. The most important limitation of this study is the small sample size (despite being the largest available sample size for this population) which precluded variants passing multiple corrections adjusted genome-wide significance. However, because we analyzed the three largest ADAD datasets available in the field, it is not possible to further increase sample size or replication in ADAD, and we therefore present these findings in light of replication with these available ADAD cohorts as well as sporadic EOAD and LOAD cohorts. Still, we recognize the speculative nature of the nominal associations identified in this study. Recruitment of more patients with early onset and/or dominant dementias from South American countries will help to overcome this limitation in future studies [54].

An important overarching theme from this analysis is that while age at dementia onset in ADAD has a strong heritable component, it is likely that, as with LOAD, there are many different genetic contributors that sum to determine an individual’s age at dementia onset for ADAD. Based on the unique demography of this population as a tri-continental admixture that passed through a narrow bottleneck [55], we conducted this study with the hypothesis that rare variants with a large effect size, i.e., the *APOE* Christchurch mutation [5], could account for much of the difference in age at dementia onset. Indeed, we identified many genetic variants of a similar rarity in this study that are candidates for having a large effect on age at dementia onset. However, we note that due to the nature of the analysis, it is possible for the presence of alleles in a small number of individuals with a particularly late age at onset to result in a low *p* value and large effect size (“winner’s curse”), therefore large effect sizes in this study should be interpreted with caution.

Importantly, we also detected common and/or lower effect size variation associated with age of dementia onset in pathways and biological processes including clusterin, heparin sulfate and amyloid processing. Because many of these variants replicate or were identified in non-admixed European populations, it suggests that the associations for many of these variants are robust to ancestral background. The identified variants in this study occur in the presence of a very strong causative mutation for ADAD, emphasizing the importance of the association signals observed for these variants and the need for more investigation of these variants in future studies.

## Data Availability

All summary statistics are available as supplemental material.

http://mendel.hudsonalpha.org/Public/ncochran/Colombia/E280A-AAO-SuppTable1.tsv.gz

## Acknowledgements

We thank the patients and their caregivers for their generous contributions to this study. We thank other members of the GNA for their contributions to study of this cohort which made this study possible. We thank Nithesh Perumal for assisting with pedigree reconstruction. Replication statistics were obtained from publicly available data for NIAGADS project NG00075 [21] and controlled access data from NIAGADS projects NG00048 [19] and NG00058 [20] and the Synapse AD Knowledge portal (Synapse ID: syn25871263). More extensive acknowledgement details for DIAN, IGAP, ADDS, and ADGC are provided in the supplemental acknowledgements. The authors have no relevant conflicts of interest to disclose.

---

**Supplemental Table 1:** Summary statistics for all variants. (see supplemental zipped text file) [Note to reviewers: we will deposit this file to NIAGADS, but pre-publication, this file can be accessed via: http://mendel.hudsonalpha.org/Public/ncochran/Colombia/E280A-AAO-SuppTable1.tsv.gz]

**Supplemental Table 2:** Annotated summary statistics including replication information from described cohorts for variants with *p*<1×10^−5^, coding variants with *p*<0.05, *APOE* coding variants, and variants that are index variants for previous GWAS. (see supplemental Excel file)

**Supplemental Table 3:** Single nucleus multiomics regions from a recent study [23] that overlap GWAS variants with *p*<1×10^−5^. (see supplemental Excel file)

**Supplemental Table 4:** Full list of GWAS hits from previous studies evaluated. (see supplemental Excel file)

**Supplemental File 1:** LocusZoom plots of all regions with *p*<1×10^−5^. (see supplemental zip file)

**Supplemental Figure 1:**
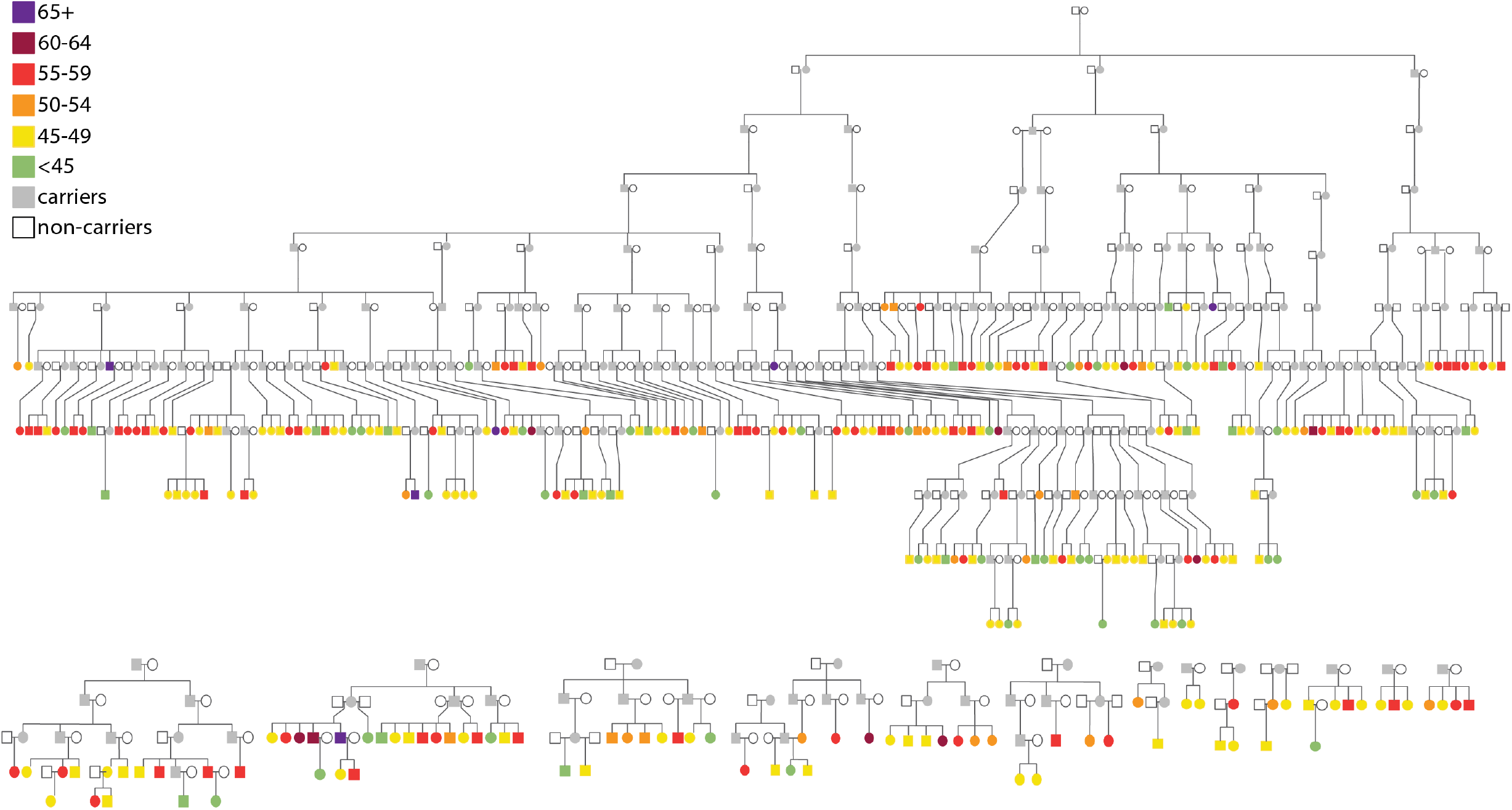
Pedigrees of included patients colored according to age at dementia onset. The E280A mutation carriers (known and presumptive) that were not included in the present study are colored in gray. The large pedigree includes 252 of the sequenced participants, and their relatedness until the most recent common ancestors (which were born in 1743–1750). The small pedigrees in the lower row represent 87 additional participants. Healthy siblings and descendants of the participants were excluded for simplicity of the pedigree.

**Supplemental Figure 2:**
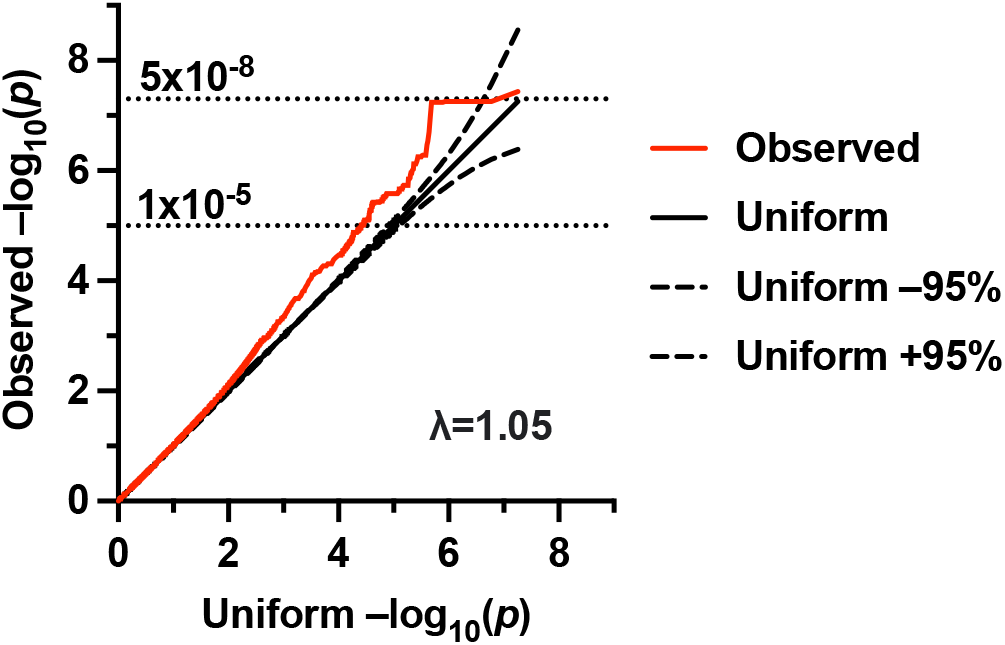
QQ plot. Observed (actual age of dementia onset) –log_10_(*p*) values are plotted vs. the uniform distribution of –log_10_(*p*) values. The genomic inflation factor (λ) is modest at 1.05.

**Supplemental Figure 3:**
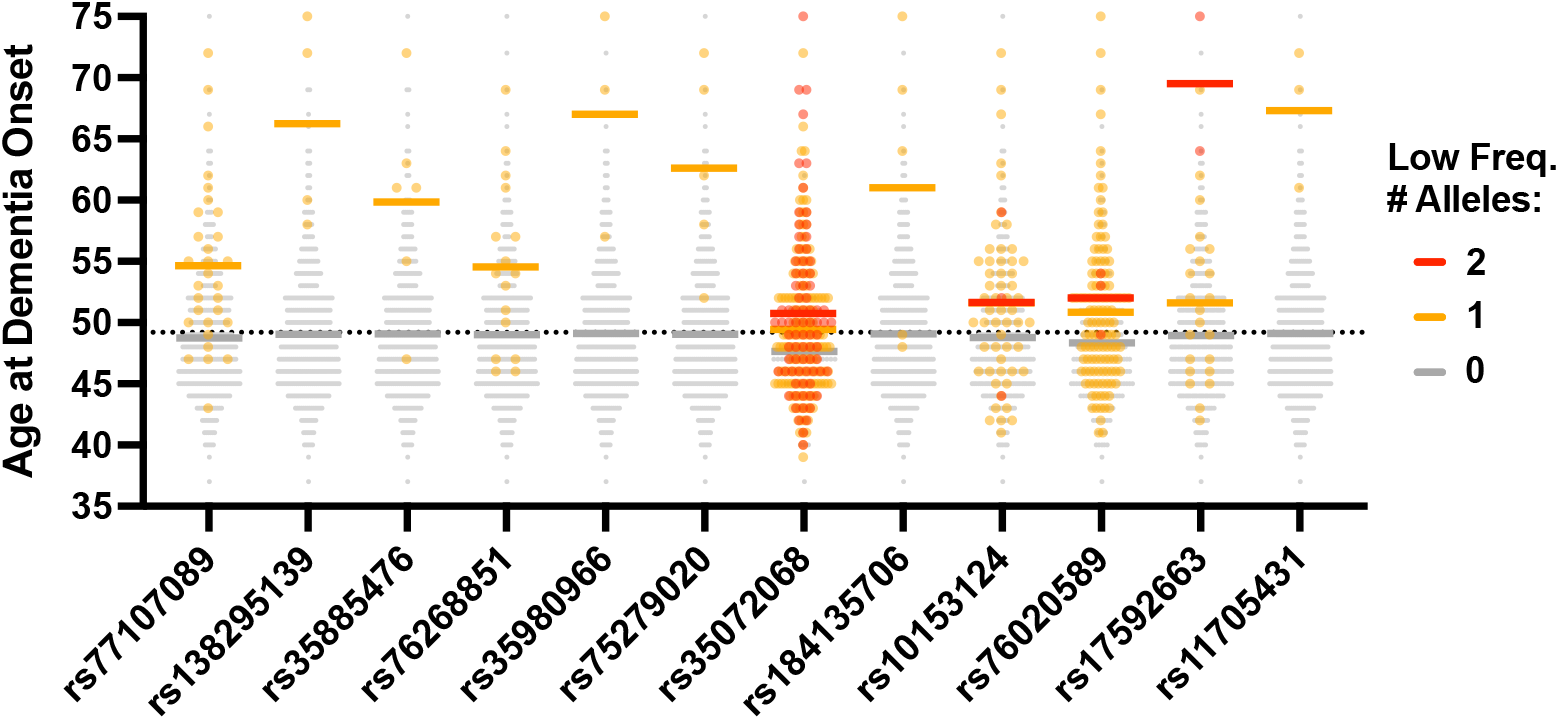
Age distribution of variants displayed in Table 1.

**Supplemental Figure 4:**
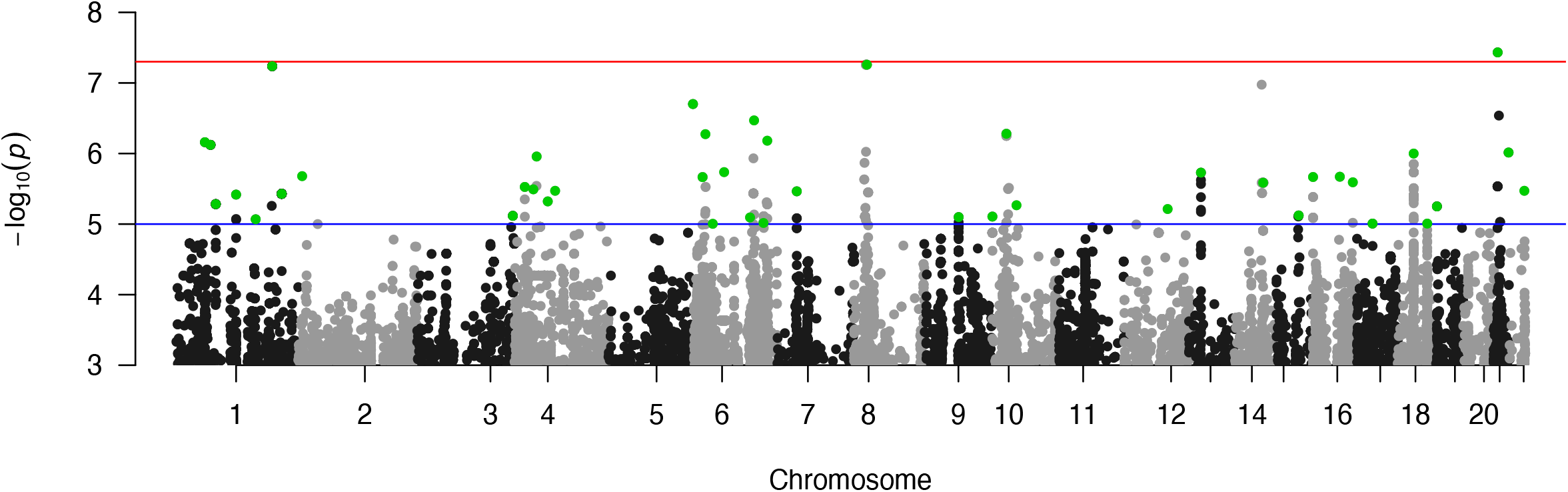
Manhattan plot. Variants selected for display in Table 1 with a discovery *p*<1×10^−5^ are highlighted in green. Blue line indicates *p*<1×10^−5^, red line indicates *p*<5×10^−8^.

