## Supplemental Text for "Genetic Associations with Age at Dementia Onset in the *PSEN1 E280A* Colombian Kindred"

### Supplemental Methods

#### *IGAP replication study details*

International Genomics of Alzheimer's Project (IGAP) is a large three-stage study based upon genome-wide association studies (GWAS) on individuals of European ancestry and served as one source of replication data. In stage 1, IGAP used genotyped and imputed data on 11,480,632 single nucleotide polymorphisms (SNPs) to meta-analyse GWAS datasets consisting of 21,982 Alzheimer's disease cases and 41,944 cognitively normal controls from four consortia: The Alzheimer Disease Genetics Consortium (ADGC); The European Alzheimer's disease Initiative (EADI); The Cohorts for Heart and Aging Research in Genomic Epidemiology Consortium (CHARGE); and The Genetic and Environmental Risk in AD Consortium Genetic and Environmental Risk in AD/Defining Genetic, Polygenic and Environmental Risk for Alzheimer's Disease Consortium (GERAD/PERADES). In stage 2, 11,632 SNPs were genotyped and tested for association in an independent set of 8,362 Alzheimer's disease cases and 10,483 controls. Meta-analysis of variants selected for analysis in stage 3A (n = 11,666) or stage 3B (n = 30,511) samples brought the final sample to 35,274 clinical and autopsy-documented Alzheimer's disease cases and 59,163 controls.

#### *Genome Sequencing and Annotation*

Genome sequencing at HudsonAlpha was conducted using the Illumina NovaSeq platform. Sequencing libraries at HudsonAlpha were prepared by Covaris shearing, end repair, adapter ligation, and PCR using standard protocols. Library concentrations were normalized using KAPA qPCR prior to sequencing. Sequencing reads were aligned to the hg19 reference genome with bwa-0.7.12 [1]. BAMs were sorted and duplicates were marked with Sambamba 0.5.4 [2]. Indels were realigned, bases were recalibrated, and gVCFs were generated with GATK 3.3 [3]. Variants were called across all samples in a single batch with GATK 3.8 using the -newQual flag to minimize false negative singleton calls. Additional genomes were sequenced previously with CompleteGenomics technology and also aligned to the hg19 reference [4]. In order to facilitate annotation, multi-allelic sites were split using Vt [5]. SnpEff 4.3s [6] was used to annotate with the gene definitions from human genome build Ensembl GRCh37.75. All variants were annotated with CADD v1.3 [7], including all indels. Population database frequency annotations included 1000 genomes phase 3, TOPMed Bravo [8] (lifted over from hg38 to hg19 using CrossMap 0.2.7 [9]), and several population database sets annotated using WGSA 0.7 [10] including ExAC [11], gnomAD [12], ESP, and UK10K. Variants were also annotated with dbSNP release 153 [13]. For intersection of array data, which was annotated with hg38, we lifted over genome sequencing data using CrossMap 0.3.4 [9].

**Supplemental Acknowledgements**

*DIAN*

Data collection and sharing for this project were supported by The Dominantly Inherited Alzheimer Network (DIAN, U19AG032438), funded by the National Institute on Aging (NIA), the Alzheimer's Association (SG-20-690363-DIAN), the German Center for Neurodegenerative Diseases (DZNE), Raul Carrea Institute for Neurological Research (FLENI), Partial support by the Research and Development Grants for Dementia from Japan Agency for Medical Research and Development, AMED, and the Korea Health Technology R&D Project through the Korea Health Industry Development Institute (KHIDI), Spanish Institute of Health Carlos III (ISCIII), Canadian Institutes of Health Research (CIHR), Canadian Consortium of Neurodegeneration and Aging, Brain Canada Foundation, and Fonds de Recherche du Québec – Santé. DIAN Study investigators have reviewed this manuscript for scientific content and consistency of data interpretation with previous DIAN Study publications. We acknowledge the altruism of the participants and their families and the contributions of the DIAN research and support staff at each of the participating sites for their contributions to this study.

*DIAN study group*

Sarah Adams, Ricardo Allegri, Aki Araki, Nicolas Barthelemy, Randall Bateman, Jacob Bechara,Tammie Benzinger, Sarah Berman, Courtney Bodge, Susan Brandon, William (Bill) Brooks, Jared

Brosch, Jill Buck, Virginia Buckles, Kathleen Carter, Lisa Cash, Charlie Chen, Jasmeer Chhatwal, Patricio Chrem, Jasmin Chua, Helena Chui, Carlos Cruchaga, Gregory S Day, Chrismary De La Cruz, Darcy Denner, Anna Diffenbacher, Aylin Dincer, Tamara Donahue, Jane Douglas, Duc Duong, Noelia Egido, Bianca Esposito, Anne Fagan, Marty Farlow, Becca Feldman, Colleen Fitzpatrick, Shaney Flores, Nick Fox, Erin Franklin, Nelly Friedrichsen, Hisako Fujii, Samantha Gardener, Bernardino Ghetti, Alison Goate, Sarah Goldberg, Jill Goldman, Alyssa Gonzalez, Brian Gordon, Susanne Gräber-Sultan, Neill Graff-Radford, Morgan Graham, Julia Gray, Emily Gremminger, Miguel Grilo, Alex Groves, Christian Haass, Lisa Häsler, Jason Hassenstab, Cortaiga Hellm, Elizabeth Herries, Laura Hoechst-Swisher, Anna Hofmann, David oltzman, Russ Hornbeck, Yakushev Igor, Ryoko Ihara, Takeshi Ikeuchi, Snezana Ikonomovic, Kenji Ishii, Clifford Jack, Gina Jerome, Erik Johnson, Mathias Jucker, Celeste Karch, Stephan Käser, Kensaku Kasuga, Sarah Keefe, William (Bill) Klunk, Robert Koeppe, Deb Koudelis, Elke Kuder-Buletta, Christoph Laske, Allan Levey, Johannes Levin, Yan Li, Oscar Lopez, Jacob Marsh, Rita Martinez, Ralph Martins, Neal Scott Mason, Colin Masters, Kwasi Mawuenyega, Austin McCullough, Eric McDade, Arlene Mejia, Estrella Morenas-Rodriguez, John Morris, James MountzMD, Cath Mummery, Neelesh Nadkarni, Akemi Nagamatsu, Katie Neimeyer, Yoshiki Niimi, James Noble, Joanne Norton, Brigitte Nuscher, Antoinette O'Connor, Ulricke Obermüller, Riddhi Patira, Richard Perrin, Lingyan Ping, Oliver Preische, Alan Renton, John Ringman, Stephen Salloway, Peter Schofield, Michio Senda, Nick Seyfried, Kristine Shady, Hiroyuki Shimada, Wendy Sigurdson, Jennifer Smith, Lori Smith, Beth Snitz, Hamid Sohrabi, Sochenda Stephens, Kevin Taddei, Sarah Thompson, Jonathan Vöglein, Peter Wang, Qing Wang, Elise Weamer, Chengjie Xiong, Jinbin Xu, and Xiong Xu.

*IGAP*

We thank the International Genomics of Alzheimer's Project (IGAP) for providing summary results data for these analyses. The investigators within IGAP contributed to the design and implementation of IGAP and/or provided data but did not participate in analysis or writing of this report. IGAP was made possible by the generous participation of the control subjects, the patients, and their families. The i–Select chips was funded by the French National Foundation on Alzheimer's disease and related disorders. EADI was supported by the

LABEX (laboratory of excellence program investment for the future) DISTALZ grant, Inserm, Institut Pasteur de Lille, Université de Lille 2 and the Lille University Hospital. GERAD/PERADES was supported by the Medical Research Council (Grant no 503480), Alzheimer's Research UK (Grant no 503176), the Wellcome Trust (Grant no 082604/2/07/Z) and German Federal Ministry of Education and Research (BMBF): Competence Network Dementia (CND) grant no 01GI0102, 01GI0711, 01GI0420. CHARGE was partly supported by the NIH/NIA grant R01 AG033193 and the NIA AG081220 and AGES contract N01–AG–12100, the NHLBI grant R01 HL105756, the Icelandic Heart Association, and the Erasmus Medical Center and Erasmus University. ADGC was supported by the NIH/NIA grants: U01 AG032984, U24 AG021886, U01 AG016976, and the Alzheimer's Association grant ADGC–10–196728.

*ADDS*

The results published here are in whole or in part based on data obtained from the AD Knowledge Portal ( https://adknowledgeportal.org ). Study data were provided by G. H. Sergievsky Center and the Taub Institute for Research for Alzheimer’s Disease and the Aging Brain at the Columbia University Irving Medical Center. Omic data collection was supported by funding from the National Institutes of Health [R56 AG061837 (Lee and Krinsky-McHale) generated GWAS, whole genome sequencing, targeted proteomic, and untargeted metabolomic data; P01HD035897 (Silverman) and R01 AG014673 (Schupf) generated demographic and clinical phenotype data] and from the Alzheimer’s Association (IIRG-08-90655 (Schupf) generated targeted proteomic data) as well as by funds from the New York State Office for People with Developmental Disabilities. Additional phenotypic data can be requested by contacting the PIs: Drs Joseph H Lee and Sharon Krinsky-McHale.

*ADGC (from ADGC website, https://www.adgenetics.org/)*

The National Institutes of Health, National Institute on Aging (NIH-NIA) supported this work through the following grants: ADGC, U01 AG032984, RC2 AG036528; Samples from the National Cell Repository for Alzheimer’s Disease (NCRAD), which receives government support under a cooperative agreement grant (U24 AG21886) awarded by the National Institute on Aging (NIA), were used in this study.

We thank contributors who collected samples used in this study, as well as patients and their families, whose help and participation made this work possible; Data for this study were prepared, archived, and distributed by the National Institute on Aging Alzheimer's Disease Data Storage Site (NIAGADS) at the University of Pennsylvania (U24-AG041689-01); NACC, U01 AG016976; NIA LOAD, U24 AG026395, R01AG041797; Banner Sun Health Research Institute P30 AG019610; Boston University, P30 AG013846, U01 AG10483, R01 CA129769, R01 MH080295, R01 AG017173, R01 AG025259, R01AG33193; Columbia University, P50 AG008702, R37 AG015473; Duke University, P30 AG028377, AG05128; Emory University, AG025688; Group Health Research Institute, UO1 AG006781, UO1 HG004610, UO1 HG006375; Indiana University, P30 AG10133; Johns Hopkins University, P50 AG005146, R01 AG020688; Massachusetts General Hospital, P50 AG005134; Mayo Clinic, P50 AG016574; Mount Sinai School of Medicine, P50 AG005138, P01 AG002219; New York University, P30 AG08051, UL1 RR029893, 5R01AG012101, 5R01AG022374, 5R01AG013616, 1RC2AG036502, 1R01AG035137; Northwestern University, P30 AG013854; Oregon Health & Science University, P30 AG008017, R01 AG026916; Rush University, P30 AG010161, R01 AG019085, R01 AG15819, R01 AG17917, R01 AG30146; TGen, R01 NS059873; University of Alabama at Birmingham, P50 AG016582; University of Arizona, R01 AG031581; University of California, Davis, P30 AG010129; University of California, Irvine, P50 AG016573; University of California, Los Angeles, P50 AG016570; University of California, San Diego, P50 AG005131; University of California, San Francisco, P50 AG023501, P01 AG019724; University of Kentucky, P30 AG028383, AG05144; University of Michigan, P50 AG008671; University of Pennsylvania, P30 AG010124; University of Pittsburgh, P50 AG005133, AG030653, AG041718, AG07562, AG02365; University of Southern California, P50 AG005142; University of Texas Southwestern, P30 AG012300; University of Miami, R01 AG027944, AG010491, AG027944, AG021547, AG019757; University of Washington, P50 AG005136; University of Wisconsin, P50 AG033514; Vanderbilt University, R01 AG019085; and Washington University, P50 AG005681, P01 AG03991.

The Kathleen Price Bryan Brain Bank at Duke University Medical Center is funded by NINDS grant # NS39764, NIMH MH60451 and by Glaxo Smith Kline. Genotyping of the TGEN2 cohort was supported by Kronos Science. The TGen series was also funded by NIA grant AG041232 to AJM and MJH, The Banner

Alzheimer's Foundation, The Johnnie B. Byrd Sr. Alzheimer's Institute, the Medical Research Council, and the state of Arizona and also includes samples from the following sites: Newcastle Brain Tissue Resource (funding via the Medical Research Council, local NHS trusts and Newcastle University), MRC London Brain Bank for Neurodegenerative Diseases (funding via the Medical Research Council),South West Dementia Brain Bank (funding via numerous sources including the Higher Education Funding Council for England (HEFCE), Alzheimer's Research Trust (ART), BRACE as well as North Bristol NHS Trust Research and Innovation Department and DeNDRoN), The Netherlands Brain Bank (funding via numerous sources including Stichting MS Research, Brain Net Europe, Hersenstichting Nederland Breinbrekend Werk, International Parkinson Fonds, Internationale Stiching Alzheimer Onderzoek), Institut de Neuropatologia, Servei Anatomia Patologica, Universitat de Barcelona.

ADNI data collection and sharing was funded by the National Institutes of Health Grant U01 AG024904 and Department of Defense award number W81XWH-12-2-0012. ADNI is funded by the National Institute on Aging, the National Institute of Biomedical Imaging and Bioengineering, and through generous contributions from the following: AbbVie, Alzheimer's Association; Alzheimer's Drug Discovery Foundation; Araclon Biotech; BioClinica, Inc.; Biogen; Bristol-Myers Squibb Company; CereSpir, Inc.; Eisai Inc.; Elan Pharmaceuticals, Inc.; Eli Lilly and Company; EuroImmun; F. Hoffmann-La Roche Ltd and its affiliated company Genentech, Inc.; Fujirebio; GE Healthcare; IXICO Ltd.; Janssen Alzheimer Immunotherapy Research & Development, LLC.; Johnson & Johnson Pharmaceutical Research & Development LLC.; Lumosity; Lundbeck; Merck & Co., Inc.; Meso Scale Diagnostics, LLC.; NeuroRx Research; Neurotrack Technologies; Novartis Pharmaceuticals Corporation; Pfizer Inc.; Piramal Imaging; Servier; Takeda Pharmaceutical Company; and Transition Therapeutics.

The Canadian Institutes of Health Research is providing funds to support ADNI clinical sites in Canada. Private sector contributions are facilitated by the Foundation for the National Institutes of Health (www.fnih.org).

The grantee organization is the Northern California Institute for Research and Education, and the study is coordinated by the Alzheimer's Disease Cooperative Study at the University of California, San Diego. ADNI data are disseminated by the Laboratory for Neuro Imaging at the University of Southern California. We thank Drs. D. Stephen Snyder and Marilyn Miller from NIA who are ex-officio ADGC members.

Support was also from the Alzheimer's Association (LAF, IIRG-08-89720; MP-V, IIRG-05-14147) and the US Department of Veterans Affairs Administration, Office of Research and Development, Biomedical Laboratory Research Program. P.S.G.-H. is supported by Wellcome Trust, Howard Hughes Medical Institute, and the Canadian Institute of Health Research.
