## Supplemental File 1 for "Genetic Associations with Age at Dementia Onset in the *PSEN1 E280A* Colombian Kindred": chr1_58029837-59029837.pdf

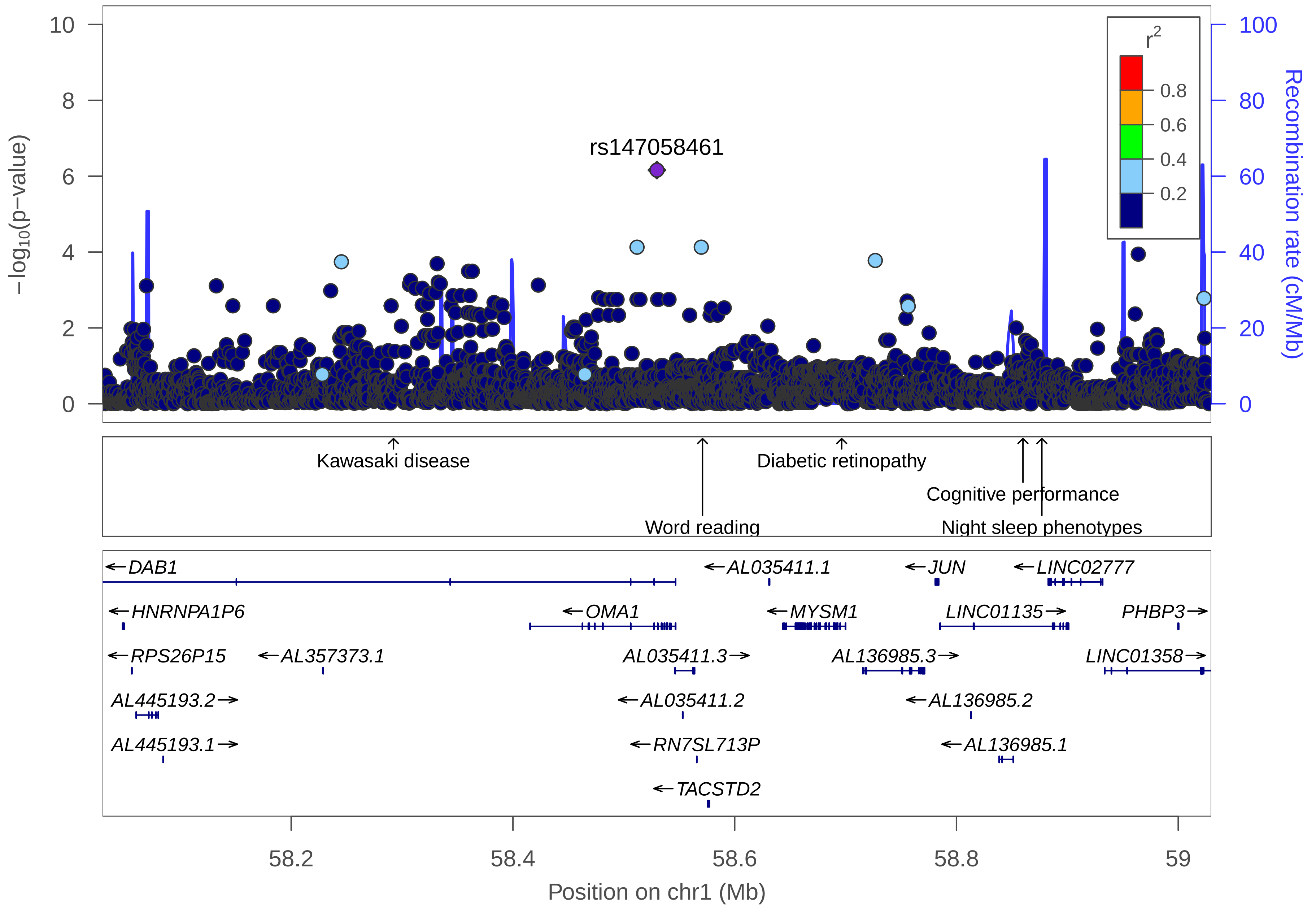

rs147058461
r2
0.8
0.6
0.4
0.2
−log10(p−value)
Recombination rate (cM/Mb)
Kawasaki disease
Diabetic retinopathy
Cognitive performance
Word reading
Night sleep phenotypes
DAB1
AL035411.1
JUN
LINC02777
HNRNPA1P6
OMA1
MYSM1
LINC01135
PHBP3
RPS26P15
AL357373.1
AL035411.3
AL136985.3
LINC01358
AL445193.2
AL035411.2
AL136985.2
AL445193.1
RN7SL713P
AL136985.1
TACSTD2
58.2
58.4
58.6
58.8
59
Position on chr1 (Mb)

| | |
|---|---|
| date: | Thu Oct 20 22:11:28 2022 |
| build: | hg38 |
| display range: | chr1:58029837−59029837 [58029837−59029837] |
| hilite range: | 0 − 0 [ 0 − 0 ] |
| reference SNP: | chr1:58529837 |
| number of SNPs plotted: | 3805 |
| min P−value: | 6.92E−7 [chr1:58529837] |
| max P−value: | 9.99E−1 [chr1:58484568] |

Make more plots at http://csg.sph.umich.edu/locuszoom/

GWAS Catalog SNPs in Region

| chr | pos (Mb) | trait | snp |
|---|---|---|---|
| 1 | 58.29224 | Kawasaki disease | rs527409 |
| 1 | 58.57095 | Word reading | rs638065 |
| 1 | 58.69648 | Diabetic retinopathy | rs2811893 |
| 1 | 58.85995 | Cognitive performance | rs4601609 |
| 1 | 58.87699 | Night sleep phenotypes | rs2764894 |
