## Supplemental File 1 for "Genetic Associations with Age at Dementia Onset in the *PSEN1 E280A* Colombian Kindred": chr1_69323901-70492002.pdf

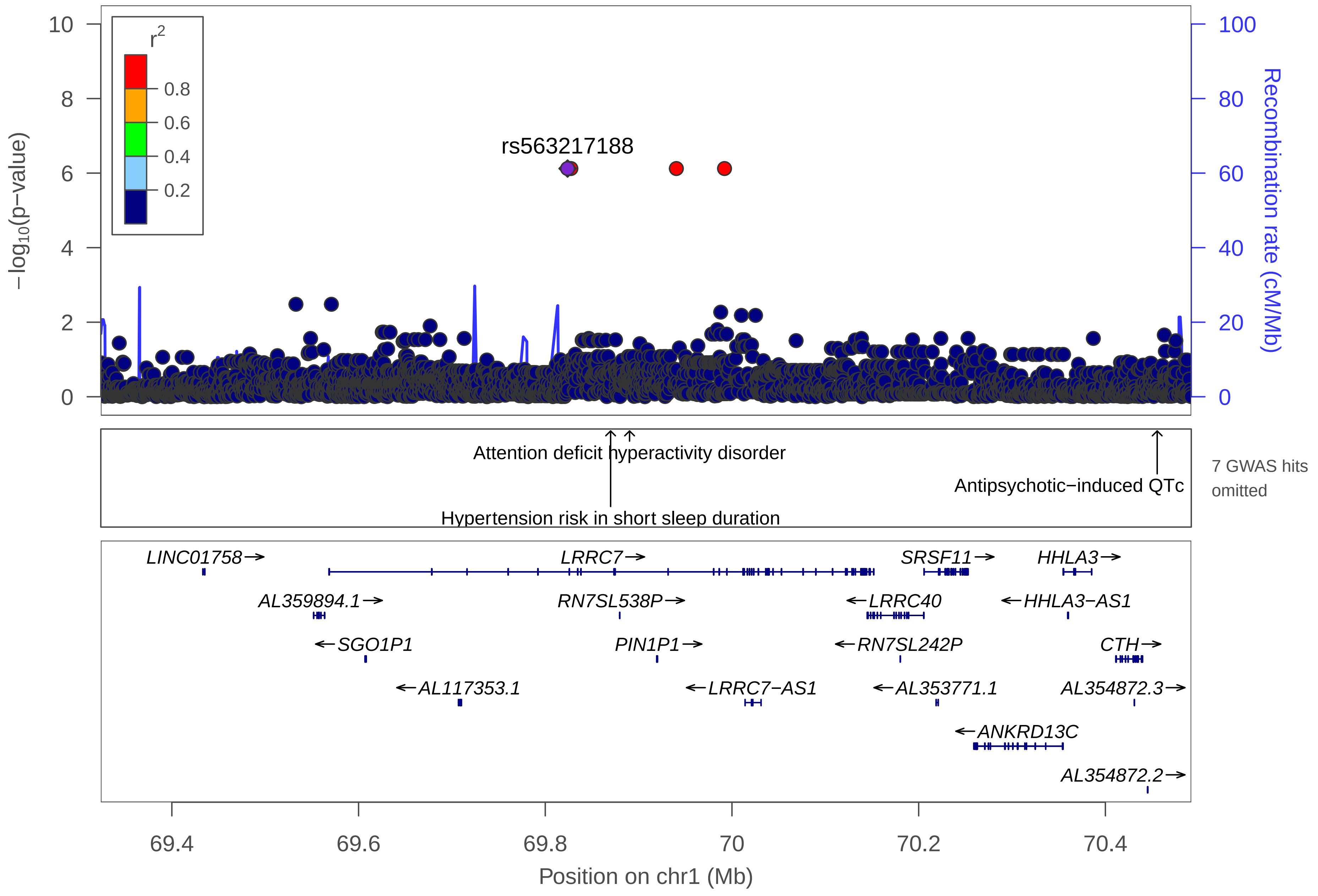

r2
0.8
0.6
0.4
0.2
rs563217188
−log10(p−value)
Recombination rate (cM/Mb)
Attention deficit hyperactivity disorder
Hypertension risk in short sleep duration
Antipsychotic−induced QTc
7 GWAS hits omitted
LINC01758→
LRRC7→
SRSF11→
HHLA3→
AL359894.1→
RN7SL538P→
←LRRC40
←HHLA3−AS1
←SGO1P1
PIN1P1→
←RN7SL242P
CTH→
←AL117353.1
←LRRC7−AS1
←AL353771.1
AL354872.3→
←ANKRD13C
AL354872.2→
69.4
69.6
69.8
70
70.2
70.4
Position on chr1 (Mb)

| | |
|---|---|
| date: | Thu Oct 20 22:13:35 2022 |
| build: | hg38 |
| display range: | chr1:69323901−70492002 [69323901−70492002] |
| hilite range: | 0 − 0 [ 0 − 0 ] |
| reference SNP: | chr1:69823901 |
| number of SNPs plotted: | 3484 |
| min P−value: | 7.54E−7 [chr1:69823901] |
| max P−value: | 10E−1 [chr1:70139284] |
| omitted GWAS Hits: | NA, NA |
| omitted GWAS Hits: | NA, NA |
| omitted GWAS Hits: | NA |

Make more plots at http://csg.sph.umich.edu/locuszoom/

GWAS Catalog SNPs in Region

| chr | pos (Mb) | trait | snp |
|---|---|---|---|
| 1 | 69.48402 | Post bronchodilator FEV1/FVC ratio | rs138006855 |
| 1 | 69.61911 | Cannabis use (initiation) | rs11809230 |
| 1 | 69.68218 | Cannabis use (age at onset) | rs181704351 |
| 1 | 69.68876 | Orofacial clefts | rs1417437 |
| 1 | 69.74140 | Body mass index | rs10889850 |
| 1 | 69.87000 | Hypertension risk in short sleep duration | rs2226284 |
| 1 | 69.89030 | Attention deficit hyperactivity disorder | rs12037173 |
| 1 | 69.93886 | Obesity−related traits | rs1023008 |
| 1 | 70.42142 | Adverse response to chemotherapy in breast cancer (alopecia) (cyclophosphamide+doxorubicin+/−5FU) | rs672203 |
| 1 | 70.45549 | Antipsychotic−induced QTc interval prolongation | rs10458561 |
