## Supplemental File 1 for "Genetic Associations with Age at Dementia Onset in the *PSEN1 E280A* Colombian Kindred": chr1_79964760-80981863.pdf

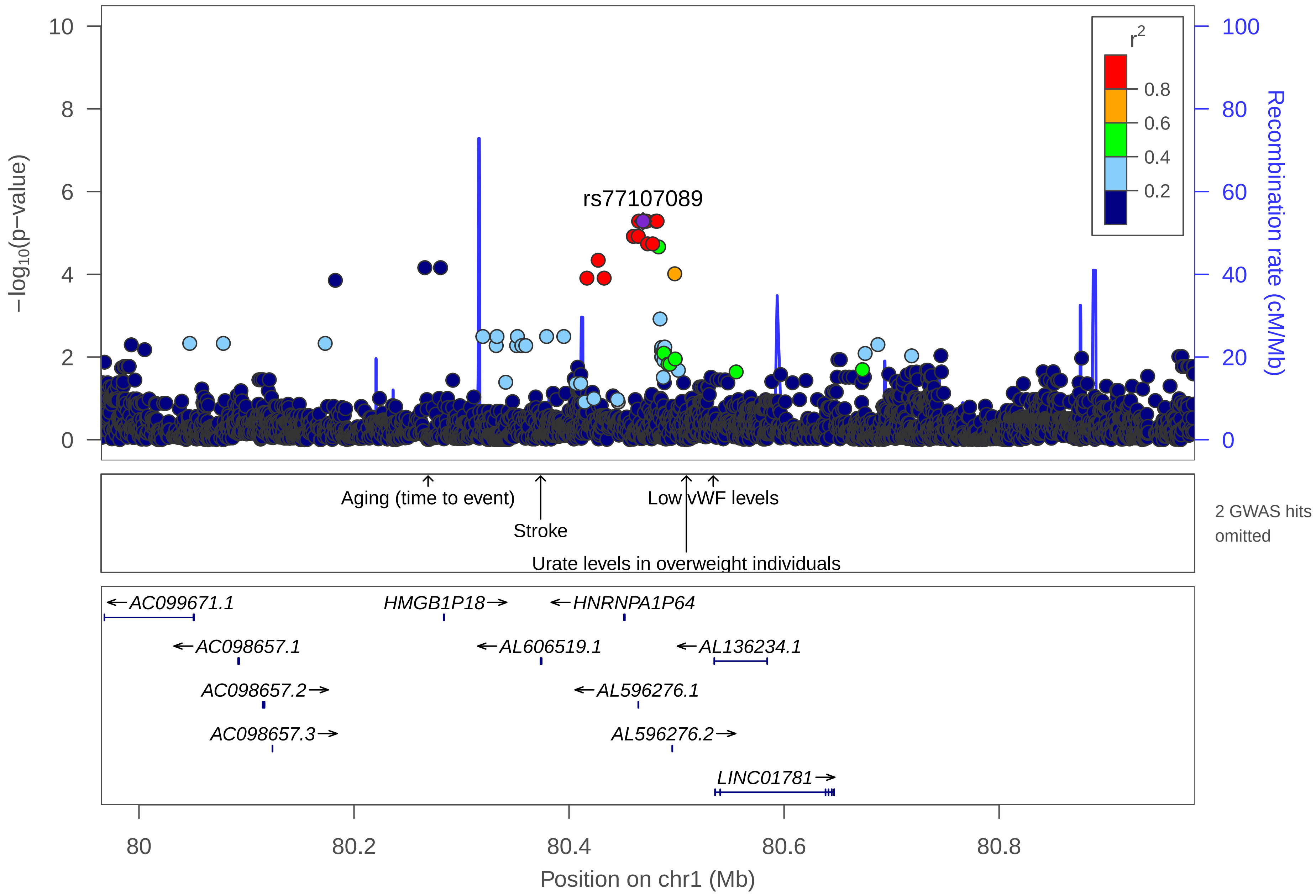

10
8
6
4
2
0
−log10(p−value)
100
80
60
40
20
0
Recombination rate (cM/Mb)
r2
0.8
0.6
0.4
0.2
rs77107089
Aging (time to event)
Stroke
Low vWF levels
Urate levels in overweight individuals
2 GWAS hits omitted
AC099671.1
HMGB1P18
HNRNPA1P64
AC098657.1
AL606519.1
AL136234.1
AC098657.2
AL596276.1
AC098657.3
AL596276.2
LINC01781
80
80.2
80.4
80.6
80.8
Position on chr1 (Mb)

| | |
|---|---|
| date: | Thu Oct 20 22:13:35 2022 |
| build: | hg38 |
| display range: | chr1:79964760−80981863 [79964760−80981863] |
| hilite range: | 0 − 0 [ 0 − 0 ] |
| reference SNP: | chr1:80468831 |
| number of SNPs plotted: | 3916 |
| min P−value: | 5.21E−6 [chr1:80464760] |
| max P−value: | 10E−1 [chr1:80809206] |
| omitted GWAS Hits: | chr1:80.534074−Low vWF levels, NA |

Make more plots at http://csg.sph.umich.edu/locuszoom/

GWAS Catalog SNPs in Region

| chr | pos (Mb) | trait | snp |
|---|---|---|---|
| 1 | 80.01220 | Obesity−related traits | rs10493631 |
| 1 | 80.26890 | Aging (time to event) | rs11162963 |
| 1 | 80.37359 | Stroke | rs1937787 |
| 1 | 80.44543 | Estradiol plasma levels (breast cancer) | rs12118390 |
| 1 | 80.50914 | Urate levels in overweight individuals | rs7539892 |
| 1 | 80.53407 | Low vWF levels | rs17398299 |
