## Supplemental File 1 for "Genetic Associations with Age at Dementia Onset in the *PSEN1 E280A* Colombian Kindred": chr1_120926988-122010262.pdf

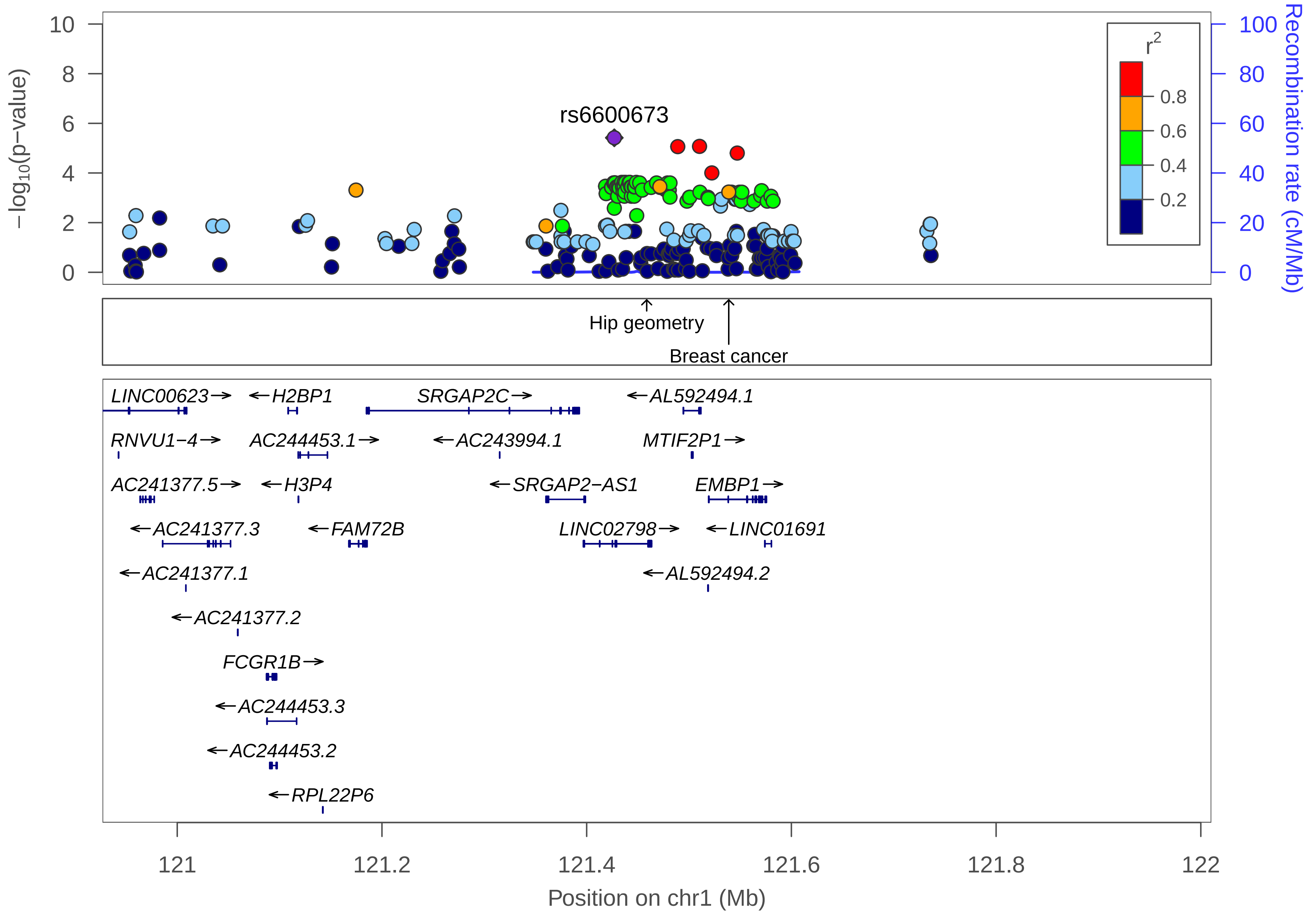
rs6600673
−log10(p−value)
Recombination rate (cM/Mb)
r2
0.8
0.6
0.4
0.2
Hip geometry
Breast cancer
LINC00623
H2BP1
SRGAP2C
AL592494.1
RNVU1−4
AC244453.1
AC243994.1
MTIF2P1
AC241377.5
H3P4
SRGAP2−AS1
EMBP1
AC241377.3
FAM72B
LINC02798
LINC01691
AC241377.1
AL592494.2
AC241377.2
FCGR1B
AC244453.3
AC244453.2
RPL22P6
Position on chr1 (Mb)

| | |
|---|---|
| date: | Thu Oct 20 22:13:31 2022 |
| build: | hg38 |
| display range: | chr1:120926988−122010262 [120926988−122010262] |
| hilite range: | 0 − 0 [ 0 − 0 ] |
| reference SNP: | chr1:121426988 |
| number of SNPs plotted: | 230 |
| min P−value: | 3.82E−6 [chr1:121426988] |
| max P−value: | 9.67E−1 [chr1:121591719] |

Make more plots at http://csg.sph.umich.edu/locuszoom/

GWAS Catalog SNPs in Region

| chr | pos (Mb) | trait | snp |
|---|---|---|---|
| 1 | 121.4586 | Hip geometry | rs6600671 |
| 1 | 121.5388 | Breast cancer | rs11249433 |
