## Supplemental File 1 for "Genetic Associations with Age at Dementia Onset in the *PSEN1 E280A* Colombian Kindred": chr1_160524526-161524526.pdf

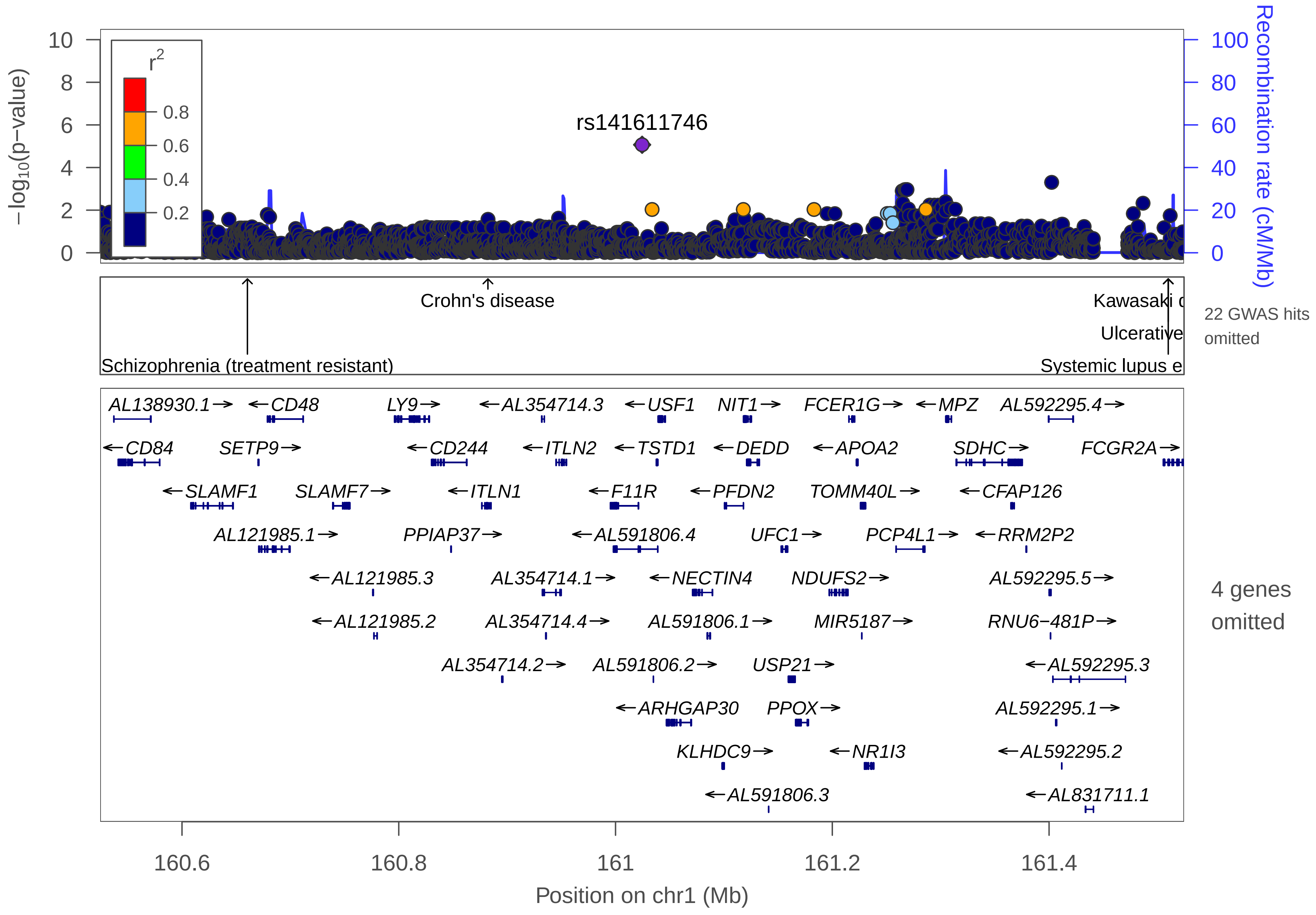

rs141611746
r2
0.8
0.6
0.4
0.2
−log10(p−value)
Recombination rate (cM/Mb)
Crohn's disease
Schizophrenia (treatment resistant)
Kawasaki d
Ulcerative
Systemic lupus e
22 GWAS hits omitted
AL138930.1→
←CD48
LY9→
←AL354714.3
←USF1
NIT1→
FCER1G→
←MPZ
AL592295.4→
←CD84
SETP9→
←CD244
←ITLN2
←TSTD1
←DEDD
←APOA2
SDHC→
FCGR2A→
←SLAMF1
SLAMF7→
←ITLN1
←F11R
←PFDN2
TOMM40L→
←CFAP126
AL121985.1→
PPIAP37→
←AL591806.4
UFC1→
PCP4L1→
←RRM2P2
←AL121985.3
AL354714.1→
←NECTIN4
NDUFS2→
AL592295.5→
←AL121985.2
AL354714.4→
AL591806.1→
MIR5187→
RNU6−481P→
AL354714.2→
AL591806.2→
USP21→
←AL592295.3
←ARHGAP30
PPOX→
AL592295.1→
KLHDC9→
←NR1I3
←AL592295.2
←AL591806.3
←AL831711.1
4 genes omitted
160.6
160.8
161
161.2
161.4
Position on chr1 (Mb)

| | |
|---|---|
| date: | Thu Oct 20 22:13:36 2022 |
| build: | hg38 |
| display range: | chr1:160524526−161524526 [160524526−161524526] |
| hilite range: | 0 − 0 [ 0 − 0 ] |
| reference SNP: | chr1:161024526 |
| number of SNPs plotted: | 2999 |
| min P−value: | 8.55E−6 [chr1:161024526] |
| max P−value: | 10E−1 [chr1:161070828] |
| omitted Genes: | AL590714.1, B4GALT3, ADAMTS4 |
| omitted Genes: | AL590385.2 |
| omitted GWAS Hits: | NA, NA |
| omitted GWAS Hits: | NA, NA |
| omitted GWAS Hits: | NA, NA |
| omitted GWAS Hits: | NA, NA |
| omitted GWAS Hits: | NA, NA |
| omitted GWAS Hits: | NA, NA |
| omitted GWAS Hits: | NA, NA |
| omitted GWAS Hits: | NA |

Make more plots at http://csg.sph.umich.edu/locuszoom/

GWAS Catalog SNPs in Region

| chr | pos (Mb) | trait | snp |
|---|---|---|---|
| 1 | 160.5465 | Response to anti−TNF therapy in rheumatoid arthritis | rs6427528 |
| 1 | 160.6604 | Schizophrenia (treatment resistant) | rs11265461 |
| 1 | 160.6733 | Microalbuminuria | rs3795324 |
| 1 | 160.7740 | Cervical cancer | rs12068654 |
| 1 | 160.7763 | Capecitabine sensitivity | rs576523 |
| 1 | 160.8605 | Crohn's disease | rs4656940 |
| 1 | 160.8823 | Crohn's disease | rs2274910 |
| 1 | 160.8847 | Inflammatory bowel disease | rs2297559 |
| 1 | 160.8872 | Ulcerative colitis | rs4656958 |
| 1 | 160.8872 | Crohn's disease | rs4656958 |
| 1 | 160.8872 | Inflammatory bowel disease | rs4656958 |
| 1 | 161.0567 | Night sleep phenotypes | rs12130871 |
| 1 | 161.0638 | Night sleep phenotypes | rs12135996 |
| 1 | 161.2205 | Metabolite levels (lipoprotein measures) | rs4503368 |
| 1 | 161.2307 | Blood metabolite levels | rs4073054 |
| 1 | 161.3027 | Visceral adipose tissue adjusted for BMI | rs4657015 |
| 1 | 161.4353 | Rheumatoid arthritis | rs72717009 |
| 1 | 161.4938 | Functional impairment in major depressive disorder, bipolar disorder and schizophrenia | rs7535475 |
| 1 | 161.5024 | Ulcerative colitis | rs10800309 |
| 1 | 161.5024 | Inflammatory bowel disease | rs10800309 |
| 1 | 161.5090 | Systemic lupus erythematosus | rs6671847 |

GWAS Catalog SNPs in Region

| chr | pos (Mb) | trait | snp |
|---|---|---|---|
| 1 | 161.5100 | Kawasaki disease | rs1801274 |
| 1 | 161.5100 | Ulcerative colitis | rs1801274 |
| 1 | 161.5100 | Systemic lupus erythematosus | rs1801274 |
| 1 | 161.5100 | Crohn's disease | rs1801274 |
| 1 | 161.5100 | Inflammatory bowel disease | rs1801274 |
| 1 | 161.5144 | Helicobacter pylori serologic status | rs368433 |
