## Supplemental File 1 for "Genetic Associations with Age at Dementia Onset in the *PSEN1 E280A* Colombian Kindred": chr1_192937542-194892636.pdf

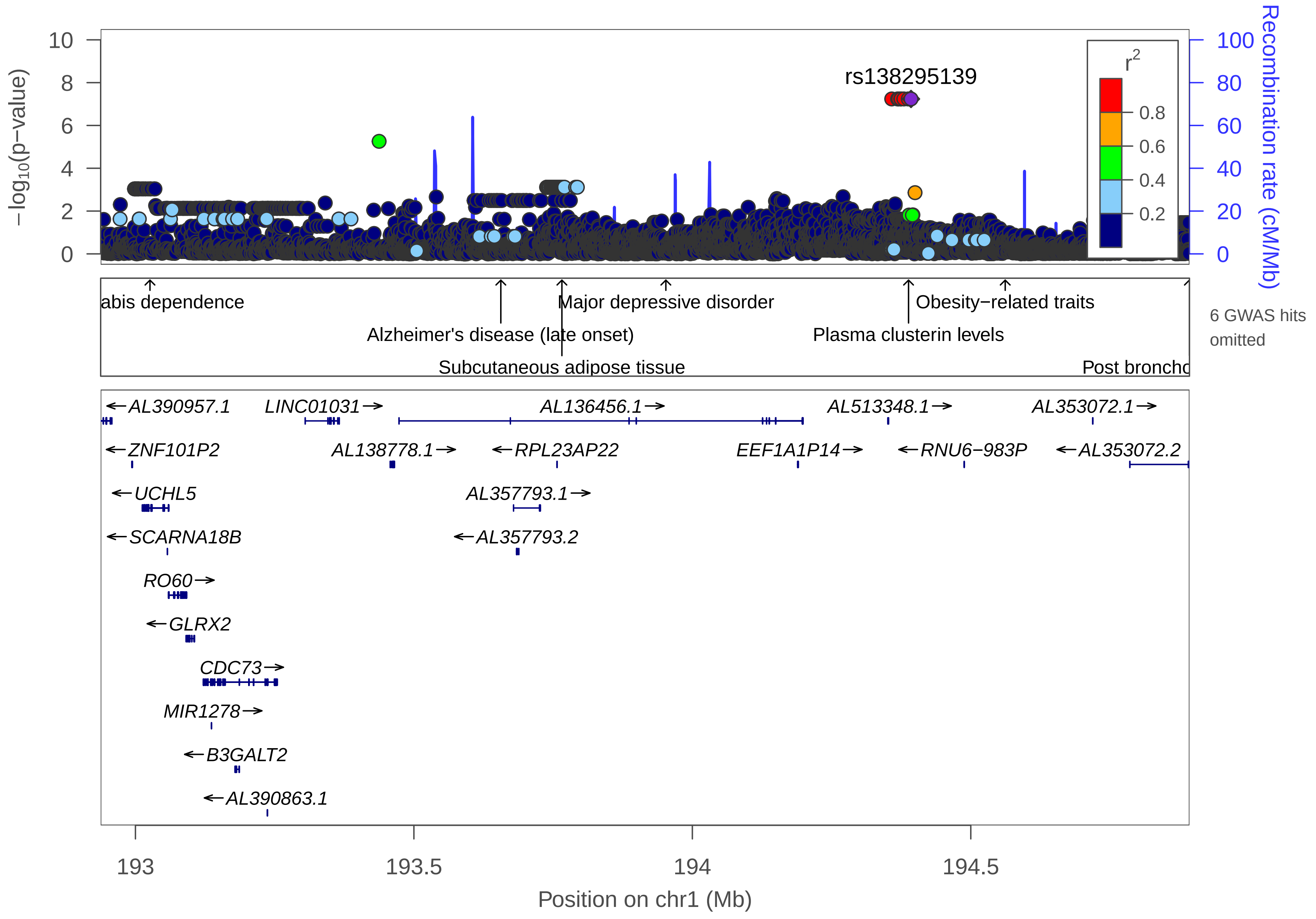

rs138295139
−log10(p−value)
Recombination rate (cM/Mb)
r2
0.8
0.6
0.4
0.2
abis dependence
Alzheimer's disease (late onset)
Subcutaneous adipose tissue
Major depressive disorder
Plasma clusterin levels
Obesity−related traits
Post broncho
6 GWAS hits omitted
AL390957.1
LINC01031
AL136456.1
AL513348.1
AL353072.1
ZNF101P2
AL138778.1
RPL23AP22
EEF1A1P14
RNU6−983P
AL353072.2
UCHL5
AL357793.1
SCARNA18B
AL357793.2
RO60
GLRX2
CDC73
MIR1278
B3GALT2
AL390863.1
193
193.5
194
194.5
Position on chr1 (Mb)

| | |
|---|---|
| date: | Thu Oct 20 22:13:36 2022 |
| build: | hg38 |
| display range: | chr1:192937542−194892636 [192937542−194892636] |
| hilite range: | 0 − 0 [ 0 − 0 ] |
| reference SNP: | chr1:194392636 |
| number of SNPs plotted: | 6800 |
| min P−value: | 5.78E−8 [chr1:194358014] |
| max P−value: | 9.99E−1 [chr1:193400222] |
| omitted GWAS Hits: | chr1:194.388206−Plasma clusterin levels, NA |
| omitted GWAS Hits: | NA, NA |

Make more plots at http://csg.sph.umich.edu/locuszoom/

GWAS Catalog SNPs in Region

| chr | pos (Mb) | trait | snp |
|---|---|---|---|
| 1 | 193.0261 | Cannabis dependence | rs9427573 |
| 1 | 193.6561 | Alzheimer's disease (late onset) | rs6678275 |
| 1 | 193.6705 | Post bronchodilator FEV1/FVC ratio | rs149819415 |
| 1 | 193.7658 | Subcutaneous adipose tissue | rs2025934 |
| 1 | 193.7936 | Daytime sleep phenotypes | rs113943039 |
| 1 | 193.9524 | Major depressive disorder | rs606149 |
| 1 | 193.9933 | White matter hyperintensity burden | rs71642944 |
| 1 | 194.3882 | Plasma clusterin levels | rs4428865 |
| 1 | 194.5618 | Obesity−related traits | rs3001167 |
| 1 | 194.5799 | Post bronchodilator FEV1 | rs79237515 |
| 1 | 194.7204 | Post bronchodilator FEV1 | rs138325697 |
| 1 | 194.7899 | Post bronchodilator FEV1 | rs144324177 |
| 1 | 194.8925 | Post bronchodilator FEV1 | rs144276489 |
