## Supplemental File 1 for "Genetic Associations with Age at Dementia Onset in the *PSEN1 E280A* Colombian Kindred": chr1_212926960-213964736.pdf

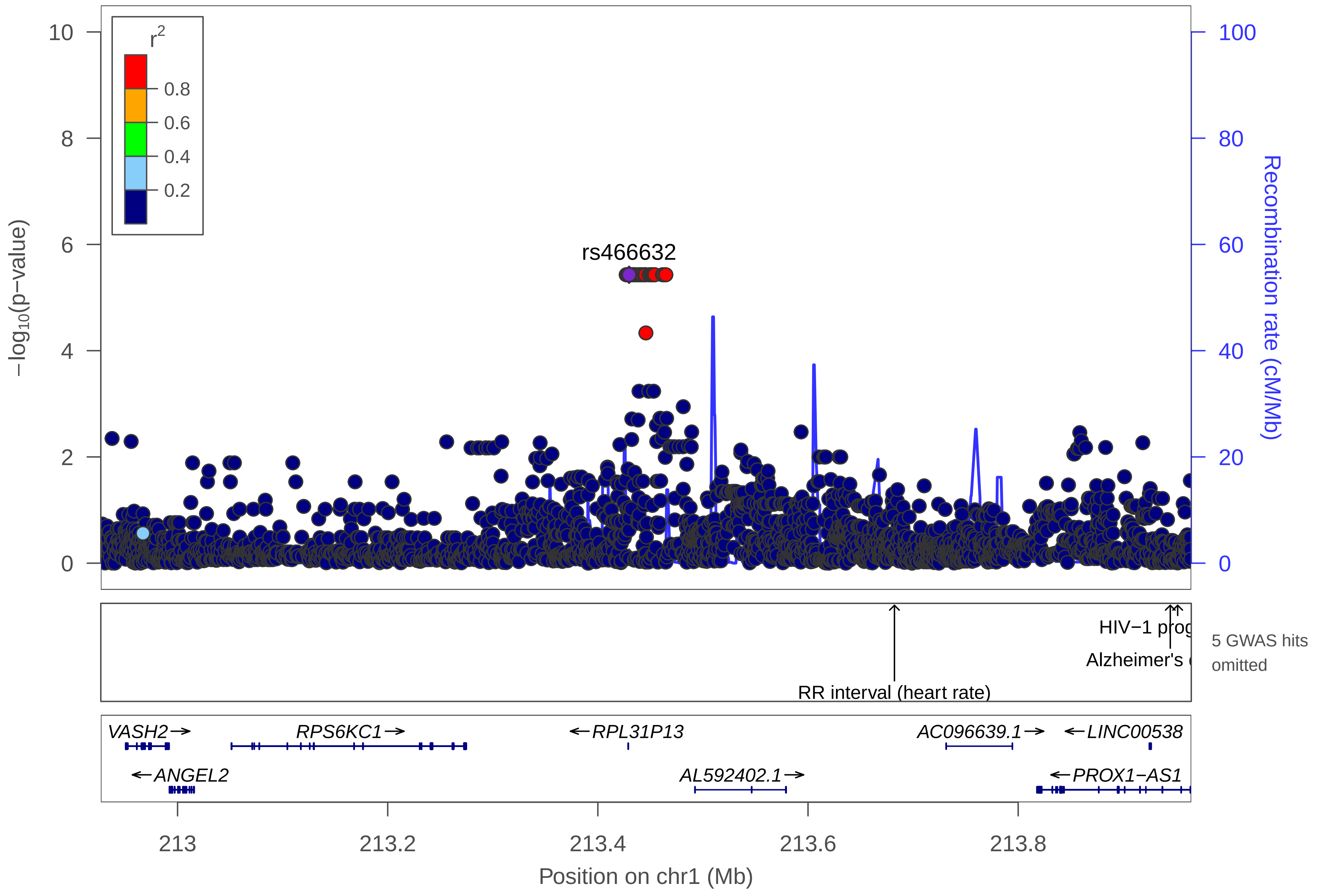
rs466632
r2
0.8
0.6
0.4
0.2
−log10(p−value)
Recombination rate (cM/Mb)
HIV−1 prog
Alzheimer's
RR interval (heart rate)
5 GWAS hits omitted
VASH2→
RPS6KC1→
←RPL31P13
AC096639.1→
←LINC00538
←ANGEL2
AL592402.1→
←PROX1−AS1
213
213.2
213.4
213.6
213.8
Position on chr1 (Mb)

| | |
|---|---|
| date: | Thu Oct 20 22:13:35 2022 |
| build: | hg38 |
| display range: | chr1:212926960−213964736 [212926960−213964736] |
| hilite range: | 0 − 0 [ 0 − 0 ] |
| reference SNP: | chr1:213429709 |
| number of SNPs plotted: | 2696 |
| min P−value: | 3.72E−6 [chr1:213426960] |
| max P−value: | 9.99E−1 [chr1:213699057] |
| omitted GWAS Hits: | NA, NA |
| omitted GWAS Hits: | NA, NA |

Make more plots at http://csg.sph.umich.edu/locuszoom/

GWAS Catalog SNPs in Region

| chr | pos (Mb) | trait | snp |
|---|---|---|---|
| 1 | 212.9532 | Eye color | rs3002288 |
| 1 | 213.4245 | Suicide | rs320461 |
| 1 | 213.6613 | Metabolite levels (5−HIAA) | rs4655303 |
| 1 | 213.6822 | RR interval (heart rate) | rs17706439 |
| 1 | 213.7372 | Obesity | rs1704198 |
| 1 | 213.9324 | Post bronchodilator FEV1/FVC ratio | rs182234674 |
| 1 | 213.9447 | Alzheimer's disease | rs340849 |
| 1 | 213.9518 | HIV−1 progression | rs17762192 |
