## Supplemental File 1 for "Genetic Associations with Age at Dementia Onset in the *PSEN1 E280A* Colombian Kindred": chr2_8594003-9594003.pdf

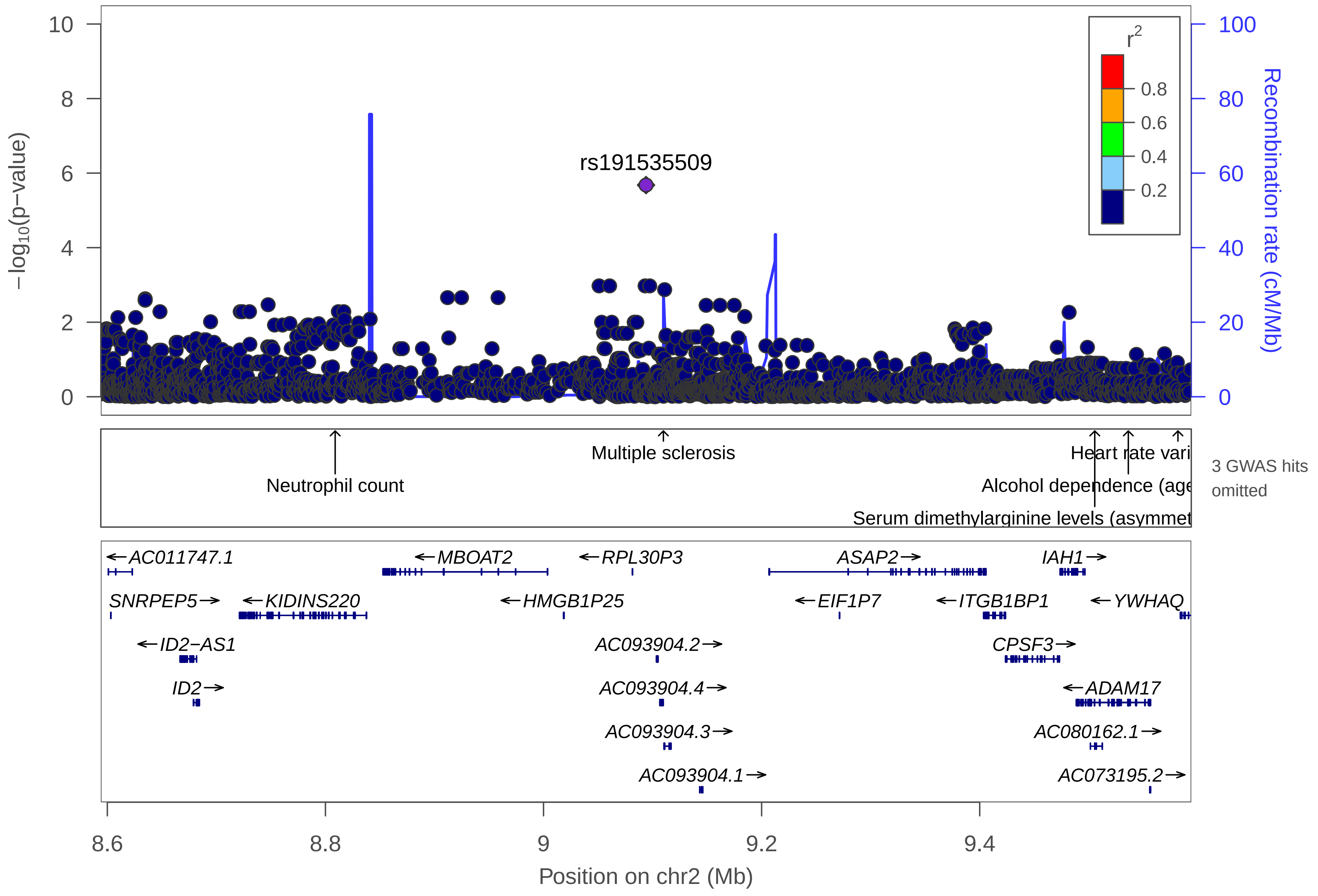

rs191535509
r2
0.8
0.6
0.4
0.2
−log10(p−value)
Recombination rate (cM/Mb)
Neutrophil count
Multiple sclerosis
Heart rate vari
Alcohol dependence (age
Serum dimethylarginine levels (asymmet
3 GWAS hits omitted
AC011747.1
MBOAT2
RPL30P3
ASAP2
IAH1
SNRPEP5
KIDINS220
HMGB1P25
EIF1P7
ITGB1BP1
YWHAQ
ID2−AS1
AC093904.2
CPSF3
ID2
AC093904.4
ADAM17
AC093904.3
AC080162.1
AC093904.1
AC073195.2
8.6
8.8
9
9.2
9.4
Position on chr2 (Mb)

| | |
|---|---|
| date: | Thu Oct 20 22:13:33 2022 |
| build: | hg38 |
| display range: | chr2:8594003−9594003 [8594003−9594003] |
| hilite range: | 0 − 0 [ 0 − 0 ] |
| reference SNP: | chr2:9094003 |
| number of SNPs plotted: | 2572 |
| min P−value: | 2.09E−6 [chr2:9094003] |
| max P−value: | 9.99E−1 [chr2:8622782] |
| omitted GWAS Hits: | NA, NA |

Make more plots at http://csg.sph.umich.edu/locuszoom/

GWAS Catalog SNPs in Region

| chr | pos (Mb) | trait | snp |
|---|---|---|---|
| 2 | 8.598773 | Schizophrenia | rs4669297 |
| 2 | 8.808940 | Neutrophil count | rs7587928 |
| 2 | 9.109909 | Multiple sclerosis | rs1109670 |
| 2 | 9.119469 | IgG glycosylation | rs9636252 |
| 2 | 9.151105 | Diisocyanate−induced asthma | rs73912949 |
| 2 | 9.505503 | Serum dimethylarginine levels (asymmetric/symetric ratio) | rs55794209 |
| 2 | 9.536284 | Alcohol dependence (age at onset) | rs17362650 |
| 2 | 9.581767 | Heart rate variability traits | rs6432018 |
