## Supplemental File 1 for "Genetic Associations with Age at Dementia Onset in the *PSEN1 E280A* Colombian Kindred": chr3_194100199-195100608.pdf

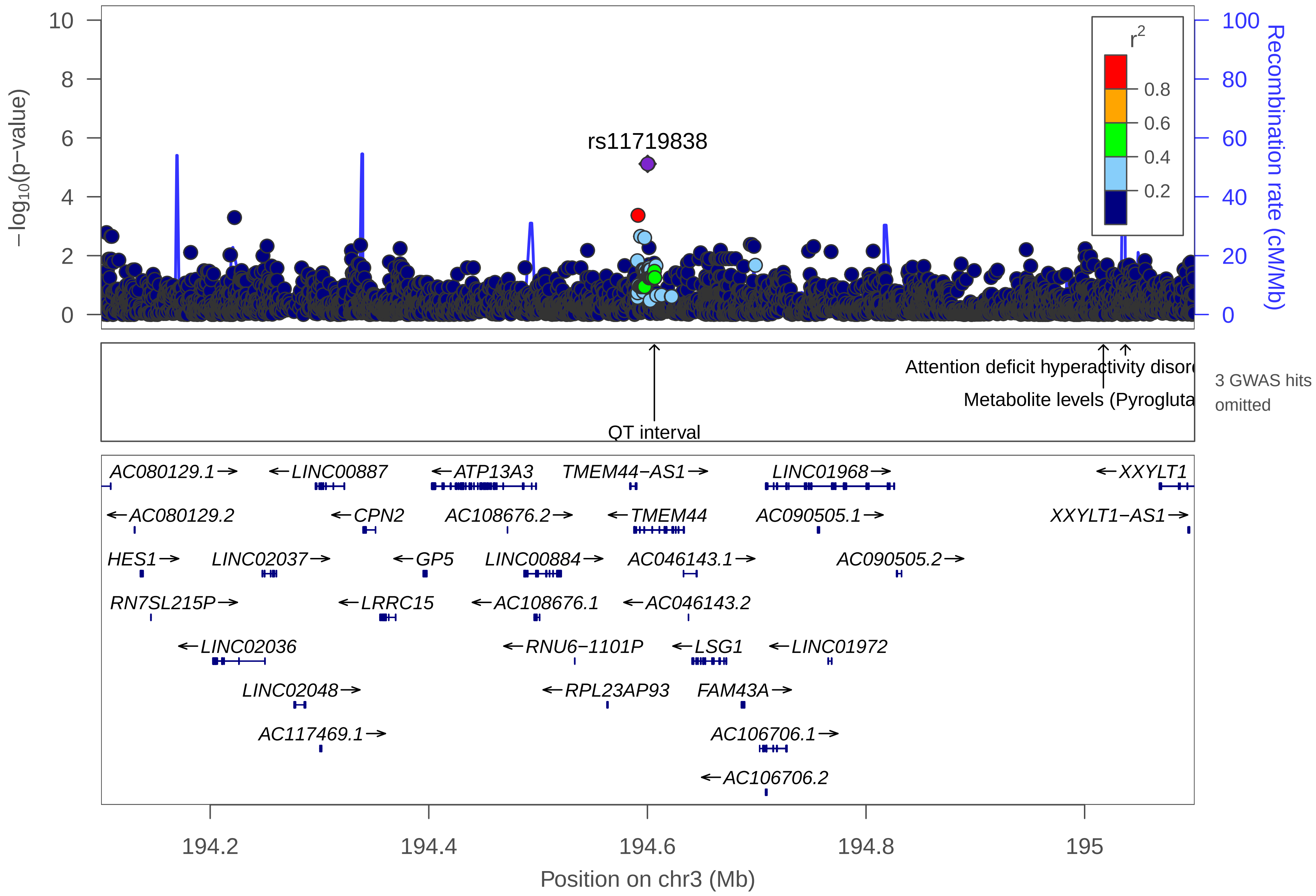

rs11719838
r2
0.8
0.6
0.4
0.2
−log10(p−value)
Recombination rate (cM/Mb)
Attention deficit hyperactivity disord
Metabolite levels (Pyrogluta
QT interval
3 GWAS hits omitted
AC080129.1→
←LINC00887
←ATP13A3
TMEM44−AS1→
LINC01968→
←XXYLT1
←AC080129.2
←CPN2
AC108676.2→
←TMEM44
AC090505.1→
XXYLT1−AS1→
HES1→
LINC02037→
←GP5
LINC00884→
AC046143.1→
AC090505.2→
RN7SL215P→
←LRRC15
←AC108676.1
←AC046143.2
←LINC02036
←RNU6−1101P
←LSG1
←LINC01972
LINC02048→
←RPL23AP93
FAM43A→
AC117469.1→
AC106706.1→
←AC106706.2
194.2
194.4
194.6
194.8
195
Position on chr3 (Mb)

| | |
|---|---|
| date: | Thu Oct 20 22:13:36 2022 |
| build: | hg38 |
| display range: | chr3:194100199−195100608 [194100199−195100608] |
| hilite range: | 0 − 0 [ 0 − 0 ] |
| reference SNP: | chr3:194600199 |
| number of SNPs plotted: | 4921 |
| min P−value: | 7.63E−6 [chr3:194600199] |
| max P−value: | 10E−1 [chr3:194346011] |
| omitted GWAS Hits: | NA, NA |

Make more plots at http://csg.sph.umich.edu/locuszoom/

GWAS Catalog SNPs in Region

| chr | pos (Mb) | trait | snp |
|---|---|---|---|
| 3 | 194.3098 | Mortality in sepsis | rs10933728 |
| 3 | 194.5172 | 3−hydroxypropylmercapturic acid levels in smokers | rs147484948 |
| 3 | 194.6064 | QT interval | rs789852 |
| 3 | 194.6852 | antipsychotic drug dosage in schizophrenia or schizoaffective disorder | rs789859 |
| 3 | 195.0171 | Metabolite levels (Pyroglutamine) | rs2676917 |
| 3 | 195.0373 | Attention deficit hyperactivity disorder (time to onset) | rs3892715 |
