## Supplemental File 1 for "Genetic Associations with Age at Dementia Onset in the *PSEN1 E280A* Colombian Kindred": chr4_20341970-21497591.pdf

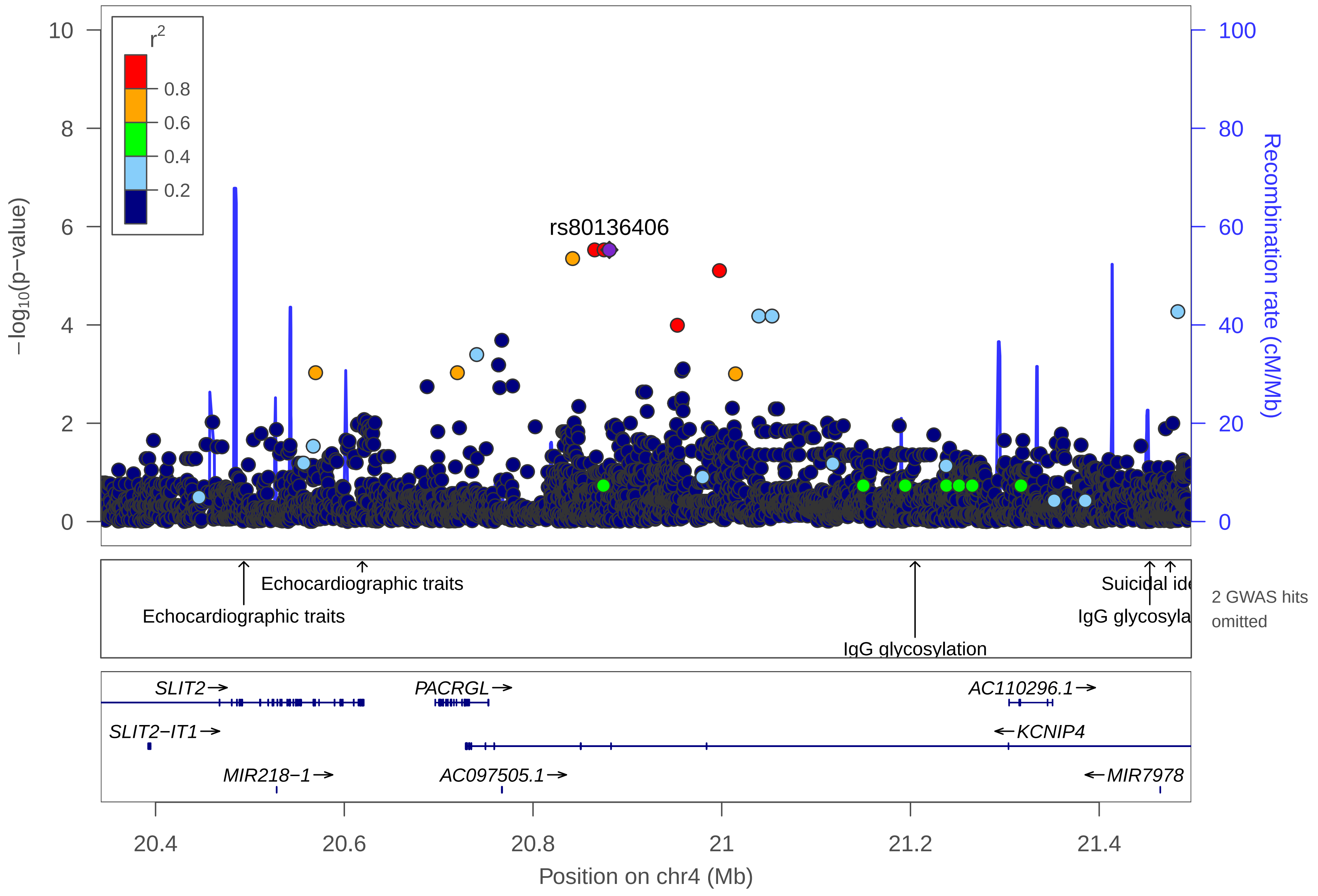

r2
0.8
0.6
0.4
0.2
rs80136406
10
8
6
4
2
0
100
80
60
40
20
0
−log10(p−value)
Recombination rate (cM/Mb)
Echocardiographic traits
Echocardiographic traits
Suicidal ide
IgG glycosyla
IgG glycosylation
2 GWAS hits omitted
SLIT2→
PACRGL→
AC110296.1→
SLIT2−IT1→
←KCNIP4
MIR218−1→
AC097505.1→
←MIR7978
20.4
20.6
20.8
21
21.2
21.4
Position on chr4 (Mb)

| | |
|---|---|
| date: | Thu Oct 20 22:13:34 2022 |
| build: | hg38 |
| display range: | chr4:20341970−21497591 [20341970−21497591] |
| hilite range: | 0 − 0 [ 0 − 0 ] |
| reference SNP: | chr4:20880902 |
| number of SNPs plotted: | 4407 |
| min P−value: | 2.98E−6 [chr4:20865311] |
| max P−value: | 10E−1 [chr4:21449638] |
| omitted GWAS Hits: | NA, NA |

Make more plots at http://csg.sph.umich.edu/locuszoom/

GWAS Catalog SNPs in Region

| chr | pos (Mb) | trait | snp |
|---|---|---|---|
| 4 | 20.49351 | Echocardiographic traits | rs666088 |
| 4 | 20.61906 | Echocardiographic traits | rs1379659 |
| 4 | 21.20487 | IgG glycosylation | rs1023721 |
| 4 | 21.29181 | Parkinson disease and lewy body pathology | rs141863958 |
| 4 | 21.39199 | Cough in response to angiotensin−converting enzyme inhibitor drugs | rs1495509 |
| 4 | 21.45363 | IgG glycosylation | rs11942476 |
| 4 | 21.47537 | Suicidal ideation | rs358592 |
