## Supplemental File 1 for "Genetic Associations with Age at Dementia Onset in the *PSEN1 E280A* Colombian Kindred": chr4_37666618-38671945.pdf

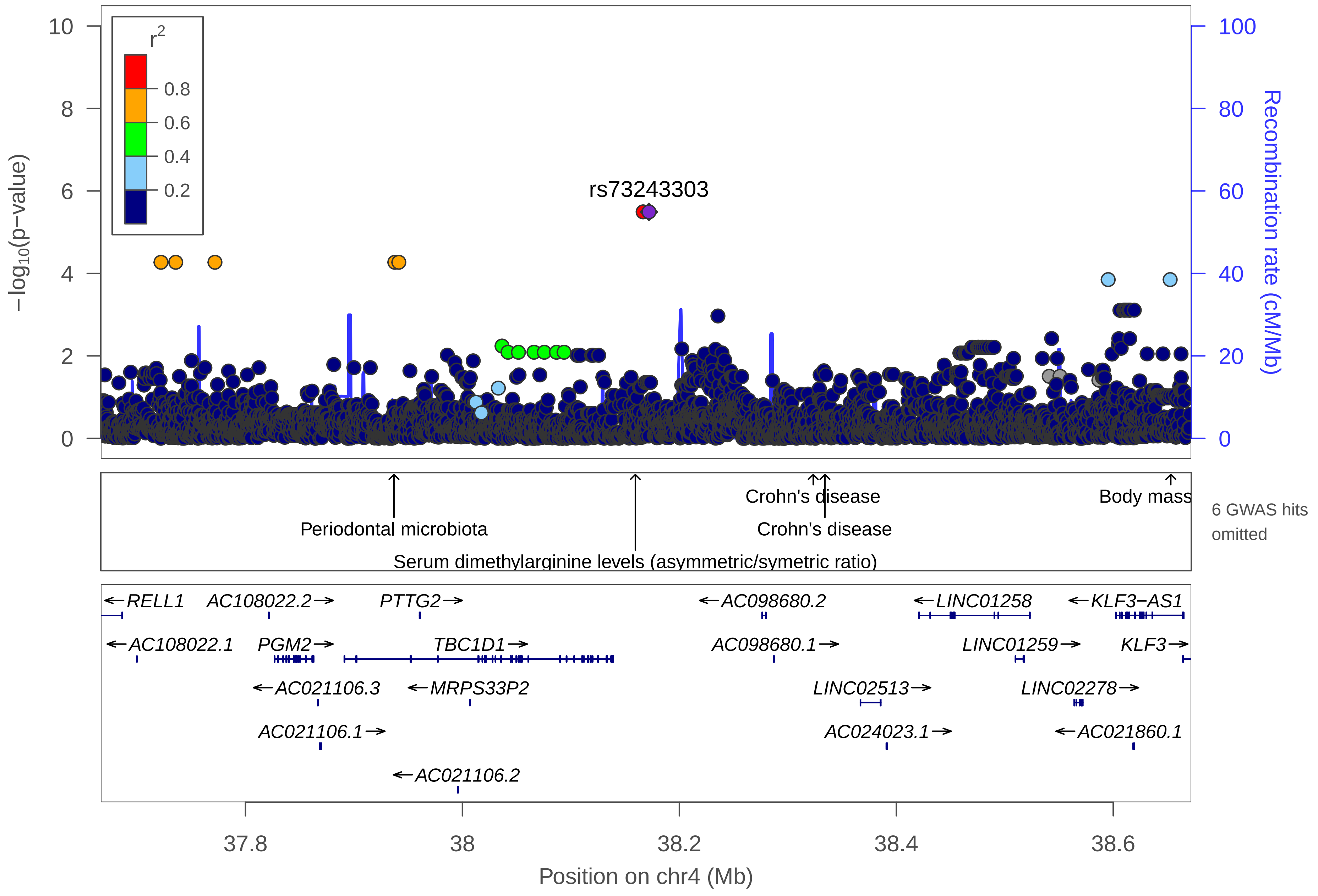

r2
0.8
0.6
0.4
0.2
rs73243303
−log10(p−value)
Recombination rate (cM/Mb)
Periodontal microbiota
Serum dimethylarginine levels (asymmetric/symetric ratio)
Crohn's disease
Crohn's disease
Body mass
6 GWAS hits omitted
←RELL1
AC108022.2→
PTTG2→
←AC098680.2
←LINC01258
←KLF3−AS1
←AC108022.1
PGM2→
TBC1D1→
AC098680.1→
LINC01259→
KLF3→
←AC021106.3
←MRPS33P2
LINC02513→
LINC02278→
AC021106.1→
AC024023.1→
←AC021860.1
←AC021106.2
37.8
38
38.2
38.4
38.6
Position on chr4 (Mb)

| | |
|---|---|
| date: | Thu Oct 20 22:13:35 2022 |
| build: | hg38 |
| display range: | chr4:37666618−38671945 [37666618−38671945] |
| hilite range: | 0 − 0 [ 0 − 0 ] |
| reference SNP: | chr4:38171945 |
| number of SNPs plotted: | 4373 |
| min P−value: | 3.21E−6 [chr4:38166618] |
| max P−value: | 9.98E−1 [chr4:37864389] |
| omitted GWAS Hits: | chr4:38.65306−Body mass index, NA |
| omitted GWAS Hits: | NA, NA |

Make more plots at http://csg.sph.umich.edu/locuszoom/

GWAS Catalog SNPs in Region

| chr | pos (Mb) | trait | snp |
|---|---|---|---|
| 4 | 37.73637 | Staphylococcus aureus nasal carriage (intermittent) | rs16993852 |
| 4 | 37.93690 | Periodontal microbiota | rs10010758 |
| 4 | 38.01141 | Verbal declarative memory | rs13132184 |
| 4 | 38.15944 | Serum dimethylarginine levels (asymmetric/symetric ratio) | rs115426111 |
| 4 | 38.15944 | Symmetrical dimethylarginine levels | rs115426111 |
| 4 | 38.32341 | Crohn's disease | rs6856616 |
| 4 | 38.32341 | Inflammatory bowel disease | rs6856616 |
| 4 | 38.33420 | Crohn's disease | rs1487630 |
| 4 | 38.46385 | Obesity−related traits | rs4615179 |
| 4 | 38.50312 | Post bronchodilator FEV1/FVC ratio in COPD | rs142846286 |
| 4 | 38.65306 | Body mass index | rs4833079 |
