## Supplemental File 1 for "Genetic Associations with Age at Dementia Onset in the *PSEN1 E280A* Colombian Kindred": chr4_44041725-45316634.pdf

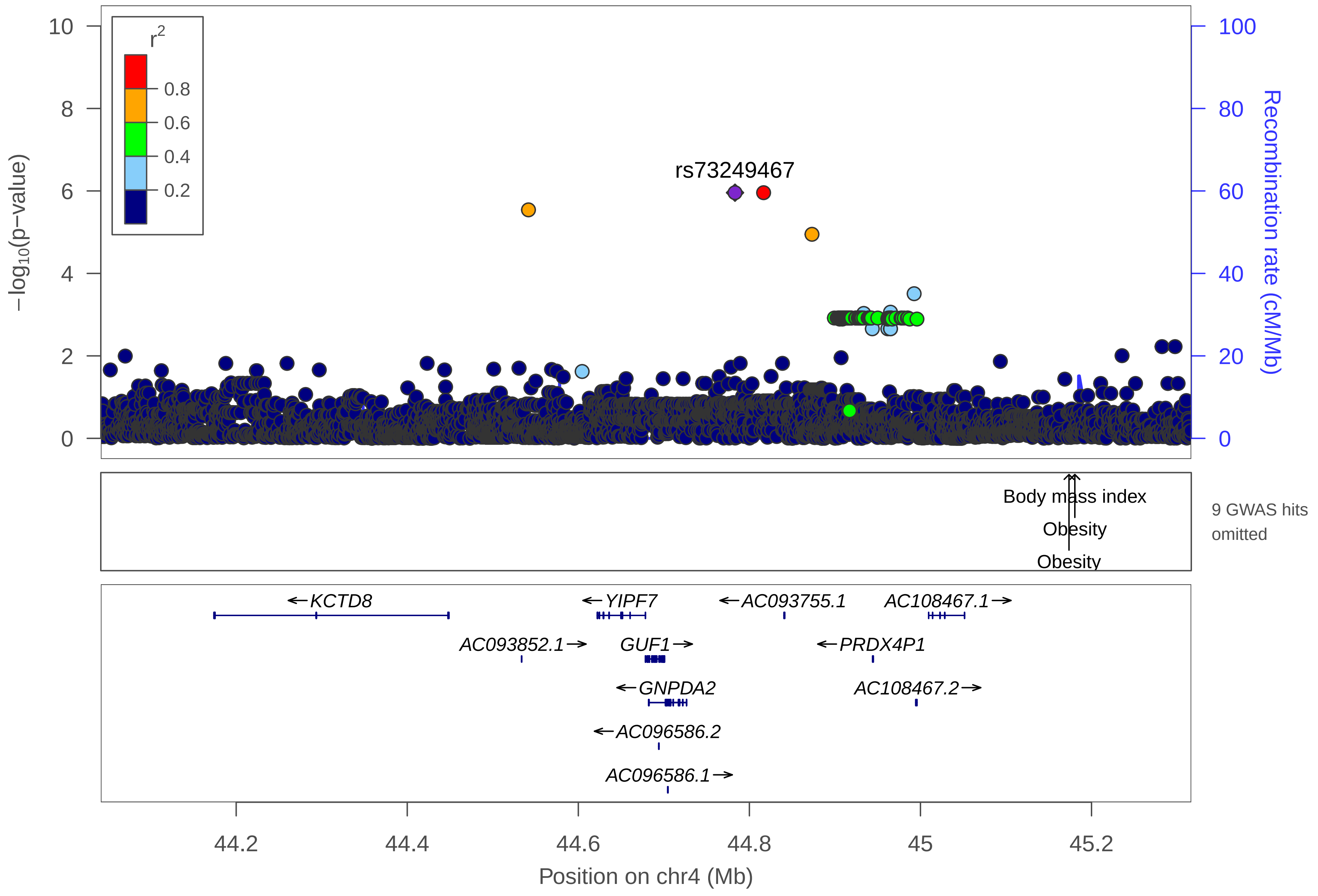

r2
0.8
0.6
0.4
0.2
rs73249467
−log10(p−value)
Recombination rate (cM/Mb)
0
2
4
6
8
10
0
20
40
60
80
100
Body mass index
Obesity
Obesity
9 GWAS hits omitted
KCTD8
YIPF7
AC093755.1
AC108467.1
AC093852.1
GUF1
PRDX4P1
GNPDA2
AC108467.2
AC096586.2
AC096586.1
44.2
44.4
44.6
44.8
45
45.2
Position on chr4 (Mb)

| | |
|---|---|
| date: | Thu Oct 20 22:13:36 2022 |
| build: | hg38 |
| display range: | chr4:44041725−45316634 [44041725−45316634] |
| hilite range: | 0 − 0 [ 0 − 0 ] |
| reference SNP: | chr4:44783196 |
| number of SNPs plotted: | 4920 |
| min P−value: | 1.1E−6 [chr4:44783196] |
| max P−value: | 10E−1 [chr4:44931134] |
| omitted GWAS Hits: | NA, NA |
| omitted GWAS Hits: | NA, NA |
| omitted GWAS Hits: | NA, NA |

Make more plots at http://csg.sph.umich.edu/locuszoom/

GWAS Catalog SNPs in Region

| chr | pos (Mb) | trait | snp |
|---|---|---|---|
| 4 | 44.21015 | Response to amphetamines | rs17641529 |
| 4 | 44.58047 | Body mass index | rs10938353 |
| 4 | 45.07699 | Systolic blood pressure | rs996004 |
| 4 | 45.17367 | Obesity | rs13130484 |
| 4 | 45.17367 | Body mass index | rs13130484 |
| 4 | 45.17367 | Childhood body mass index | rs13130484 |
| 4 | 45.17379 | Body mass index | rs16858082 |
| 4 | 45.18051 | Body mass index | rs10938397 |
| 4 | 45.18051 | Obesity | rs10938397 |
| 4 | 45.18051 | Menarche (age at onset) | rs10938397 |
| 4 | 45.18051 | Childhood body mass index | rs10938397 |
| 4 | 45.18243 | Body mass index | rs348495 |
