## Supplemental File 1 for "Genetic Associations with Age at Dementia Onset in the *PSEN1 E280A* Colombian Kindred": chr4_66743770-67753470.pdf

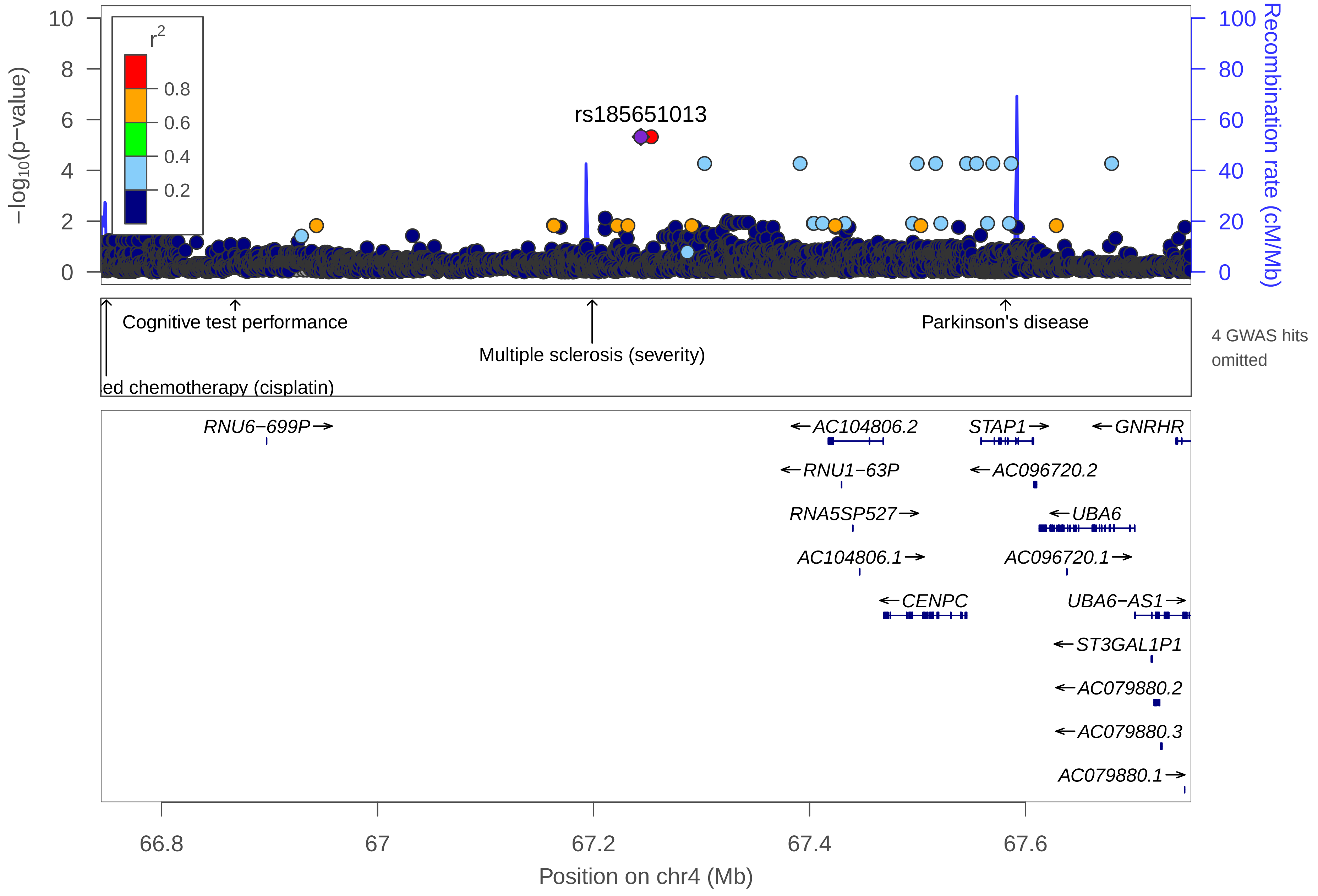

rs185651013
r2
0.8
0.6
0.4
0.2
−log10(p−value)
Recombination rate (cM/Mb)
Cognitive test performance
Multiple sclerosis (severity)
Parkinson's disease
ed chemotherapy (cisplatin)
4 GWAS hits omitted
RNU6−699P
AC104806.2
STAP1
GNRHR
RNU1−63P
AC096720.2
RNA5SP527
UBA6
AC104806.1
AC096720.1
CENPC
UBA6−AS1
ST3GAL1P1
AC079880.2
AC079880.3
AC079880.1
66.8
67
67.2
67.4
67.6
Position on chr4 (Mb)

| | |
|---|---|
| date: | Thu Oct 20 22:13:32 2022 |
| build: | hg38 |
| display range: | chr4:66743770−67753470 [66743770−67753470] |
| hilite range: | 0 − 0 [ 0 − 0 ] |
| reference SNP: | chr4:67243770 |
| number of SNPs plotted: | 3487 |
| min P−value: | 4.77E−6 [chr4:67243770] |
| max P−value: | 9.99E−1 [chr4:67170358] |
| omitted GWAS Hits: | NA, NA |
| omitted GWAS Hits: | NA |

GWAS Catalog SNPs in Region

| chr | pos (Mb) | trait | snp |
|---|---|---|---|
| 4 | 66.74885 | Response to platinum−based chemotherapy (cisplatin) | rs4860223 |
| 4 | 66.86814 | Cognitive test performance | rs1155865 |
| 4 | 66.86818 | Urate levels (BMI interaction) | rs1155866 |
| 4 | 66.95616 | Educational attainment (years of education) | rs4308415 |
| 4 | 67.19089 | Glucose homeostasis traits | rs7690543 |
| 4 | 67.19863 | Positive affect | rs17766081 |
| 4 | 67.19872 | Multiple sclerosis (severity) | rs10518025 |
| 4 | 67.58153 | Parkinson's disease | rs2242330 |
