## Supplemental File 1 for "Genetic Associations with Age at Dementia Onset in the *PSEN1 E280A* Colombian Kindred": chr4_81596651-82596651.pdf

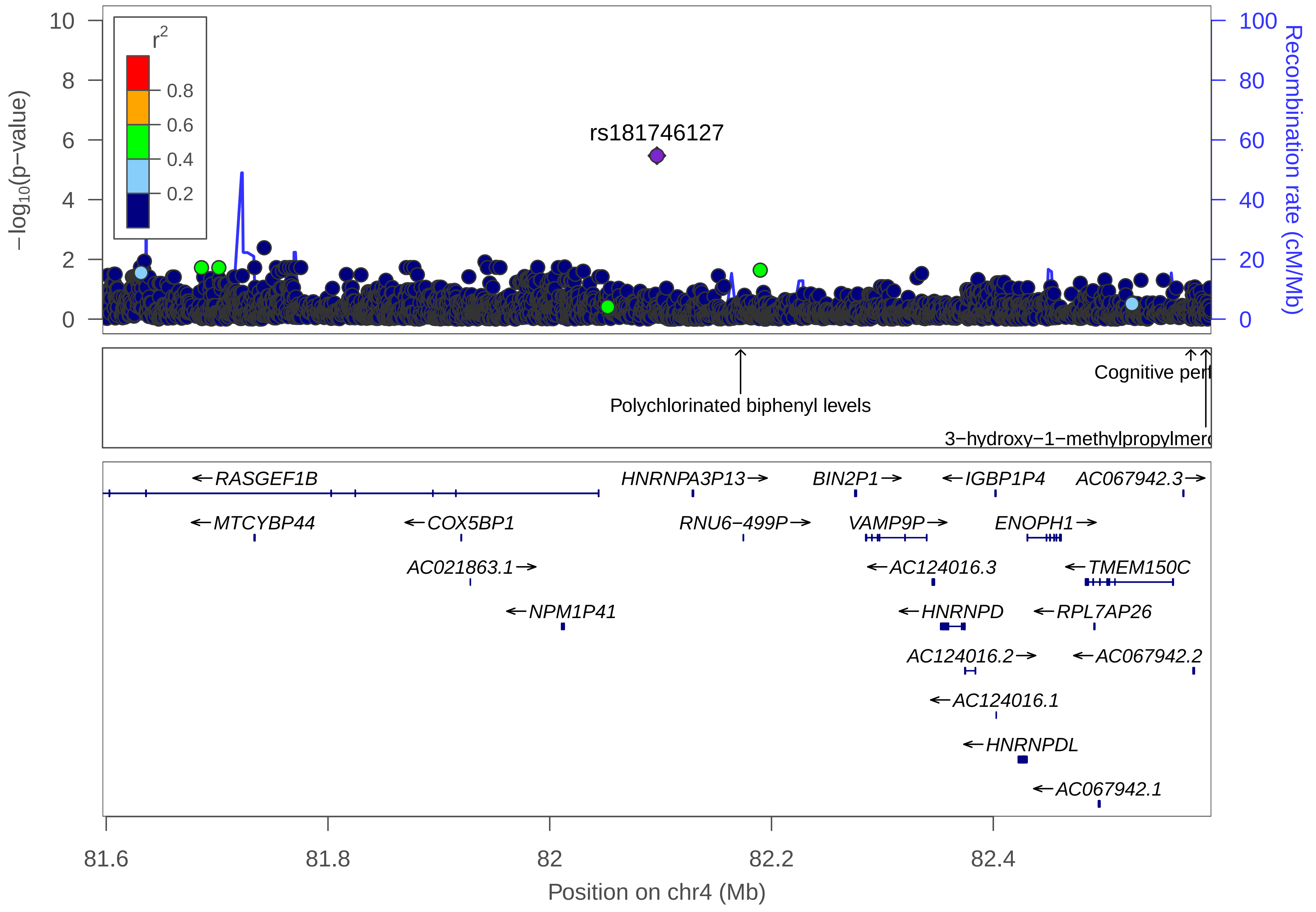

rs181746127
r2
0.8
0.6
0.4
0.2
−log10(p−value)
Recombination rate (cM/Mb)
0
2
4
6
8
10
0
20
40
60
80
100
Polychlorinated biphenyl levels
Cognitive perf
3−hydroxy−1−methylpropylmerc
RASGEF1B
HNRNPA3P13
BIN2P1
IGBP1P4
AC067942.3
MTCYBP44
COX5BP1
RNU6−499P
VAMP9P
ENOPH1
AC021863.1
AC124016.3
TMEM150C
NPM1P41
HNRNPD
RPL7AP26
AC124016.2
AC067942.2
AC124016.1
HNRNPDL
AC067942.1
81.6
81.8
82
82.2
82.4
Position on chr4 (Mb)

| | |
|---|---|
| date: | Thu Oct 20 22:13:31 2022 |
| build: | hg38 |
| display range: | chr4:81596651−82596651 [81596651−82596651] |
| hilite range: | 0 − 0 [ 0 − 0 ] |
| reference SNP: | chr4:82096651 |
| number of SNPs plotted: | 3440 |
| min P−value: | 3.38E−6 [chr4:82096651] |
| max P−value: | 10E−1 [chr4:82107786] |

Make more plots at http://csg.sph.umich.edu/locuszoom/

GWAS Catalog SNPs in Region

| chr | pos (Mb) | trait | snp |
|---|---|---|---|
| 4 | 82.17208 | Polychlorinated biphenyl levels | rs182442984 |
| 4 | 82.57816 | Cognitive performance | rs7658637 |
| 4 | 82.59169 | 3−hydroxy−1−methylpropylmercapturic acid levels in smokers | rs72909131 |
