## Supplemental File 1 for "Genetic Associations with Age at Dementia Onset in the *PSEN1 E280A* Colombian Kindred": chr5_175753006-176753006.pdf

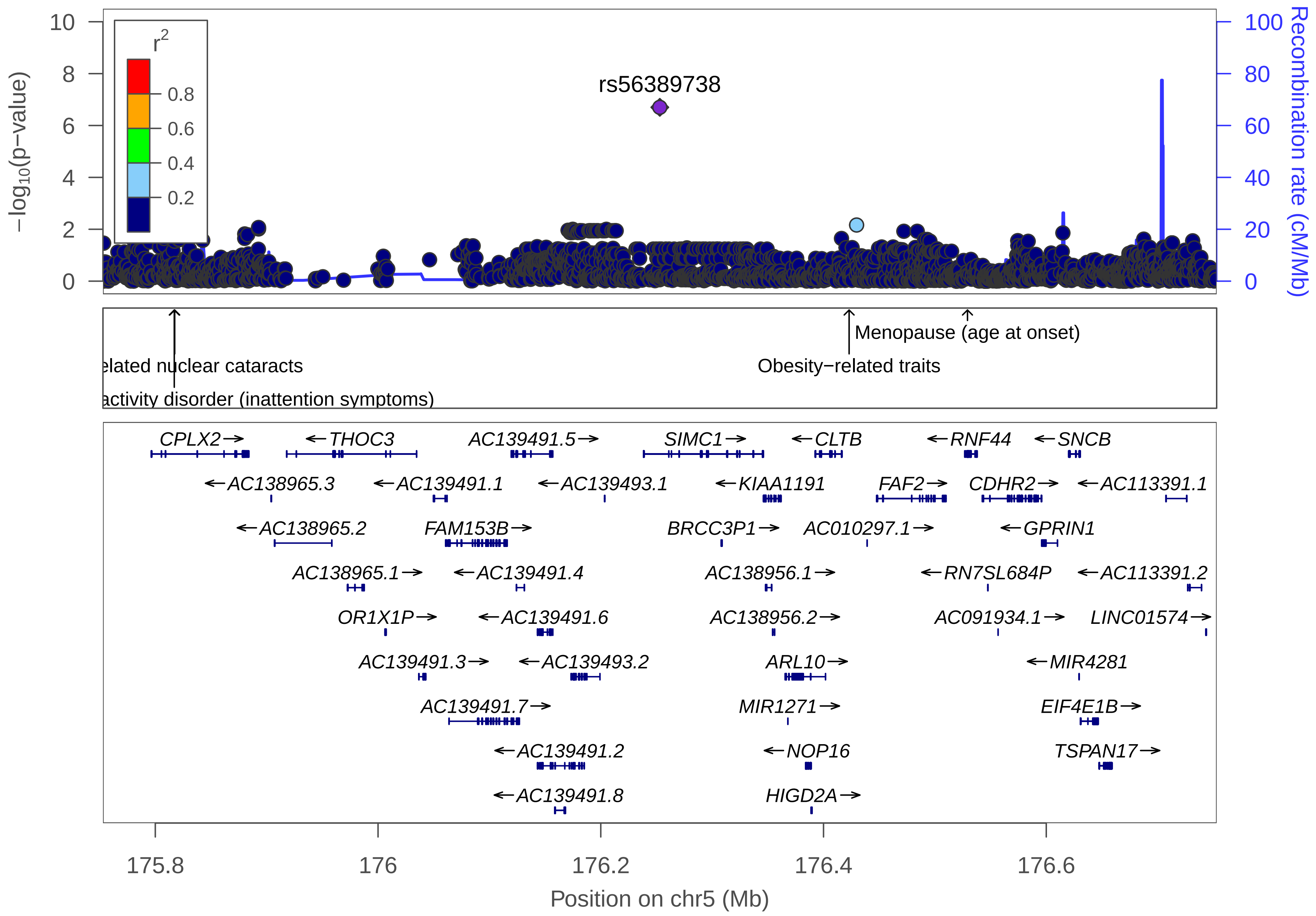

rs56389738
r2
0.8
0.6
0.4
0.2
−log10(p−value)
Recombination rate (cM/Mb)
elated nuclear cataracts
activity disorder (inattention symptoms)
Menopause (age at onset)
Obesity−related traits
CPLX2→
←THOC3
AC139491.5→
SIMC1→
←CLTB
←RNF44
←SNCB
←AC138965.3
←AC139491.1
←AC139493.1
←KIAA1191
FAF2→
CDHR2→
←AC113391.1
←AC138965.2
FAM153B→
BRCC3P1→
AC010297.1→
←GPRIN1
AC138965.1→
←AC139491.4
AC138956.1→
←RN7SL684P
←AC113391.2
OR1X1P→
←AC139491.6
AC138956.2→
AC091934.1→
LINC01574→
AC139491.3→
←AC139493.2
ARL10→
←MIR4281
AC139491.7→
MIR1271→
EIF4E1B→
←AC139491.2
←NOP16
TSPAN17→
←AC139491.8
HIGD2A→
175.8
176
176.2
176.4
176.6
Position on chr5 (Mb)

| | |
|---|---|
| date: | Thu Oct 20 22:13:33 2022 |
| build: | hg38 |
| display range: | chr5:175753006−176753006 [175753006−176753006] |
| hilite range: | 0 − 0 [ 0 − 0 ] |
| reference SNP: | chr5:176253006 |
| number of SNPs plotted: | 2346 |
| min P−value: | 1.99E−7 [chr5:176253006] |
| max P−value: | 9.99E−1 [chr5:176519943] |

Make more plots at http://csg.sph.umich.edu/locuszoom/

GWAS Catalog SNPs in Region

| chr | pos (Mb) | trait | snp |
|---|---|---|---|
| 5 | 175.8170 | Attention deficit hyperactivity disorder (inattention symptoms) | rs7448069 |
| 5 | 175.8174 | Age−related nuclear cataracts | rs55914911 |
| 5 | 176.4230 | Obesity−related traits | rs2913737 |
| 5 | 176.5293 | Menopause (age at onset) | rs890835 |
