## Supplemental File 1 for "Genetic Associations with Age at Dementia Onset in the *PSEN1 E280A* Colombian Kindred": chr6_17003320-18026280.pdf

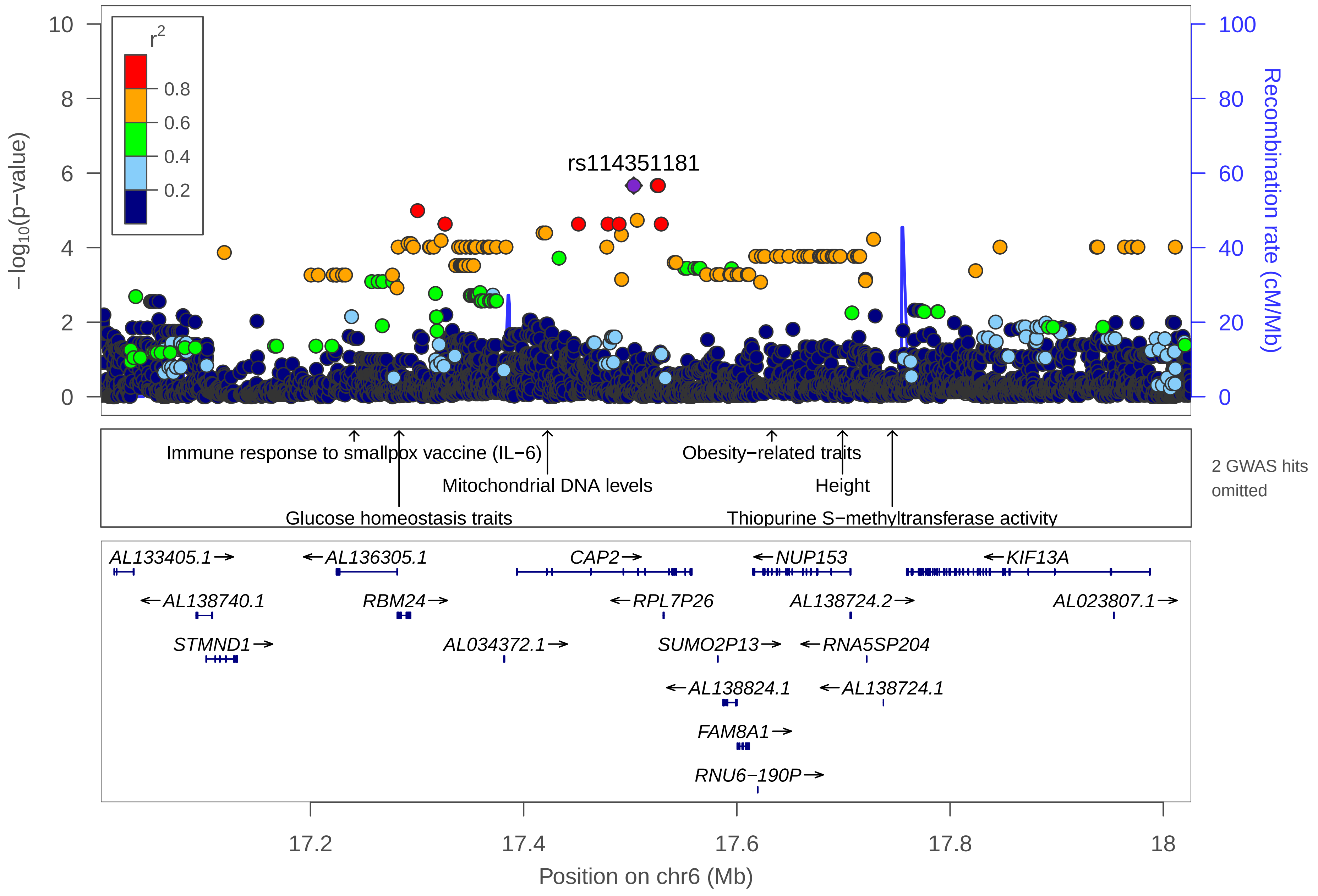

rs114351181
r2
0.8
0.6
0.4
0.2
−log10(p−value)
Recombination rate (cM/Mb)
Immune response to smallpox vaccine (IL−6)
Glucose homeostasis traits
Mitochondrial DNA levels
Obesity−related traits
Height
Thiopurine S−methyltransferase activity
2 GWAS hits omitted
AL133405.1→
←AL136305.1
CAP2→
←NUP153
←KIF13A
←AL138740.1
RBM24→
←RPL7P26
AL138724.2→
AL023807.1→
STMND1→
AL034372.1→
SUMO2P13→
←RNA5SP204
←AL138824.1
←AL138724.1
FAM8A1→
RNU6−190P→
17.2
17.4
17.6
17.8
18
Position on chr6 (Mb)

| | |
|---|---|
| date: | Thu Oct 20 22:13:34 2022 |
| build: | hg38 |
| display range: | chr6:17003320−18026280 [17003320−18026280] |
| hilite range: | 0 − 0 [ 0 − 0 ] |
| reference SNP: | chr6:17503320 |
| number of SNPs plotted: | 3761 |
| min P−value: | 2.16E−6 [chr6:17503320] |
| max P−value: | 9.99E−1 [chr6:17657409] |
| omitted GWAS Hits: | chr6:17.63283−Obesity−related traits, NA |

Make more plots at http://csg.sph.umich.edu/locuszoom/

GWAS Catalog SNPs in Region

| chr | pos (Mb) | trait | snp |
|---|---|---|---|
| 6 | 17.16089 | Response to lithium treatment in bipolar disorder | rs9396756 |
| 6 | 17.24092 | Immune response to smallpox vaccine (IL−6) | rs1322846 |
| 6 | 17.28309 | Glucose homeostasis traits | rs3749855 |
| 6 | 17.42231 | Mitochondrial DNA levels | rs10485376 |
| 6 | 17.50571 | Response to serotonin reuptake inhibitors in major depressive disorder | rs910039 |
| 6 | 17.63283 | Obesity−related traits | rs2274136 |
| 6 | 17.69909 | Height | rs12199222 |
| 6 | 17.74578 | Thiopurine S−methyltransferase activity | rs187976596 |
