## Supplemental File 1 for "Genetic Associations with Age at Dementia Onset in the *PSEN1 E280A* Colombian Kindred": chr6_22275312-24141446.pdf

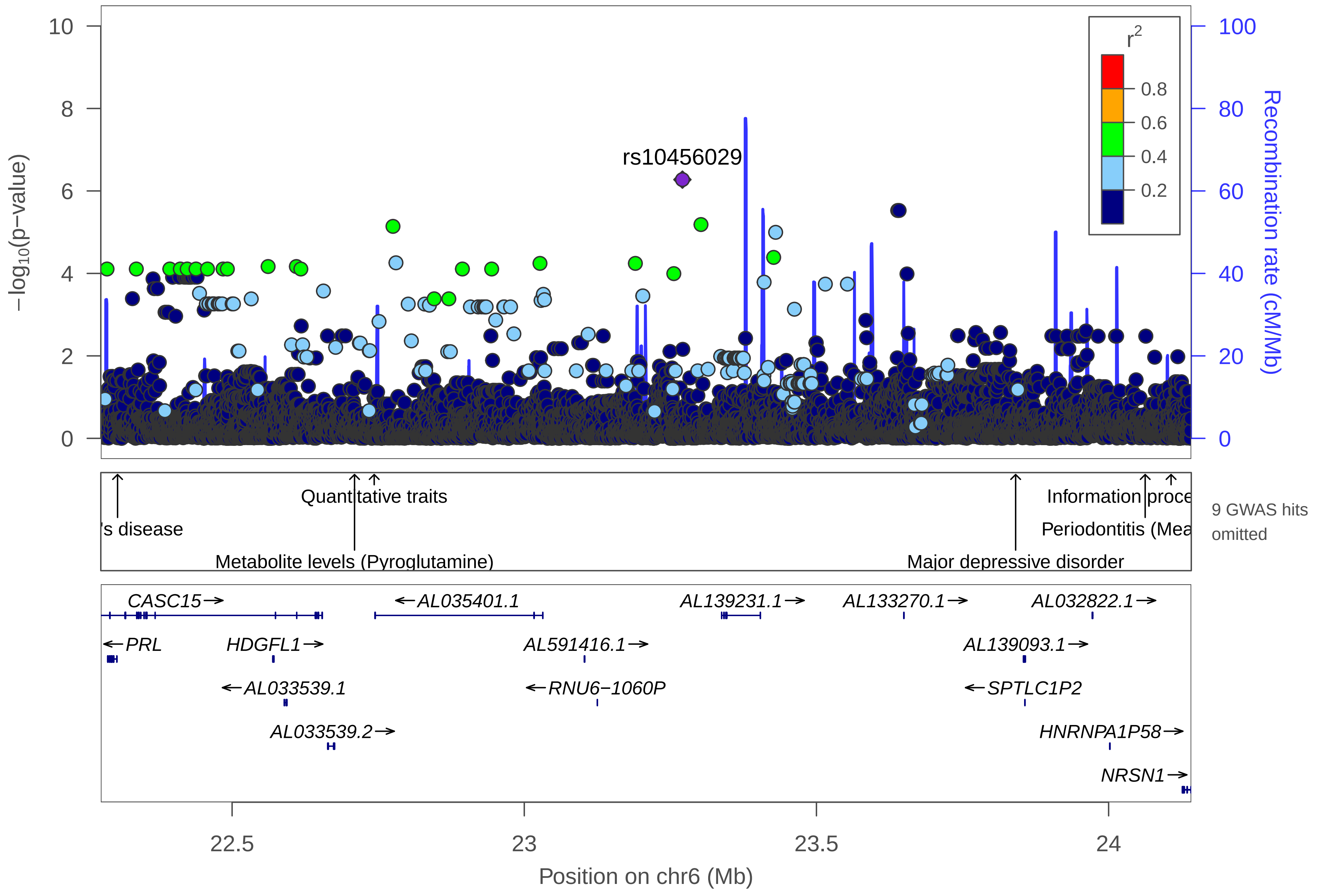

rs10456029
r²
0.8
0.6
0.4
0.2
−log10(p−value)
Recombination rate (cM/Mb)
Quantitative traits
's disease
Metabolite levels (Pyroglutamine)
Information proce
Periodontitis (Mea
Major depressive disorder
9 GWAS hits omitted
CASC15→
←AL035401.1
AL139231.1→
AL133270.1→
AL032822.1→
←PRL
HDGFL1→
AL591416.1→
AL139093.1→
←AL033539.1
←RNU6−1060P
←SPTLC1P2
AL033539.2→
HNRNPA1P58→
NRSN1→
22.5
23
23.5
24
Position on chr6 (Mb)

| | |
|---|---|
| date: | Thu Oct 20 22:13:35 2022 |
| build: | hg38 |
| display range: | chr6:22275312−24141446 [22275312−24141446] |
| hilite range: | 0 − 0 [ 0 − 0 ] |
| reference SNP: | chr6:23270633 |
| number of SNPs plotted: | 8410 |
| min P−value: | 5.31E−7 [chr6:23270633] |
| max P−value: | 9.99E−1 [chr6:23951286] |
| omitted GWAS Hits: | chr6:23.840904−Major depressive disorder, NA |
| omitted GWAS Hits: | NA, NA |
| omitted GWAS Hits: | NA, NA |

Make more plots at http://csg.sph.umich.edu/locuszoom/

GWAS Catalog SNPs in Region

| chr | pos (Mb) | trait | snp |
|---|---|---|---|
| 6 | 22.30398 | Paget's disease | rs1341239 |
| 6 | 22.70951 | Metabolite levels (Pyroglutamine) | rs4712709 |
| 6 | 22.70981 | Number of children (6+ vs. 0 or 1) | rs4712710 |
| 6 | 22.71372 | Number of children (6+ vs. 0 or 1) | rs7775534 |
| 6 | 22.74317 | Quantitative traits | rs10498712 |
| 6 | 22.75282 | Obesity−related traits | rs7356884 |
| 6 | 23.07708 | Tourette syndrome | rs9393366 |
| 6 | 23.31863 | Late−onset Alzheimer's disease | rs193129245 |
| 6 | 23.43745 | Immune reponse to smallpox (secreted IL−12p40) | rs7771911 |
| 6 | 23.84090 | Major depressive disorder | rs9466930 |
| 6 | 24.00633 | Post bronchodilator FEV1 in COPD | rs138325416 |
| 6 | 24.06256 | Periodontitis (Mean PAL) | rs146696563 |
| 6 | 24.07783 | Response to abacavir−containing treatment in HIV−1 infection (virologic failure) | rs9358749 |
| 6 | 24.07783 | Response to efavirenz−containing treatment in HIV 1 infection (virologic failure) | rs9358749 |
| 6 | 24.10686 | Information processing speed | rs6922632 |
