## Supplemental File 1 for "Genetic Associations with Age at Dementia Onset in the *PSEN1 E280A* Colombian Kindred": chr6_37607725-38607725.pdf

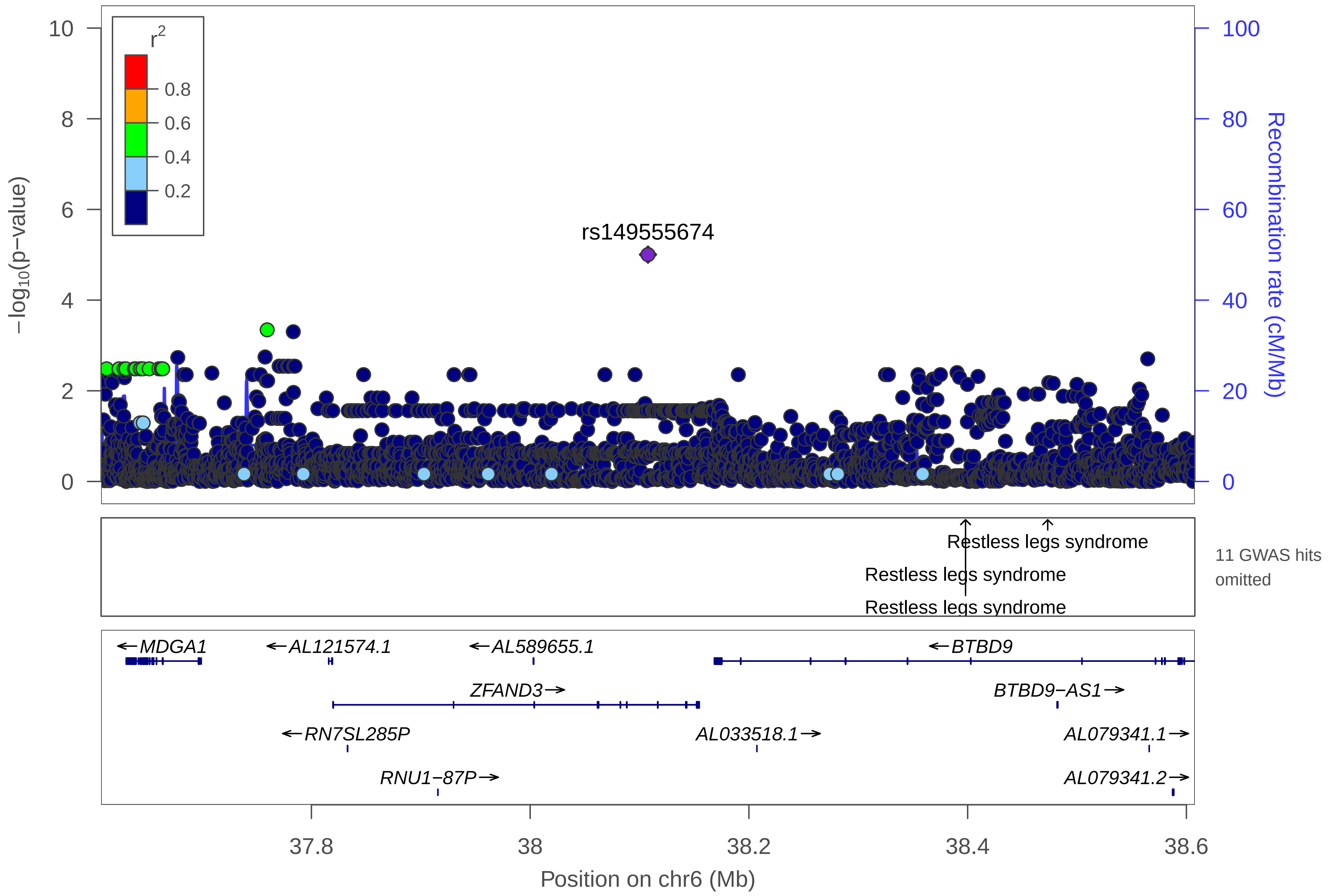

r2
0.8
0.6
0.4
0.2
rs149555674
−log10(p−value)
Recombination rate (cM/Mb)
Restless legs syndrome
Restless legs syndrome
Restless legs syndrome
11 GWAS hits omitted
MDGA1
AL121574.1
AL589655.1
BTBD9
ZFAND3
BTBD9−AS1
RN7SL285P
AL033518.1
AL079341.1
RNU1−87P
AL079341.2
37.8
38
38.2
38.4
38.6
Position on chr6 (Mb)

| | |
|---|---|
| date: | Thu Oct 20 22:13:34 2022 |
| build: | hg38 |
| display range: | chr6:37607725−38607725 [37607725−38607725] |
| hilite range: | 0 − 0 [ 0 − 0 ] |
| reference SNP: | chr6:38107725 |
| number of SNPs plotted: | 3196 |
| min P−value: | 9.91E−6 [chr6:38107725] |
| max P−value: | 10E−1 [chr6:38562524] |
| omitted GWAS Hits: | NA, NA |
| omitted GWAS Hits: | NA, NA |
| omitted GWAS Hits: | NA, NA |
| omitted GWAS Hits: | NA, NA |

Make more plots at http://csg.sph.umich.edu/locuszoom/

GWAS Catalog SNPs in Region

| chr | pos (Mb) | trait | snp |
| --- | --- | --- | --- |
| 6 | 37.67774 | Coronary artery aneurysm in Kawasaki disease | rs12210919 |
| 6 | 37.73083 | Sitting height ratio | rs6458016 |
| 6 | 38.06864 | Insulin resistance/response | rs17589516 |
| 6 | 38.13907 | Type 2 diabetes | rs9470794 |
| 6 | 38.15097 | Post bronchodilator FEV1/FVC ratio | rs190027738 |
| 6 | 38.28360 | Post bronchodilator FEV1/FVC ratio | rs77928651 |
| 6 | 38.28604 | Post bronchodilator FEV1/FVC ratio | rs139632137 |
| 6 | 38.37176 | Post bronchodilator FEV1/FVC ratio in COPD | rs150713181 |
| 6 | 38.38334 | Post bronchodilator FEV1/FVC ratio in COPD | rs114407411 |
| 6 | 38.38753 | Post bronchodilator FEV1/FVC ratio in COPD | rs192452148 |
| 6 | 38.39807 | Restless legs syndrome | rs9296249 |
| 6 | 38.39810 | Restless legs syndrome | rs9357271 |
| 6 | 38.43236 | Mammographic density | rs4565302 |
| 6 | 38.47319 | Restless legs syndrome | rs3923809 |
