## Supplemental File 1 for "Genetic Associations with Age at Dementia Onset in the *PSEN1 E280A* Colombian Kindred": chr6_60536388-61536388.pdf

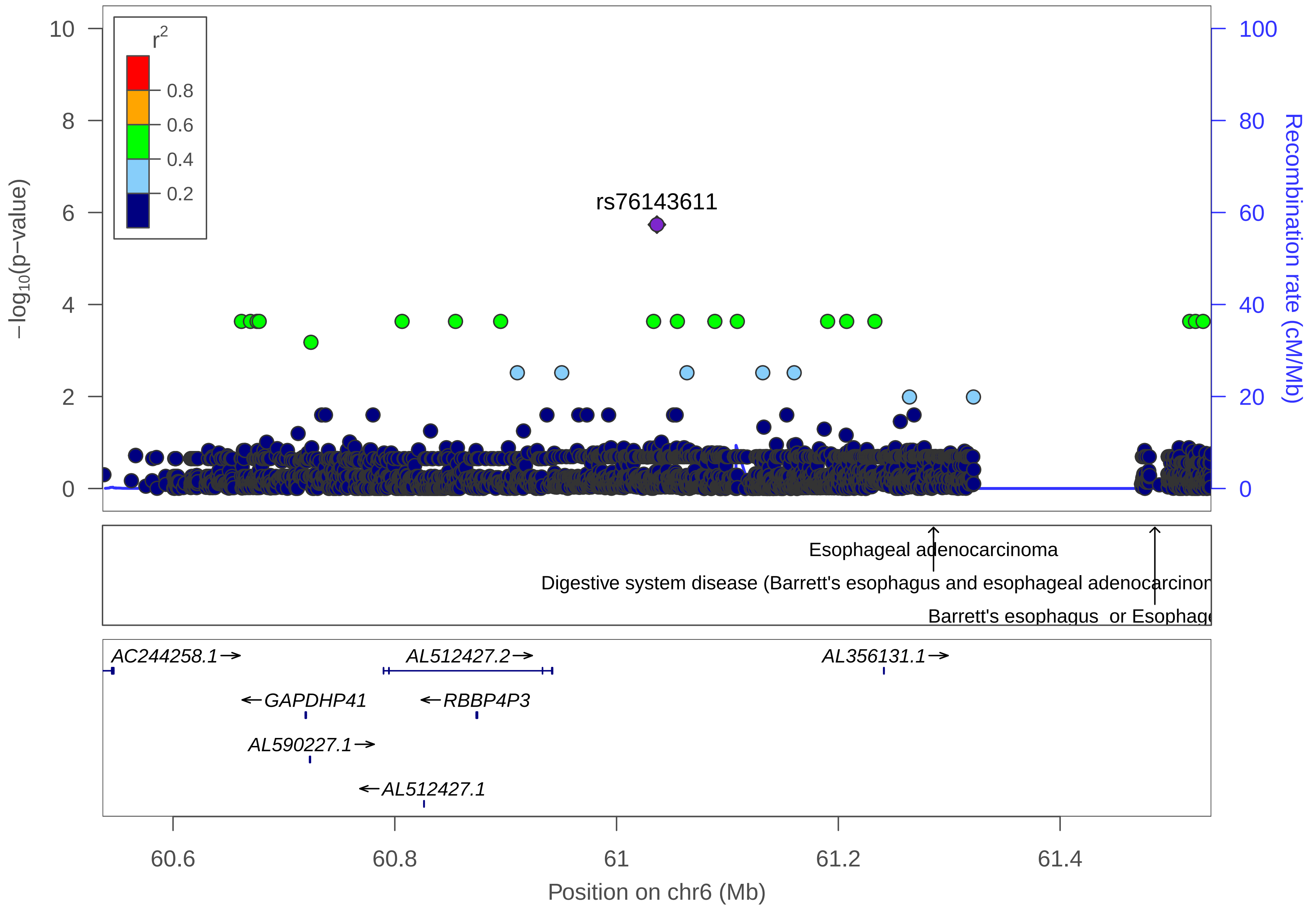

rs76143611
r2
0.8
0.6
0.4
0.2
−log10(p−value)
Recombination rate (cM/Mb)
Esophageal adenocarcinoma
Digestive system disease (Barrett's esophagus and esophageal adenocarcinom
Barrett's esophagus or Esophage
AC244258.1
GAPDHP41
AL590227.1
AL512427.2
RBBP4P3
AL512427.1
AL356131.1
60.6
60.8
61
61.2
61.4
Position on chr6 (Mb)

| | |
|---|---|
| date: | Thu Oct 20 22:13:33 2022 |
| build: | hg38 |
| display range: | chr6:60536388−61536388 [60536388−61536388] |
| hilite range: | 0 − 0 [ 0 − 0 ] |
| reference SNP: | chr6:61036388 |
| number of SNPs plotted: | 3033 |
| min P−value: | 1.84E−6 [chr6:61036388] |
| max P−value: | 10E−1 [chr6:61078936] |

Make more plots at http://csg.sph.umich.edu/locuszoom/

GWAS Catalog SNPs in Region

| chr | pos (Mb) | trait | snp |
|---|---|---|---|
| 6 | 61.28585 | Esophageal adenocarcinoma | rs2342002 |
| 6 | 61.28585 | Digestive system disease (Barrett's esophagus and esophageal adenocarcinoma combined) | rs2342002 |
| 6 | 61.48546 | Barrett's esophagus or Esophageal adenocarcinoma | rs62423175 |
