## Supplemental File 1 for "Genetic Associations with Age at Dementia Onset in the *PSEN1 E280A* Colombian Kindred": chr6_112863777-113865898.pdf

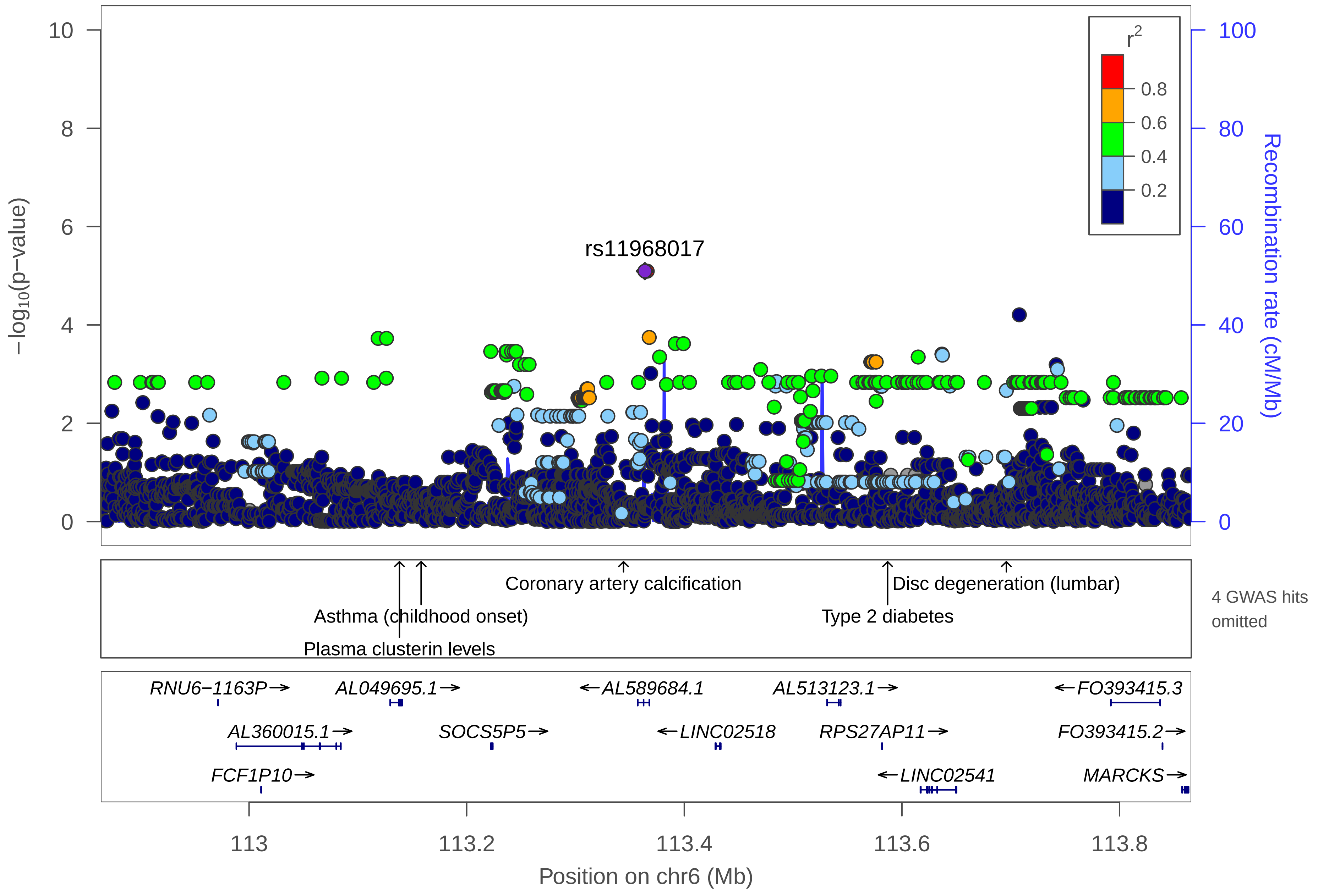

rs11968017
r2
0.8
0.6
0.4
0.2
−log10(p−value)
Recombination rate (cM/Mb)
0
2
4
6
8
10
0
20
40
60
80
100
Coronary artery calcification
Disc degeneration (lumbar)
Asthma (childhood onset)
Type 2 diabetes
Plasma clusterin levels
4 GWAS hits omitted
RNU6−1163P→
AL049695.1→
←AL589684.1
AL513123.1→
←FO393415.3
AL360015.1→
SOCS5P5→
←LINC02518
RPS27AP11→
FO393415.2→
FCF1P10→
←LINC02541
MARCKS→
113
113.2
113.4
113.6
113.8
Position on chr6 (Mb)

| | |
|---|---|
| date: | Thu Oct 20 22:13:31 2022 |
| build: | hg38 |
| display range: | chr6:112863777−113865898 [112863777−113865898] |
| hilite range: | 0 − 0 [ 0 − 0 ] |
| reference SNP: | chr6:113363777 |
| number of SNPs plotted: | 3079 |
| min P−value: | 8.06E−6 [chr6:113363777] |
| max P−value: | 10E−1 [chr6:113388411] |
| omitted GWAS Hits: | chr6:113.586847−Type 2 diabetes, NA |
| omitted GWAS Hits: | NA |

Make more plots at http://csg.sph.umich.edu/locuszoom/

GWAS Catalog SNPs in Region

| chr | pos (Mb) | trait | snp |
|---|---|---|---|
| 6 | 113.1382 | Plasma clusterin levels | rs2502399 |
| 6 | 113.1581 | Asthma (childhood onset) | rs2473967 |
| 6 | 113.3200 | Plasma omega−6 polyunsaturated fatty acid levels (arachidonic acid) | rs9942436 |
| 6 | 113.3247 | Plasma omega−6 polyunsaturated fatty acid levels (arachidonic acid) | rs2637523 |
| 6 | 113.3441 | Coronary artery calcification | rs7765175 |
| 6 | 113.3990 | Alzheimer disease and age of onset | rs11961710 |
| 6 | 113.5868 | Type 2 diabetes | rs149358103 |
| 6 | 113.6959 | Disc degeneration (lumbar) | rs9488238 |
| 6 | 113.7505 | Event free survival in diffuse large B−cell lymphoma treated with immunochemotherapy | rs7765004 |
