## Supplemental File 1 for "Genetic Associations with Age at Dementia Onset in the *PSEN1 E280A* Colombian Kindred": chr6_119411624-121939143.pdf

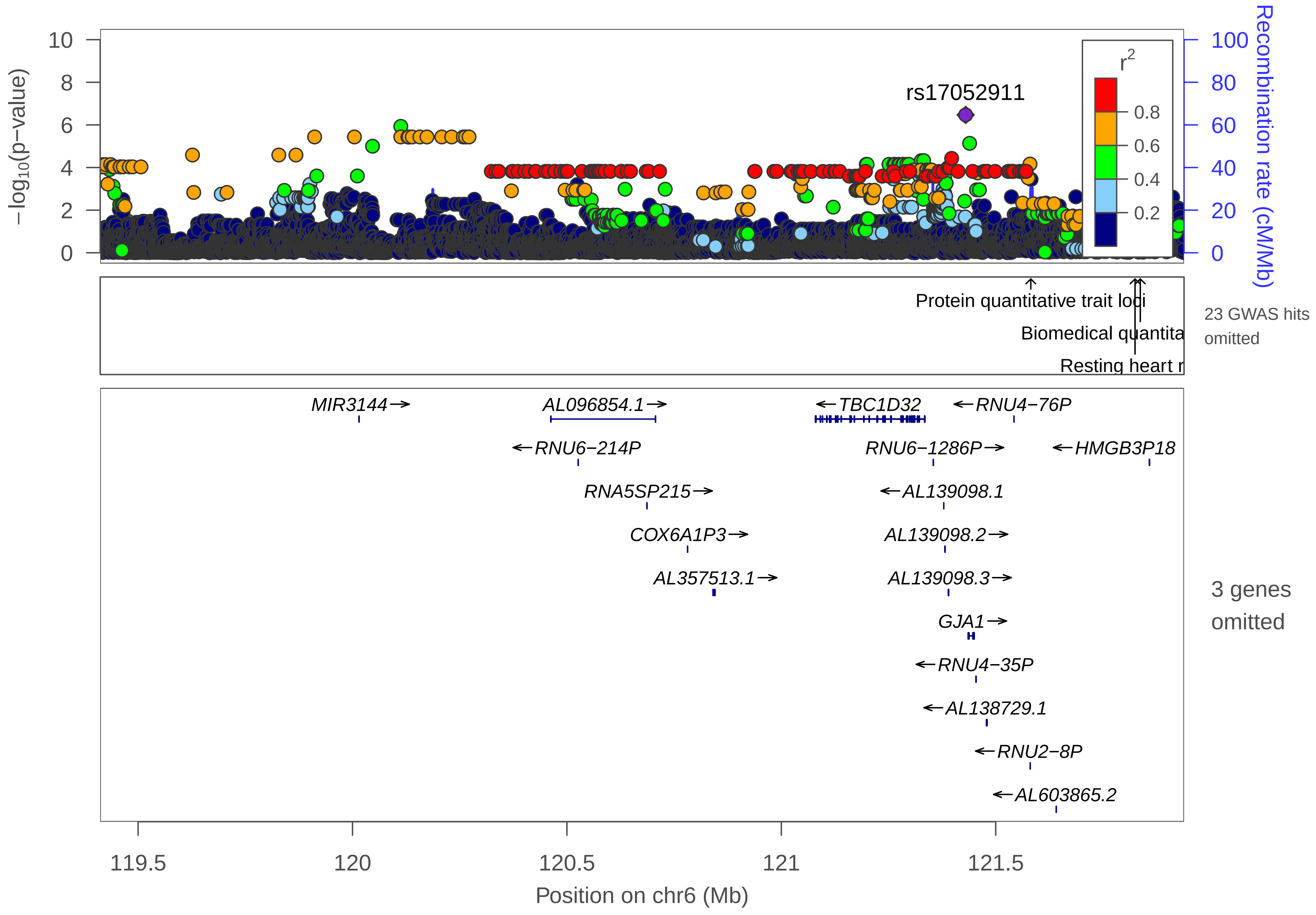
rs17052911
r²
0.8
0.6
0.4
0.2
−log10(p−value)
Recombination rate (cM/Mb)
Protein quantitative trait loci
Biomedical quantita
Resting heart r
23 GWAS hits omitted
MIR3144→
AL096854.1→
←TBC1D32
←RNU4−76P
←RNU6−214P
RNU6−1286P→
←HMGB3P18
RNA5SP215→
←AL139098.1
COX6A1P3→
AL139098.2→
AL357513.1→
AL139098.3→
GJA1→
←RNU4−35P
←AL138729.1
←RNU2−8P
←AL603865.2
3 genes omitted
119.5
120
120.5
121
121.5
Position on chr6 (Mb)

| | |
|---|---|
| date: | Thu Oct 20 22:13:36 2022 |
| build: | hg38 |
| display range: | chr6:119411624−121939143 [119411624−121939143] |
| hilite range: | 0 − 0 [ 0 − 0 ] |
| reference SNP: | chr6:121429849 |
| number of SNPs plotted: | 9023 |
| min P−value: | 3.39E−7 [chr6:121429849] |
| max P−value: | 10E−1 [chr6:121311588] |
| omitted Genes: | SLC25A5P7, RPL23AP48, AL603865.1 |
| omitted GWAS Hits: | NA, NA |
| omitted GWAS Hits: | NA, NA |
| omitted GWAS Hits: | NA, NA |
| omitted GWAS Hits: | NA, NA |
| omitted GWAS Hits: | NA, NA |
| omitted GWAS Hits: | NA, NA |
| omitted GWAS Hits: | NA, NA |
| omitted GWAS Hits: | NA, NA |

Make more plots at http://csg.sph.umich.edu/locuszoom/

GWAS Catalog SNPs in Region

| chr | pos (Mb) | trait | snp |
|---|---|---|---|
| 6 | 119.5450 | Immune reponse to smallpox (secreted IL−2) | rs1392089 |
| 6 | 119.6222 | Spontaneous preterm birth (preterm birth) | rs2794256 |
| 6 | 119.8645 | Body mass index | rs9374842 |
| 6 | 119.9464 | Neuroticism | rs6569095 |
| 6 | 119.9536 | Systemic lupus erythematosus | rs4574684 |
| 6 | 120.1544 | Post bronchodilator FEV1/FVC ratio | rs56804048 |
| 6 | 120.1760 | Post bronchodilator FEV1/FVC ratio | rs56392554 |
| 6 | 120.4428 | Post bronchodilator FEV1 in COPD | rs73527062 |
| 6 | 120.7773 | Primary tooth development (number of teeth) | rs2817937 |
| 6 | 120.7790 | HIV−1 viral setpoint | rs2789066 |
| 6 | 120.8687 | Peak creatinine levels in vancomycin therapy | rs2789047 |
| 6 | 121.0180 | Cognitive performance | rs1343075 |
| 6 | 121.4096 | Heart rate | rs17083533 |
| 6 | 121.4274 | Resting heart rate | rs11154022 |
| 6 | 121.4374 | Resting heart rate | rs3792943 |
| 6 | 121.4602 | Heart rate | rs11154027 |
| 6 | 121.4957 | Migraine | rs9490306 |
| 6 | 121.4957 | Migraine without aura | rs9490306 |
| 6 | 121.5249 | Migraine | rs28455731 |
| 6 | 121.5817 | Protein quantitative trait loci | rs4541776 |
| 6 | 121.6145 | Systolic blood pressure response to hydrochlorothiazide in hypertension | rs11750990 |

GWAS Catalog SNPs in Region

| chr | pos (Mb) | trait | snp |
|---|---|---|---|
| 6 | 121.7933 | Resting heart rate | rs9320841 |
| 6 | 121.8103 | Heart rate | rs1015451 |
| 6 | 121.8249 | Resting heart rate | rs9398652 |
| 6 | 121.8371 | Biomedical quantitative traits | rs12110693 |
| 6 | 121.8470 | Resting heart rate | rs1320761 |
