## Supplemental File 1 for "Genetic Associations with Age at Dementia Onset in the *PSEN1 E280A* Colombian Kindred": chr6_139376036-141445884.pdf

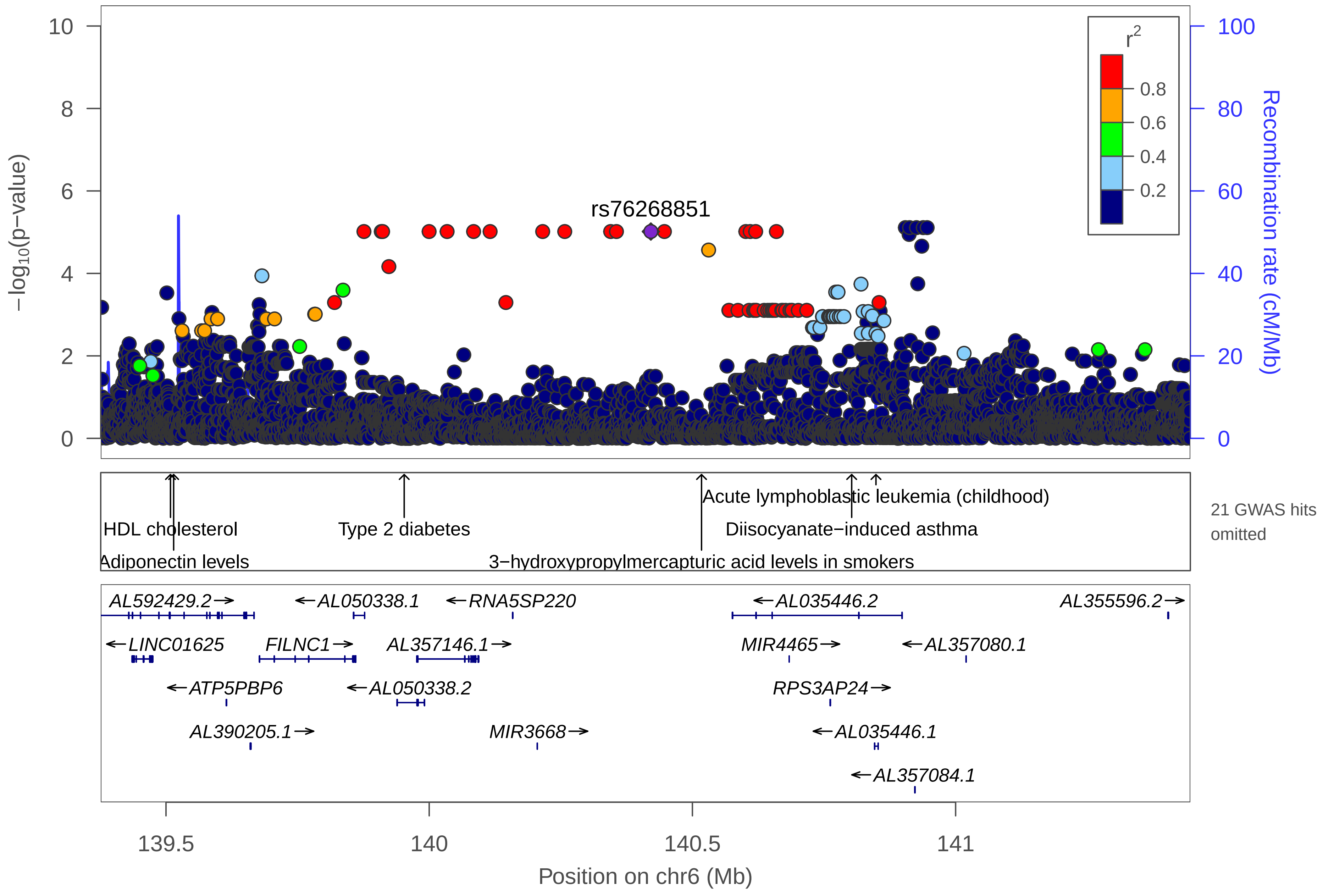
rs76268851
−log10(p−value)
Recombination rate (cM/Mb)
r2
0.8
0.6
0.4
0.2
10
8
6
4
2
0
100
80
60
40
20
0
HDL cholesterol
Adiponectin levels
Type 2 diabetes
Acute lymphoblastic leukemia (childhood)
Diisocyanate−induced asthma
3−hydroxypropylmercapturic acid levels in smokers
21 GWAS hits omitted
AL592429.2
AL050338.1
RNA5SP220
AL035446.2
AL355596.2
LINC01625
FILNC1
AL357146.1
MIR4465
AL357080.1
ATP5PBP6
AL050338.2
RPS3AP24
AL390205.1
MIR3668
AL035446.1
AL357084.1
139.5
140
140.5
141
Position on chr6 (Mb)

| | |
|---|---|
| date: | Thu Oct 20 22:13:35 2022 |
| build: | hg38 |
| display range: | chr6:139376036−141445884 [139376036−141445884] |
| hilite range: | 0 − 0 [ 0 − 0 ] |
| reference SNP: | chr6:140421061 |
| number of SNPs plotted: | 5695 |
| min P−value: | 7.76E−6 [chr6:140904514] |
| max P−value: | 10E−1 [chr6:140223977] |
| omitted GWAS Hits: | chr6:140.802574−Diisocyanate−induced asthma, chr6:140.517337−3−hydroxypropylmercapturi |
| omitted GWAS Hits: | NA, NA |
| omitted GWAS Hits: | NA, NA |
| omitted GWAS Hits: | NA, NA |
| omitted GWAS Hits: | NA, NA |
| omitted GWAS Hits: | NA, NA |
| omitted GWAS Hits: | NA, NA |

Make more plots at http://csg.sph.umich.edu/locuszoom/

GWAS Catalog SNPs in Region

| chr | pos (Mb) | trait | snp |
|---|---|---|---|
| 6 | 139.4567 | Post bronchodilator FEV1/FVC ratio | rs184091137 |
| 6 | 139.5085 | HDL cholesterol | rs605066 |
| 6 | 139.5100 | Blood trace element (Se levels) | rs679582 |
| 6 | 139.5129 | Mean corpuscular hemoglobin | rs632057 |
| 6 | 139.5129 | Mean corpuscular volume | rs632057 |
| 6 | 139.5144 | Triglycerides | rs17585887 |
| 6 | 139.5146 | Adiponectin levels | rs668459 |
| 6 | 139.5146 | Mean corpuscular volume | rs668459 |
| 6 | 139.5146 | Mean corpuscular hemoglobin | rs668459 |
| 6 | 139.5173 | Mean corpuscular hemoglobin | rs628751 |
| 6 | 139.5183 | Mean corpuscular volume | rs643381 |
| 6 | 139.5184 | Triglycerides | rs608736 |
| 6 | 139.5196 | Adiponectin levels | rs592423 |
| 6 | 139.5233 | Red blood cell traits | rs590856 |
| 6 | 139.5777 | Urate levels in lean individuals | rs9495528 |
| 6 | 139.8187 | IgG glycosylation | rs12664111 |
| 6 | 139.8323 | Response to simvastatin treatment (PCSK9 protein level change) | rs6903961 |
| 6 | 139.8749 | Alzheimer disease and age of onset | rs17069431 |
| 6 | 139.9525 | Type 2 diabetes | rs642858 |
| 6 | 140.1277 | 3−hydroxypropylmercapturic acid levels in smokers | rs75402058 |
| 6 | 140.1958 | 3−hydroxypropylmercapturic acid levels in smokers | rs76846186 |

GWAS Catalog SNPs in Region

| chr | pos (Mb) | trait | snp |
|---|---|---|---|
| 6 | 140.5173 | 3−hydroxypropylmercapturic acid levels in smokers | rs146383502 |
| 6 | 140.7007 | Night sleep phenotypes | rs6900992 |
| 6 | 140.7014 | Night sleep phenotypes | rs7740706 |
| 6 | 140.7368 | Systolic blood pressure response to hydrochlorothiazide in hypertension | rs2876449 |
| 6 | 140.8026 | Diisocyanate−induced asthma | rs61613191 |
| 6 | 140.8487 | Acute lymphoblastic leukemia (childhood) | rs11155133 |
