## Supplemental File 1 for "Genetic Associations with Age at Dementia Onset in the *PSEN1 E280A* Colombian Kindred": chr6_146313216-148663893.pdf

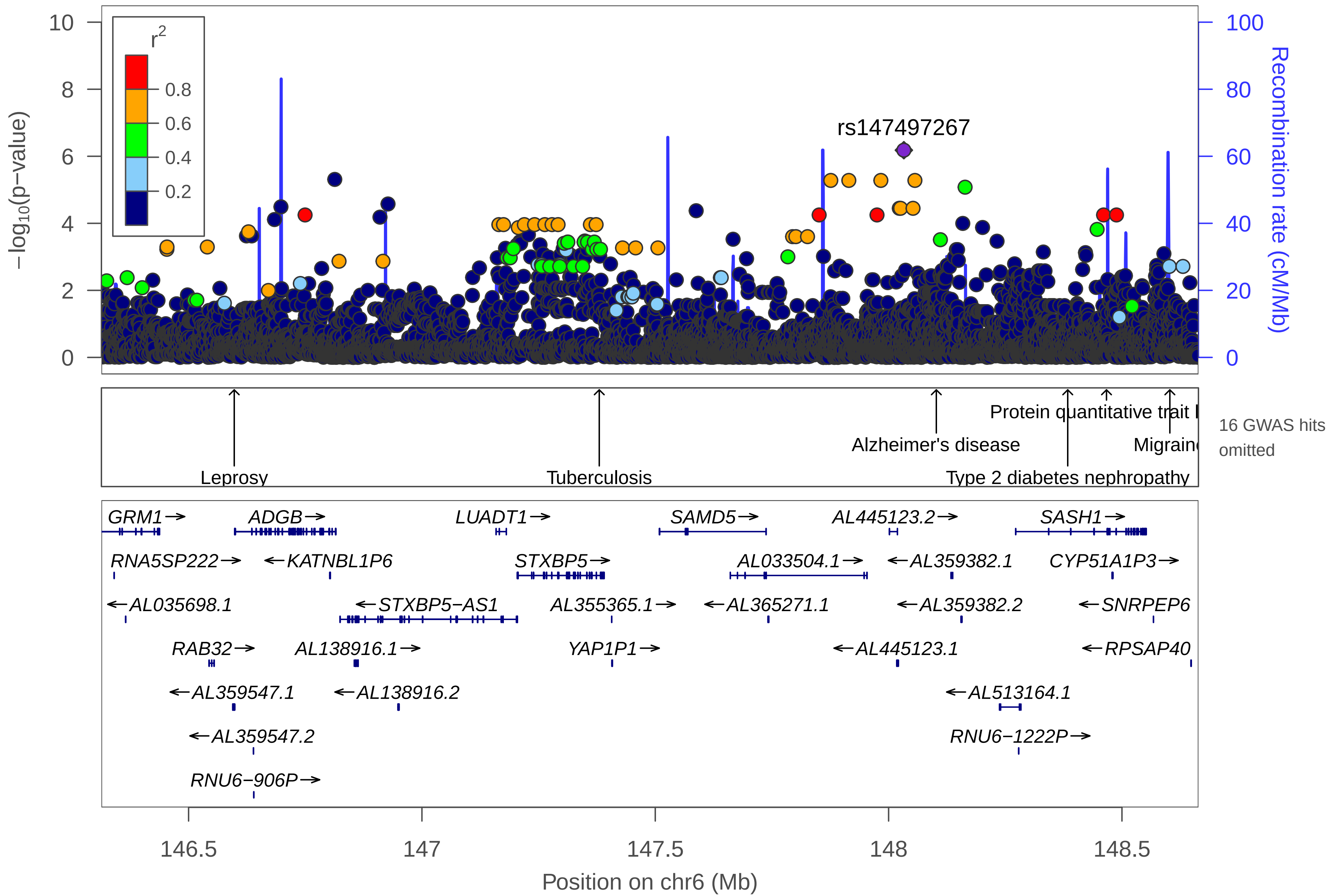
r2
0.8
0.6
0.4
0.2
−log10(p−value)
Recombination rate (cM/Mb)
rs147497267
Leprosy
Tuberculosis
Alzheimer's disease
Protein quantitative trait l
Migrain
Type 2 diabetes nephropathy
16 GWAS hits omitted
GRM1→
ADGB→
LUADT1→
SAMD5→
AL445123.2→
SASH1→
RNA5SP222→
←KATNBL1P6
STXBP5→
AL033504.1→
←AL359382.1
CYP51A1P3→
←AL035698.1
←STXBP5−AS1
AL355365.1→
←AL365271.1
←AL359382.2
←SNRPEP6
RAB32→
AL138916.1→
YAP1P1→
←AL445123.1
←RPSAP40
←AL359547.1
←AL138916.2
←AL513164.1
←AL359547.2
RNU6−1222P→
RNU6−906P→
146.5
147
147.5
148
148.5
Position on chr6 (Mb)

| | |
|---|---|
| date: | Thu Oct 20 22:13:36 2022 |
| build: | hg38 |
| display range: | chr6:146313216−148663893 [146313216−148663893] |
| hilite range: | 0 − 0 [ 0 − 0 ] |
| reference SNP: | chr6:148032677 |
| number of SNPs plotted: | 8154 |
| min P−value: | 6.58E−7 [chr6:148032677] |
| max P−value: | 9.99E−1 [chr6:147745470] |
| omitted GWAS Hits: | chr6:148.602653−Migraine, NA |
| omitted GWAS Hits: | NA, NA |
| omitted GWAS Hits: | NA, NA |
| omitted GWAS Hits: | NA, NA |
| omitted GWAS Hits: | NA, NA |
| omitted GWAS Hits: | NA |

Make more plots at http://csg.sph.umich.edu/locuszoom/

GWAS Catalog SNPs in Region

| chr | pos (Mb) | trait | snp |
|---|---|---|---|
| 6 | 146.3600 | Cerebral amyloid deposition (PET imaging) | rs362902 |
| 6 | 146.5765 | Leprosy | rs13220141 |
| 6 | 146.5978 | Leprosy | rs2275606 |
| 6 | 147.2099 | Bronchopulmonary dysplasia | rs2786189 |
| 6 | 147.2142 | Low vWF levels | rs1221638 |
| 6 | 147.2282 | Bronchopulmonary dysplasia | rs556493 |
| 6 | 147.3592 | vWF levels | rs9390459 |
| 6 | 147.3800 | Tuberculosis | rs9373523 |
| 6 | 147.3822 | Plasma plasminogen activator levels | rs9399599 |
| 6 | 147.4384 | IgG glycosylation | rs9403856 |
| 6 | 147.6138 | Obesity−related traits | rs9377063 |
| 6 | 147.6504 | Myopia (pathological) | rs1302019 |
| 6 | 147.6719 | Asthma | rs9390491 |
| 6 | 148.0693 | Night sleep phenotypes | rs112390069 |
| 6 | 148.1022 | Alzheimer's disease | rs9390537 |
| 6 | 148.2002 | Thyroid hormone levels | rs9497965 |
| 6 | 148.2267 | HIV−1 control | rs9497975 |
| 6 | 148.3838 | Type 2 diabetes nephropathy | rs6930576 |
| 6 | 148.4660 | Rotator cuff tears | rs12527089 |
| 6 | 148.4669 | Protein quantitative trait loci | rs6930337 |
| 6 | 148.5206 | Smoking quantity | rs1147857 |

### GWAS Catalog SNPs in Region

| chr | pos (Mb) | trait | snp |
|---|---|---|---|
| 6 | 148.6027 | Migraine | rs13196614 |
