## Supplemental File 1 for "Genetic Associations with Age at Dementia Onset in the *PSEN1 E280A* Colombian Kindred": chr7_42808424-43818207.pdf

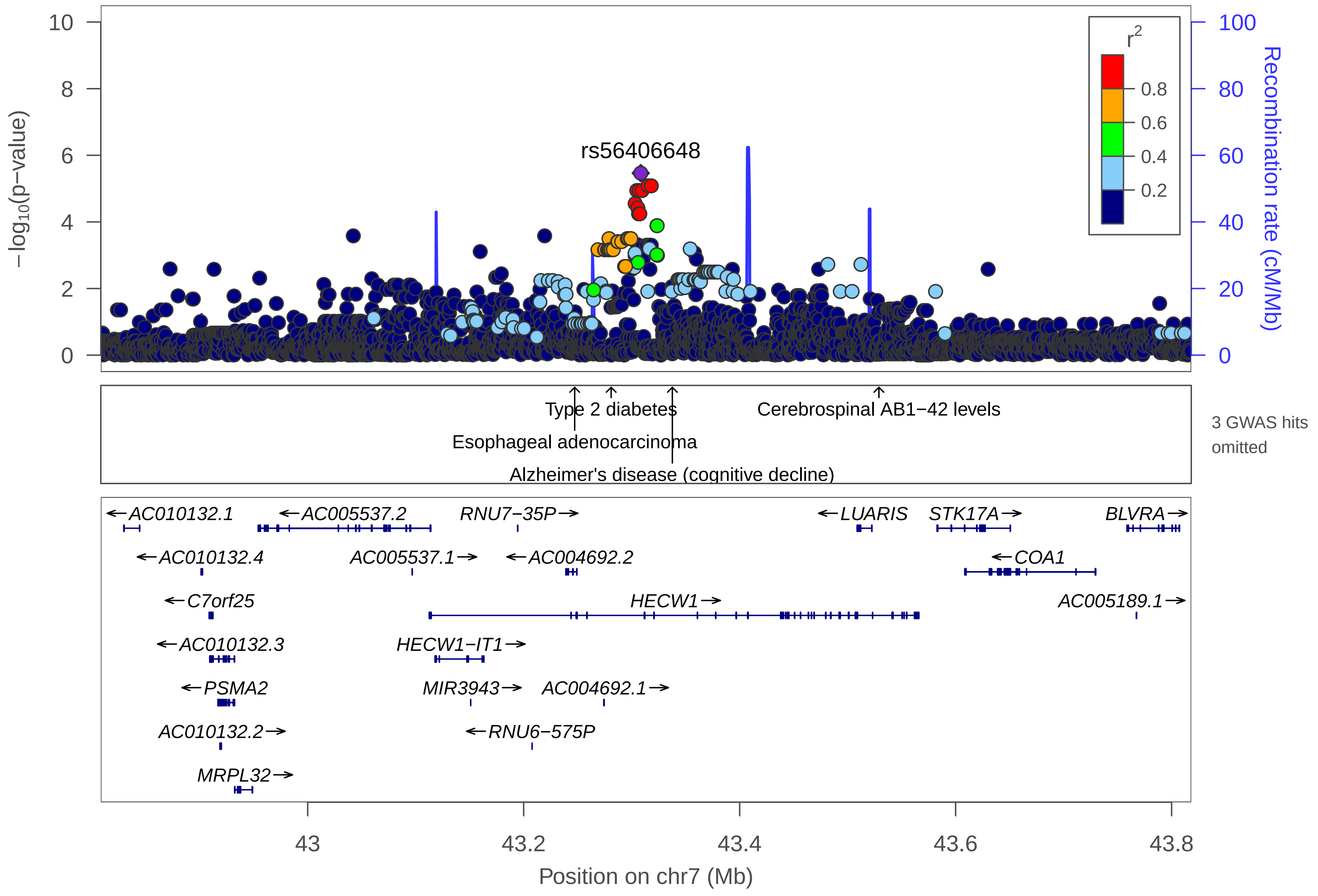

rs56406648
−log10(p−value)
Recombination rate (cM/Mb)
r2
0.8
0.6
0.4
0.2
Type 2 diabetes
Esophageal adenocarcinoma
Alzheimer's disease (cognitive decline)
Cerebrospinal AB1−42 levels
3 GWAS hits omitted
AC010132.1
AC005537.2
RNU7−35P
LUARIS
STK17A
BLVRA
AC010132.4
AC005537.1
AC004692.2
COA1
C7orf25
HECW1
AC005189.1
AC010132.3
HECW1−IT1
PSMA2
MIR3943
AC004692.1
AC010132.2
RNU6−575P
MRPL32
43
43.2
43.4
43.6
43.8
Position on chr7 (Mb)

| | |
|---|---|
| date: | Thu Oct 20 22:13:35 2022 |
| build: | hg38 |
| display range: | chr7:42808424−43818207 [42808424−43818207] |
| hilite range: | 0 − 0 [ 0 − 0 ] |
| reference SNP: | chr7:43308424 |
| number of SNPs plotted: | 3546 |
| min P−value: | 3.43E−6 [chr7:43308424] |
| max P−value: | 9.99E−1 [chr7:43371610] |
| omitted GWAS Hits: | NA, NA |

Make more plots at http://csg.sph.umich.edu/locuszoom/

GWAS Catalog SNPs in Region

| chr | pos (Mb) | trait | snp |
|---|---|---|---|
| 7 | 43.11888 | Obesity−related traits | rs2024125 |
| 7 | 43.18704 | 3−hydroxy−1−methylpropylmercapturic acid levels in smokers | rs73099645 |
| 7 | 43.24724 | Esophageal adenocarcinoma | rs17172185 |
| 7 | 43.28099 | Type 2 diabetes | rs10231619 |
| 7 | 43.29498 | Bone mineral density (Ward's triangle area) | rs34610745 |
| 7 | 43.33768 | Alzheimer's disease (cognitive decline) | rs17172199 |
| 7 | 43.52897 | Cerebrospinal AB1−42 levels | rs55704525 |
