## Supplemental File 1 for "Genetic Associations with Age at Dementia Onset in the *PSEN1 E280A* Colombian Kindred": chr8_21836974-30743250.pdf

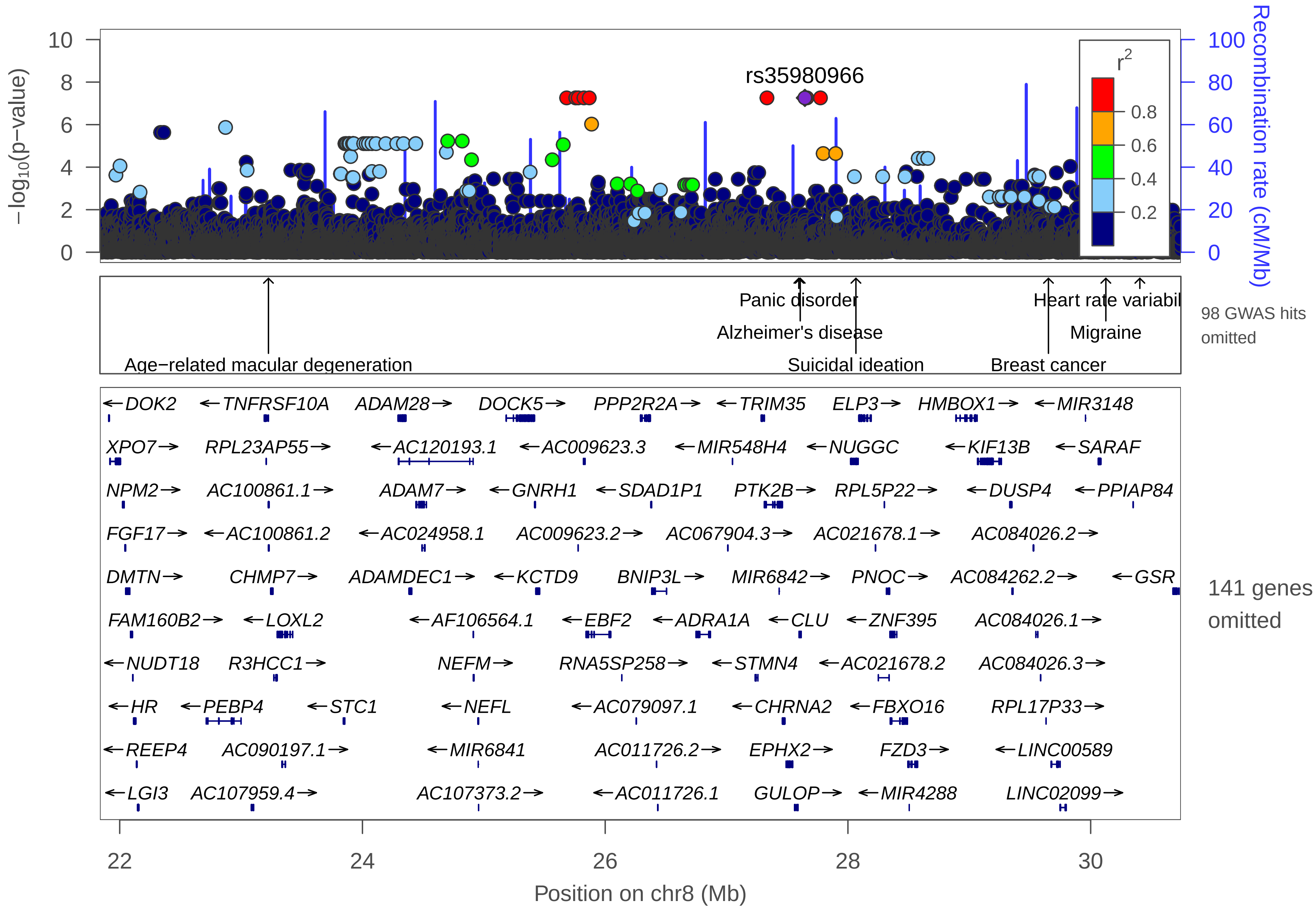

10
8
6
4
2
0
−log10(p−value)
100
80
60
40
20
0
Recombination rate (cM/Mb)
r2
0.8
0.6
0.4
0.2
rs35980966
Panic disorder
Alzheimer's disease
Suicidal ideation
Age−related macular degeneration
Heart rate variabil
Migraine
Breast cancer
98 GWAS hits omitted
DOK2
TNFRSF10A
ADAM28
DOCK5
PPP2R2A
TRIM35
ELP3
HMBOX1
MIR3148
XPO7
RPL23AP55
AC120193.1
AC009623.3
MIR548H4
NUGGC
KIF13B
SARAF
NPM2
AC100861.1
ADAM7
GNRH1
SDAD1P1
PTK2B
RPL5P22
DUSP4
PPIAP84
FGF17
AC100861.2
AC024958.1
AC009623.2
AC067904.3
AC021678.1
AC084026.2
DMTN
CHMP7
ADAMDEC1
KCTD9
BNIP3L
MIR6842
PNOC
AC084262.2
GSR
FAM160B2
LOXL2
AF106564.1
EBF2
ADRA1A
CLU
ZNF395
AC084026.1
NUDT18
R3HCC1
NEFM
RNA5SP258
STMN4
AC021678.2
AC084026.3
HR
PEBP4
STC1
NEFL
AC079097.1
CHRNA2
FBXO16
RPL17P33
REEP4
AC090197.1
MIR6841
AC011726.2
EPHX2
FZD3
LINC00589
LGI3
AC107959.4
AC107373.2
AC011726.1
GULOP
MIR4288
LINC02099
141 genes omitted
22
24
26
28
30
Position on chr8 (Mb)

| | |
|---|---|
| date: | Thu Oct 20 22:13:43 2022 |
| build: | hg38 |
| display range: | chr8:21836974−30743250 [21836974−30743250] |
| hilite range: | 0 − 0 [ 0 − 0 ] |
| reference SNP: | chr8:27645419 |
| number of SNPs plotted: | 29523 |
| min P−value: | 5.51E−8 [chr8:25682310] |
| max P−value: | 10E−1 [chr8:25282458] |
| omitted Genes: | SFTPC, BMP1, AC105206.1 |
| omitted Genes: | AC105206.3, PHYHIP, MIR320A |
| omitted Genes: | POLR3D, AC105206.2, PIWIL2 |
| omitted Genes: | SLC39A14, AC105910.1, RNU6−336P |
| omitted Genes: | PPP3CC, AC087854.1, AC087854.2 |
| omitted Genes: | SORBS3, AC037459.4, AC037459.3 |
| omitted Genes: | PDLIM2, AC037459.1, C8orf58 |
| omitted Genes: | CCAR2, AC037459.2, BIN3 |
| omitted Genes: | AC105046.1, EGR3, AC055854.1 |
| omitted Genes: | AC105046.2, AC037441.1, AC037441.2 |
| omitted Genes: | RN7SL303P, AC107959.1, RHOBTB2 |
| omitted Genes: | TNFRSF10B, AC107959.2, AC107959.3 |
| omitted Genes: | AC107959.5, TNFRSF10C, TNFRSF10D |
| omitted Genes: | TNFRSF10A−AS1, ENTPD4, AC104561.1 |
| omitted Genes: | AC104561.2, AC104561.4, AC104561.3 |

Make more plots at http://csg.sph.umich.edu/locuszoom/

GWAS Catalog SNPs in Region

| chr | pos (Mb) | trait | snp |
|---|---|---|---|
| 8 | 21.85392 | Neuropathic pain in type 2 diabetes | rs17428041 |
| 8 | 21.96330 | Mean corpuscular volume | rs7843479 |
| 8 | 22.02703 | Obesity−related traits | rs11776272 |
| 8 | 22.12086 | Obesity−related traits | rs76547188 |
| 8 | 22.22420 | Subjective well−being | rs4075536 |
| 8 | 22.23092 | Hypertriglyceridemia | rs12541335 |
| 8 | 22.40682 | Verbal declarative memory | rs7828089 |
| 8 | 22.54348 | HIV−1 viral setpoint | rs4872511 |
| 8 | 22.54733 | HIV−1 viral setpoint | rs2280890 |
| 8 | 22.60534 | Exploratory eye movement dysfunction in schizophrenia (cognitive search score) | rs11778693 |
| 8 | 22.60534 | Exploratory eye movement dysfunction in schizophrenia (responsive search score) | rs11778693 |
| 8 | 22.64432 | Exploratory eye movement dysfunction in schizophrenia (cognitive search score) | rs4242434 |
| 8 | 22.76204 | Visceral fat | rs11998649 |
| 8 | 22.78619 | Asymmetrical dimethylarginine levels | rs12155789 |
| 8 | 22.90573 | Preschool internalizing problems | rs6557600 |
| 8 | 23.08115 | Parasitemia in Tripanosoma cruzi seropositivity | rs57302454 |
| 8 | 23.10484 | Exploratory eye movement dysfunction in schizophrenia (responsive search score) | rs4077341 |
| 8 | 23.22546 | Age−related macular degeneration | rs13278062 |
| 8 | 23.22546 | Advanced age−related macular degeneration | rs79037040 |
| 8 | 23.24411 | Attention deficit hyperactivity disorder | rs7463256 |
| 8 | 23.64975 | Prostate cancer | rs142463603 |

GWAS Catalog SNPs in Region

| chr | pos (Mb) | trait | snp |
|---|---|---|---|
| 8 | 23.64987 | Post bronchodilator FEV1/FVC ratio in COPD | rs147410223 |
| 8 | 23.64987 | Post bronchodilator FEV1/FVC ratio | rs147410223 |
| 8 | 23.66895 | Prostate cancer | rs1512268 |
| 8 | 23.67650 | Prostate cancer | rs10503733 |
| 8 | 23.72671 | Treatment response for severe sepsis | rs13273073 |
| 8 | 23.74581 | Waist−to−hip ratio adjusted for body mass index | rs7830933 |
| 8 | 23.77877 | Response to mTOR inhibitor (everolimus) | rs218869 |
| 8 | 23.78598 | Preschool internalizing problems | rs310272 |
| 8 | 23.85748 | Glomerular filtration rate (creatinine) | rs3758086 |
| 8 | 23.89364 | Glomerular filtration rate in non diabetics (creatinine) | rs10109414 |
| 8 | 23.89364 | Chronic kidney disease | rs10109414 |
| 8 | 23.89364 | Glomerular filtration rate | rs10109414 |
| 8 | 23.91949 | Urate levels | rs17786744 |
| 8 | 23.96117 | Subcutaneous adipose tissue | rs7833268 |
| 8 | 24.18679 | Migraine − clinic−based | rs11777116 |
| 8 | 24.25879 | Height | rs1013209 |
| 8 | 24.35133 | Hair greying | rs7009516 |
| 8 | 24.37684 | Post bronchodilator FEV1 | rs147851892 |
| 8 | 24.37684 | Post bronchodilator FEV1/FVC ratio | rs147851892 |
| 8 | 24.52123 | White matter integrity | rs11781622 |
| 8 | 24.53775 | Post bronchodilator FEV1/FVC ratio | rs113414285 |

GWAS Catalog SNPs in Region

| chr | pos (Mb) | trait | snp |
|---|---|---|---|
| 8 | 24.54313 | Post bronchodilator FEV1/FVC ratio | rs111754512 |
| 8 | 24.67568 | Cingulate cortical amyloid beta load | rs4625043 |
| 8 | 24.87716 | Seasonality | rs196889 |
| 8 | 24.94223 | Response to anti−retroviral therapy (ddI/d4T) in HIV−1 infection (Grade 2 peripheral neuropathy) | rs6557786 |
| 8 | 25.25409 | Post bronchodilator FEV1/FVC ratio | rs187162159 |
| 8 | 25.27121 | Post bronchodilator FEV1/FVC ratio | rs182115318 |
| 8 | 25.27219 | Post bronchodilator FEV1 | rs182387400 |
| 8 | 25.27219 | Post bronchodilator FEV1/FVC ratio | rs182387400 |
| 8 | 25.27272 | Post bronchodilator FEV1 | rs147913210 |
| 8 | 25.27272 | Post bronchodilator FEV1/FVC ratio | rs147913210 |
| 8 | 25.27394 | Post bronchodilator FEV1/FVC ratio | rs150281976 |
| 8 | 25.27401 | Post bronchodilator FEV1/FVC ratio | rs190951784 |
| 8 | 25.33919 | Glucose homeostasis traits | rs13267838 |
| 8 | 25.68716 | Response to efavirenz−containing treatment in HIV 1 infection (virologic failure) | rs9314308 |
| 8 | 25.84990 | Inguinal hernia | rs6991952 |
| 8 | 26.03463 | Prostate cancer | rs11135910 |
| 8 | 26.09183 | Multiple system atrophy | rs4872401 |
| 8 | 26.34856 | Height | rs1594829 |
| 8 | 26.41167 | Schizophrenia | rs1042992 |
| 8 | 26.45458 | Cognitive decline rate in late mild cognitive impairment | rs77609452 |
| 8 | 26.85736 | Blood metabolite levels | rs13278849 |

GWAS Catalog SNPs in Region

| chr | pos (Mb) | trait | snp |
|---|---|---|---|
| 8 | 26.86136 | Response to amphetamines | rs4732957 |
| 8 | 27.03703 | Epilepsy and lamotrigine−induced maculopapular eruptions | rs7461897 |
| 8 | 27.06460 | Musician's dystonia | rs17060993 |
| 8 | 27.21908 | Bipolar disorder and schizophrenia | rs1446682 |
| 8 | 27.33760 | Alzheimer's disease (late onset) | rs28834970 |
| 8 | 27.37004 | Crohn's disease | rs17057051 |
| 8 | 27.37004 | Inflammatory bowel disease | rs17057051 |
| 8 | 27.42717 | Neuroticism | rs3779635 |
| 8 | 27.45860 | Alzheimer's disease in APOE e4− carriers | rs2271920 |
| 8 | 27.47050 | Schizophrenia | rs2565065 |
| 8 | 27.48027 | Response to zileuton treatment in asthma (FEV1 change interaction) | rs2741335 |
| 8 | 27.55928 | Dental caries | rs17057381 |
| 8 | 27.58461 | Schizophrenia | rs73229090 |
| 8 | 27.59533 | Panic disorder | rs17466684 |
| 8 | 27.59874 | Alzheimer's disease in APOE e4− carriers | rs2279590 |
| 8 | 27.60700 | Alzheimer's disease | rs11136000 |
| 8 | 27.60880 | Alzheimer's disease (late onset) | rs1532278 |
| 8 | 27.61017 | Alzheimer's disease (late onset) | rs9331896 |
| 8 | 27.61017 | Alzheimer's disease in APOE e4+ carriers | rs9331896 |
| 8 | 27.63027 | Alzheimer's disease | rs569214 |
| 8 | 27.69901 | Plasma clusterin levels | rs4545046 |

GWAS Catalog SNPs in Region

| chr | pos (Mb) | trait | snp |
|---|---|---|---|
| 8 | 27.88290 | Adverse response to chemotherapy (neutropenia/leucopenia) (cisplatin) | rs11774576 |
| 8 | 27.94608 | Low vWF levels | rs4276643 |
| 8 | 28.04837 | Cardiac Troponin−T levels | rs6983473 |
| 8 | 28.06587 | Suicidal ideation | rs4732812 |
| 8 | 28.20446 | Childhood body mass index | rs13253111 |
| 8 | 28.32424 | IgG glycosylation | rs11779594 |
| 8 | 28.74727 | Height | rs10448080 |
| 8 | 28.93364 | Obesity−related traits | rs2221894 |
| 8 | 29.09737 | Metabolite levels (X−11787) | rs7844537 |
| 8 | 29.13751 | Metabolite levels (HVA−5−HIAA Factor score) | rs13251954 |
| 8 | 29.22477 | PR interval in Tripanosoma cruzi seropositivity | rs75609241 |
| 8 | 29.24854 | Metabolite levels (HVA) | rs2954793 |
| 8 | 29.29626 | Corticobasal degeneration | rs643472 |
| 8 | 29.48694 | Colorectal cancer | rs12548021 |
| 8 | 29.50279 | Response to statin therapy | rs10091038 |
| 8 | 29.65210 | Breast cancer | rs9693444 |
| 8 | 29.85874 | Diisocyanate−induced asthma | rs16876083 |
| 8 | 30.12500 | Migraine | rs12681963 |
| 8 | 30.40527 | Heart rate variability traits | rs2979481 |
| 8 | 30.57934 | Metabolite levels (X−11787) | rs10503871 |
| 8 | 30.64134 | Cognitive performance | rs2978263 |
