## Supplemental File 1 for "Genetic Associations with Age at Dementia Onset in the *PSEN1 E280A* Colombian Kindred": chr9_69338597-70361150.pdf

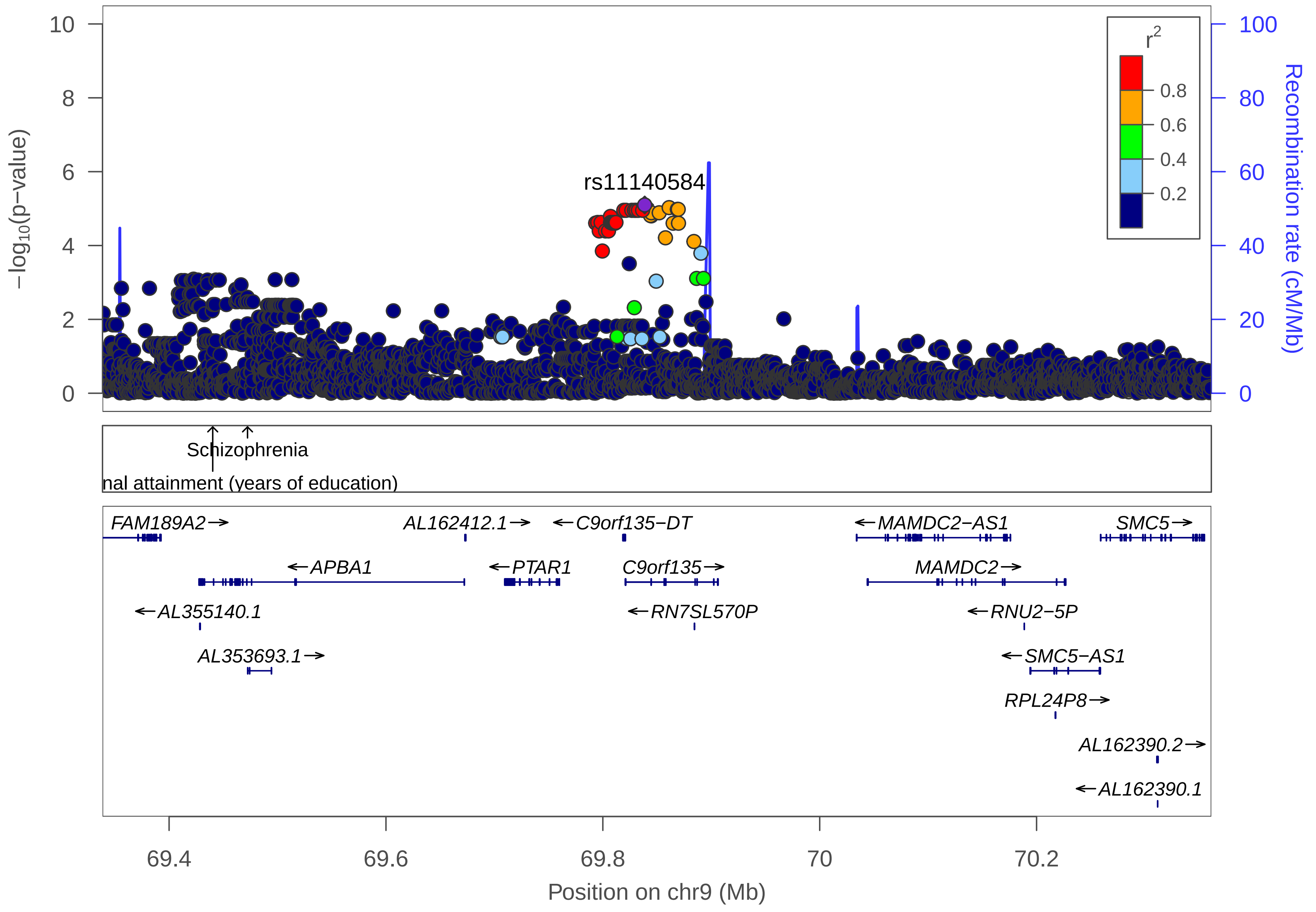

rs11140584
r2
0.8
0.6
0.4
0.2
−log10(p−value)
Recombination rate (cM/Mb)
Schizophrenia
nal attainment (years of education)
FAM189A2
AL162412.1
C9orf135−DT
MAMDC2−AS1
SMC5
APBA1
PTAR1
C9orf135
MAMDC2
AL355140.1
RN7SL570P
RNU2−5P
AL353693.1
SMC5−AS1
RPL24P8
AL162390.2
AL162390.1
69.4
69.6
69.8
70
70.2
Position on chr9 (Mb)

| | |
|---|---|
| date: | Thu Oct 20 22:13:35 2022 |
| build: | hg38 |
| display range: | chr9:69338597−70361150 [69338597−70361150] |
| hilite range: | 0 − 0 [ 0 − 0 ] |
| reference SNP: | chr9:69838597 |
| number of SNPs plotted: | 3077 |
| min P−value: | 7.96E−6 [chr9:69838597] |
| max P−value: | 9.99E−1 [chr9:69679829] |

Make more plots at http://csg.sph.umich.edu/locuszoom/

GWAS Catalog SNPs in Region

| chr | pos (Mb) | trait | snp |
|---|---|---|---|
| 9 | 69.44024 | Educational attainment (years of education) | rs7033137 |
| 9 | 69.47232 | Schizophrenia | rs4744901 |
