## Supplemental File 1 for "Genetic Associations with Age at Dementia Onset in the *PSEN1 E280A* Colombian Kindred": chr10_61630-1080236.pdf

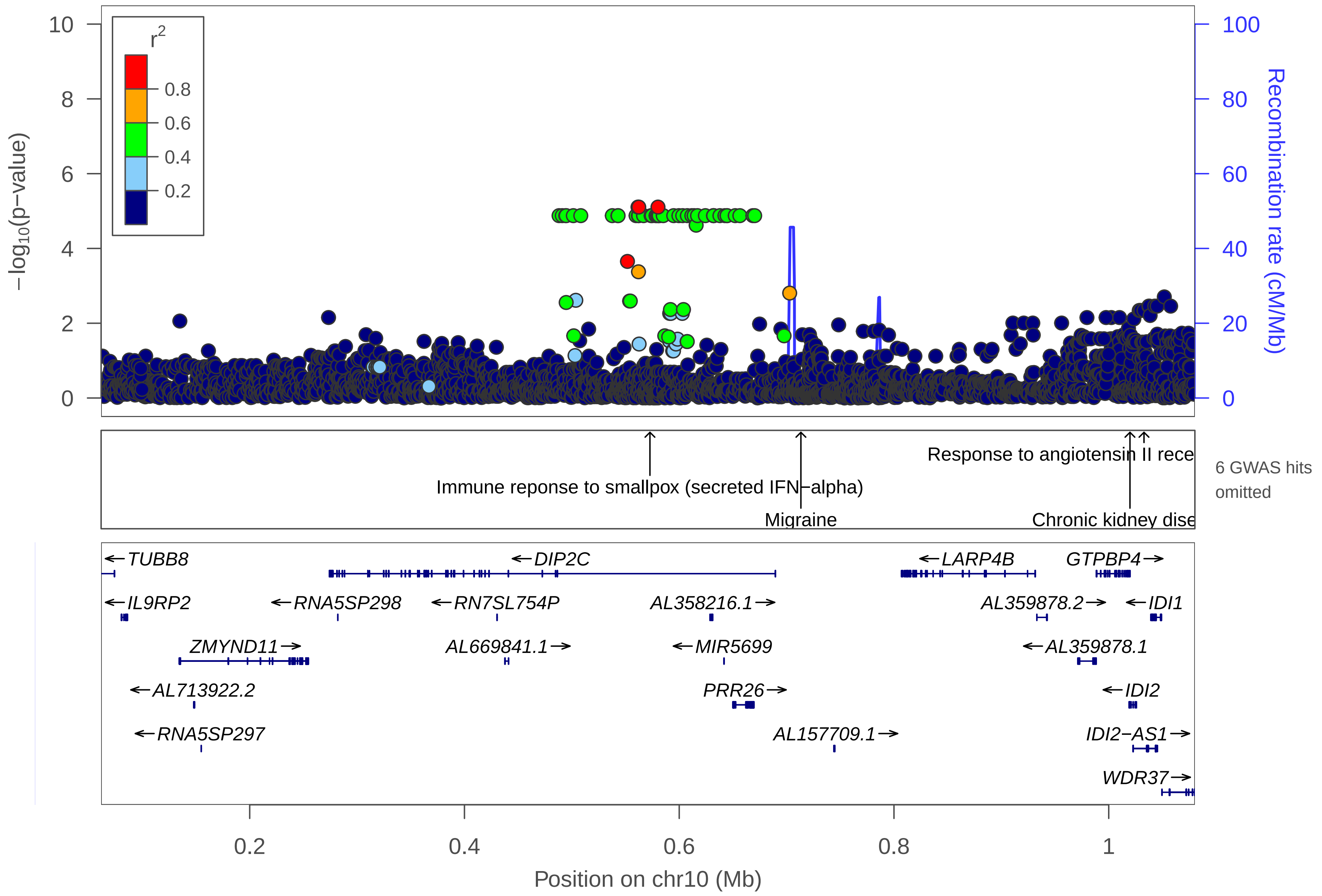
r2
0.8
0.6
0.4
0.2
−log10(p−value)
Recombination rate (cM/Mb)
Response to angiotensin II rece
Immune reponse to smallpox (secreted IFN−alpha)
Migraine
Chronic kidney dise
6 GWAS hits omitted
TUBB8
DIP2C
LARP4B
GTPBP4
IL9RP2
RNA5SP298
RN7SL754P
AL358216.1
AL359878.2
IDI1
ZMYND11
AL669841.1
MIR5699
AL359878.1
AL713922.2
PRR26
IDI2
RNA5SP297
AL157709.1
IDI2−AS1
WDR37
Position on chr10 (Mb)

| | |
|---|---|
| date: | Thu Oct 20 22:13:36 2022 |
| build: | hg38 |
| display range: | chr10:61630−1080236 [61630−1080236] |
| hilite range: | 0 − 0 [ 0 − 0 ] |
| reference SNP: | chr10:580238 |
| number of SNPs plotted: | 2940 |
| min P−value: | 7.82E−6 [chr10:561630] |
| max P−value: | 9.99E−1 [chr10:759496] |
| omitted GWAS Hits: | chr10:1.01977−Chronic kidney disease, NA |
| omitted GWAS Hits: | NA, NA |

Make more plots at http://csg.sph.umich.edu/locuszoom/

GWAS Catalog SNPs in Region

| chr | pos (Mb) | trait | snp |
|---|---|---|---|
| 10 | 0.272136 | Blood metabolite levels | rs12246027 |
| 10 | 0.378205 | Neuroticism | rs77829203 |
| 10 | 0.520439 | Psychosis in Alzheimer's disease | rs11252926 |
| 10 | 0.572745 | Immune reponse to smallpox (secreted IFN−alpha) | rs17221323 |
| 10 | 0.713328 | Migraine | rs7916968 |
| 10 | 0.725592 | Uric acid levels | rs877282 |
| 10 | 0.791467 | Survival in rectal cancer | rs1555895 |
| 10 | 1.019770 | Chronic kidney disease | rs1044261 |
| 10 | 1.019770 | Glomerular filtration rate (creatinine) | rs1044261 |
| 10 | 1.032842 | Response to angiotensin II receptor blocker therapy | rs12762955 |
