## Supplemental File 1 for "Genetic Associations with Age at Dementia Onset in the *PSEN1 E280A* Colombian Kindred": chr10_28292958-34410406.pdf

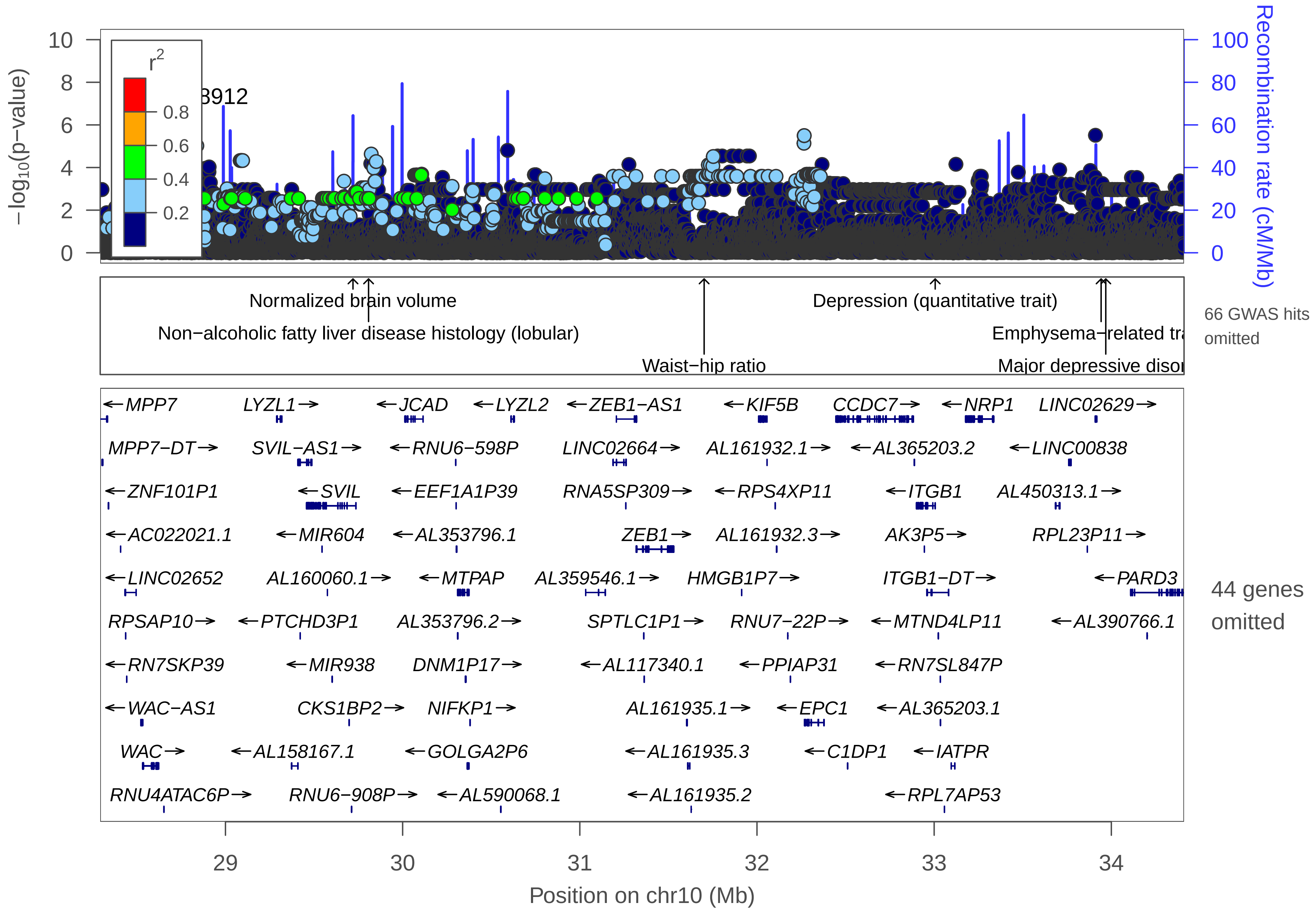

−log10(p−value)
Recombination rate (cM/Mb)
r2
0.8
0.6
0.4
0.2
3912
Normalized brain volume
Non−alcoholic fatty liver disease histology (lobular)
Waist−hip ratio
Depression (quantitative trait)
Emphysema−related tra
Major depressive disor
66 GWAS hits omitted
←MPP7
LYZL1→
←JCAD
←LYZL2
←ZEB1−AS1
←KIF5B
CCDC7→
←NRP1
LINC02629→
MPP7−DT→
SVIL−AS1→
←RNU6−598P
LINC02664→
AL161932.1→
←AL365203.2
←LINC00838
←ZNF101P1
←SVIL
←EEF1A1P39
RNA5SP309→
←RPS4XP11
←ITGB1
AL450313.1→
←AC022021.1
←MIR604
←AL353796.1
ZEB1→
AL161932.3→
AK3P5→
RPL23P11→
←LINC02652
AL160060.1→
←MTPAP
AL359546.1→
HMGB1P7→
ITGB1−DT→
←PARD3
RPSAP10→
←PTCHD3P1
AL353796.2→
SPTLC1P1→
RNU7−22P→
←MTND4LP11
←AL390766.1
←RN7SKP39
←MIR938
DNM1P17→
←AL117340.1
←PPIAP31
←RN7SL847P
←WAC−AS1
CKS1BP2→
NIFKP1→
AL161935.1→
←EPC1
←AL365203.1
WAC→
←AL158167.1
←GOLGA2P6
←AL161935.3
←C1DP1
←IATPR
RNU4ATAC6P→
RNU6−908P→
←AL590068.1
←AL161935.2
←RPL7AP53
44 genes omitted
29
30
31
32
33
34
Position on chr10 (Mb)

| | |
|---|---|
| date: | Thu Oct 20 22:13:40 2022 |
| build: | hg38 |
| display range: | chr10:28292958−34410406 [28292958−34410406] |
| hilite range: | 0 − 0 [ 0 − 0 ] |
| reference SNP: | chr10:28792958 |
| number of SNPs plotted: | 24287 |
| min P−value: | 5.25E−7 [chr10:28792958] |
| max P−value: | 10E−1 [chr10:32084757] |
| omitted Genes: | TPRKBP1, RNU6−1067P, BAMBI |
| omitted Genes: | LINC01517, AL355376.1, RNU6−270P |
| omitted Genes: | LINC00837, AL355376.2, RNA5SP308 |
| omitted Genes: | RPL21P93, AL353093.1, RN7SL241P |
| omitted Genes: | MIR7162, AL161651.1, CCND3P1 |
| omitted Genes: | MAP3K8, HNRNPA1P32, AL590068.4 |
| omitted Genes: | RN7SL63P, AL590068.2, AL590068.3 |
| omitted Genes: | SVIL2P, LINC02644, ZNF438 |
| omitted Genes: | AL359532.1, DDX10P1, MACORIS |
| omitted Genes: | AL353769.1, ARHGAP12, RPL34P19 |
| omitted Genes: | RN7SL825P, AL161932.2, RPS24P13 |
| omitted Genes: | AL158834.2, AL158834.1, AL391839.2 |
| omitted Genes: | AL391839.1, RNU6−1244P, AL391839.3 |
| omitted Genes: | AL121748.1, RN7SL398P, AL353600.1 |
| omitted Genes: | AL353600.2, LINC02628 |

Make more plots at http://csg.sph.umich.edu/locuszoom/

GWAS Catalog SNPs in Region

| chr | pos (Mb) | trait | snp |
|---|---|---|---|
| 10 | 28.30145 | Diisocyanate−induced asthma | rs115882204 |
| 10 | 28.32871 | Pulmonary function decline | rs1148186 |
| 10 | 28.34873 | Lung cancer | rs7900823 |
| 10 | 28.42068 | Pelvic organ prolapse | rs10826468 |
| 10 | 28.46318 | Obesity−related traits | rs2153299 |
| 10 | 28.46633 | Aggressiveness in attention deficit hyperactivity disorder | rs117063229 |
| 10 | 28.56309 | Alzheimer disease and age of onset | rs181666690 |
| 10 | 28.68808 | Percentage gas trapping | rs655766 |
| 10 | 28.73829 | Preeclampsia | rs1248993 |
| 10 | 28.94747 | Aggressiveness in attention deficit hyperactivity disorder | rs10826545 |
| 10 | 28.94853 | Aggressiveness in attention deficit hyperactivity disorder | rs10826546 |
| 10 | 28.95728 | Aggressiveness in attention deficit hyperactivity disorder | rs117352156 |
| 10 | 28.95732 | Aggressiveness in attention deficit hyperactivity disorder | rs188370812 |
| 10 | 28.95732 | Aggressiveness in attention deficit hyperactivity disorder | rs192649950 |
| 10 | 28.96666 | Aggressiveness in attention deficit hyperactivity disorder | rs10826548 |
| 10 | 29.00010 | Cannabis dependence | rs11007350 |
| 10 | 29.00297 | Major depressive disorder (broad) | rs1612122 |
| 10 | 29.04792 | Palmitoleic acid (16:1n−7) plasma levels | rs788076 |
| 10 | 29.07577 | Migraine with aura | rs10826566 |
| 10 | 29.09798 | Gut microbiota (bacterial taxa) | rs1248290 |
| 10 | 29.09971 | Gut microbiota (bacterial taxa) | rs1889714 |

GWAS Catalog SNPs in Region

| chr | pos (Mb) | trait | snp |
|---|---|---|---|
| 10 | 29.19844 | Methotrexate phramacokinetics (acute lymphoblastic leukemia) | rs4387258 |
| 10 | 29.52367 | Platelet thrombus formation | rs7070678 |
| 10 | 29.71973 | Normalized brain volume | rs1927457 |
| 10 | 29.79843 | Migraine with aura | rs2986961 |
| 10 | 29.80770 | Non−alcoholic fatty liver disease histology (lobular) | rs2986971 |
| 10 | 29.88896 | Orofacial clefts | rs3006564 |
| 10 | 29.94094 | Alzheimer disease and age of onset | rs146650065 |
| 10 | 29.94526 | Emphysema distribution in smoking | rs35374984 |
| 10 | 30.02714 | Coronary heart disease | rs3739998 |
| 10 | 30.03496 | Coronary artery disease | rs2487928 |
| 10 | 30.04619 | Coronary heart disease | rs2505083 |
| 10 | 30.04619 | Myocardial infarction | rs2505083 |
| 10 | 30.20178 | Suicide risk | rs2462021 |
| 10 | 30.23090 | Pancreatitis | rs2995271 |
| 10 | 30.28801 | 3−hydroxypropylmercapturic acid levels in smokers | rs12220014 |
| 10 | 30.40257 | Inflammatory bowel disease | rs2050392 |
| 10 | 30.43917 | Crohn's disease | rs1042058 |
| 10 | 30.43917 | Inflammatory bowel disease | rs1042058 |
| 10 | 30.54570 | QT interval | rs11008099 |
| 10 | 30.61036 | Bone mineral density (femoral neck) | rs73245065 |
| 10 | 30.62310 | Dental caries | rs399593 |

GWAS Catalog SNPs in Region

| chr | pos (Mb) | trait | snp |
|---|---|---|---|
| 10 | 30.75356 | Congenital left−sided heart lesions (maternal effect) | rs11008222 |
| 10 | 30.83824 | Height | rs12413361 |
| 10 | 31.07734 | Response to antipsychotic treatment | rs2994684 |
| 10 | 31.11547 | Late−onset Alzheimer's disease | rs141387448 |
| 10 | 31.12618 | Multiple sclerosis | rs793108 |
| 10 | 31.12618 | Rheumatoid arthritis | rs793108 |
| 10 | 31.17267 | Glucose homeostasis traits | rs1148233 |
| 10 | 31.20705 | Disc degeneration (lumbar) | rs1250307 |
| 10 | 31.22317 | Disc degeneration (lumbar) | rs2484992 |
| 10 | 31.22620 | Disc degeneration (lumbar) | rs2484990 |
| 10 | 31.67980 | C−reactive protein levels | rs796127 |
| 10 | 31.69246 | Cannabis dependence | rs115553536 |
| 10 | 31.70170 | Waist−hip ratio | rs7081678 |
| 10 | 31.99136 | Alzheimer disease and age of onset | rs211257 |
| 10 | 32.00401 | Schizophrenia | rs1251350 |
| 10 | 32.02008 | Bipolar disorder with mood−incongruent psychosis | rs1775715 |
| 10 | 32.33609 | Serum dimethylarginine levels (asymmetric/symetric ratio) | rs143713361 |
| 10 | 32.34604 | Sexual dysfunction (female) | rs2370759 |
| 10 | 32.70434 | Suicide in bipolar disorder | rs7079041 |
| 10 | 32.87929 | Post bronchodilator FEV1/FVC ratio | rs183323229 |
| 10 | 33.00585 | Depression (quantitative trait) | rs11009175 |

GWAS Catalog SNPs in Region

| chr | pos (Mb) | trait | snp |
| --- | --- | --- | --- |
| 10 | 33.11616 | IgG glycosylation | rs7902627 |
| 10 | 33.17920 | Migraine | rs2506142 |
| 10 | 33.18582 | Parental extreme longevity (95 years and older) | rs12768811 |
| 10 | 33.18635 | Tetralogy of Fallot | rs2228638 |
| 10 | 33.21425 | Migraine | rs2506155 |
| 10 | 33.32711 | Parental extreme longevity (95 years and older) | rs2804497 |
| 10 | 33.79912 | Schizophrenia | rs1412115 |
| 10 | 33.94235 | Emphysema−related traits | rs7905537 |
| 10 | 33.96868 | Major depressive disorder | rs1780436 |
