## Supplemental File 1 for "Genetic Associations with Age at Dementia Onset in the *PSEN1 E280A* Colombian Kindred": chr10_48668045-49668045.pdf

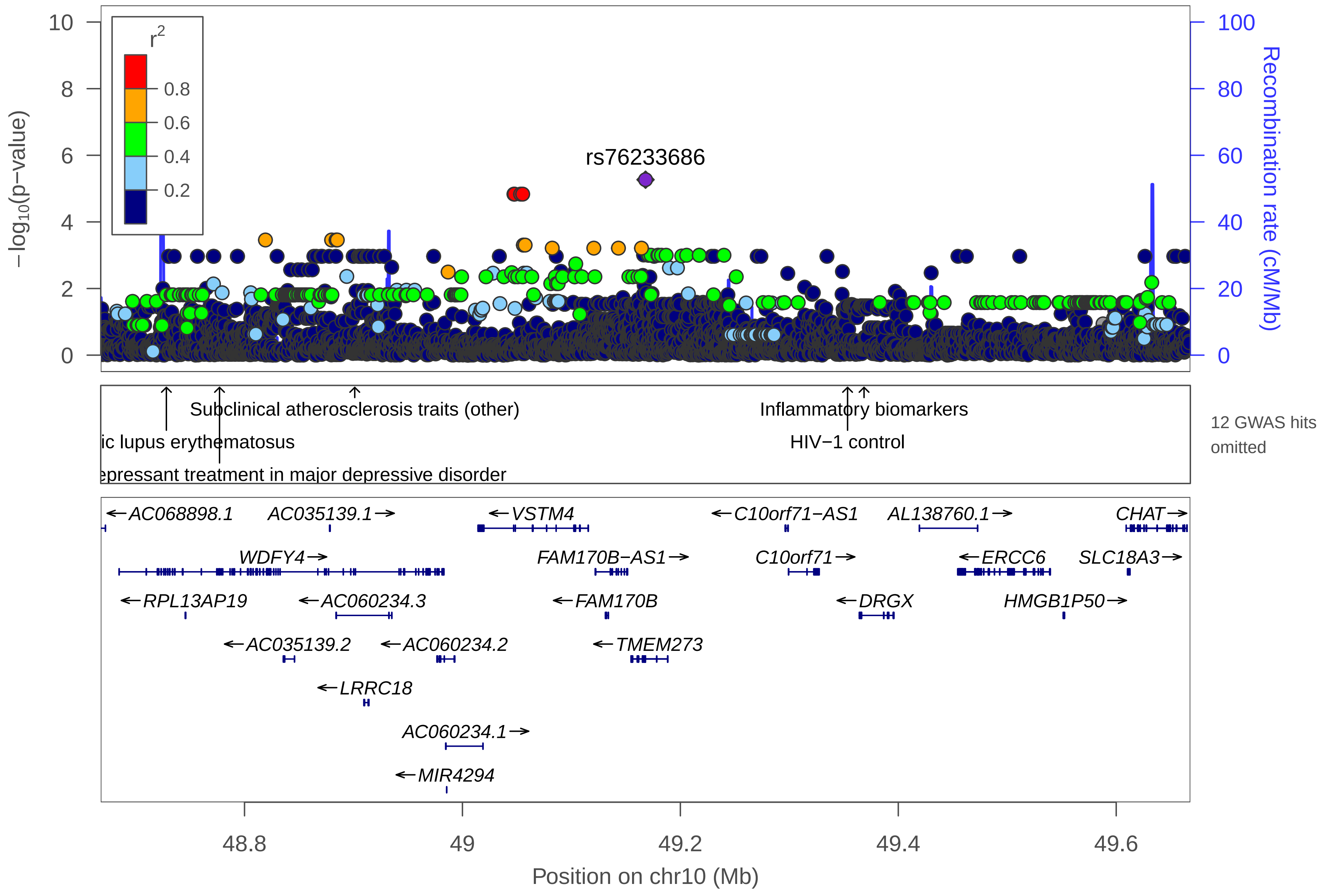

rs76233686
r2
0.8
0.6
0.4
0.2
−log10(p−value)
Recombination rate (cM/Mb)
Subclinical atherosclerosis traits (other)
ic lupus erythematosus
epressant treatment in major depressive disorder
Inflammatory biomarkers
HIV−1 control
12 GWAS hits omitted
AC068898.1
AC035139.1
VSTM4
C10orf71−AS1
AL138760.1
CHAT
WDFY4
FAM170B−AS1
C10orf71
ERCC6
SLC18A3
RPL13AP19
AC060234.3
FAM170B
DRGX
HMGB1P50
AC035139.2
AC060234.2
TMEM273
LRRC18
AC060234.1
MIR4294
Position on chr10 (Mb)

| | |
|---|---|
| date: | Thu Oct 20 22:13:35 2022 |
| build: | hg38 |
| display range: | chr10:48668045−49668045 [48668045−49668045] |
| hilite range: | 0 − 0 [ 0 − 0 ] |
| reference SNP: | chr10:49168045 |
| number of SNPs plotted: | 4483 |
| min P−value: | 5.41E−6 [chr10:49168045] |
| max P−value: | 10E−1 [chr10:49018759] |
| omitted GWAS Hits: | chr10:49.353462−HIV−1 control, NA |
| omitted GWAS Hits: | NA, NA |
| omitted GWAS Hits: | NA, NA |
| omitted GWAS Hits: | NA, NA |

Make more plots at http://csg.sph.umich.edu/locuszoom/

GWAS Catalog SNPs in Region

| chr | pos (Mb) | trait | snp |
|---|---|---|---|
| 10 | 48.70331 | Stroke | rs17771318 |
| 10 | 48.72829 | Systemic lupus erythematosus | rs11101442 |
| 10 | 48.77707 | Response to anti−depressant treatment in major depressive disorder | rs10857636 |
| 10 | 48.81012 | Systemic lupus erythematosus | rs2928402 |
| 10 | 48.81735 | Systemic lupus erythematosus | rs7097397 |
| 10 | 48.83491 | Systemic lupus erythematosus | rs877819 |
| 10 | 48.86135 | Systemic lupus erythematosus | rs2663052 |
| 10 | 48.88977 | Rheumatoid arthritis | rs2671692 |
| 10 | 48.89503 | Airway responsiveness in chronic obstructive pulmonary disease | rs12252129 |
| 10 | 48.90118 | Subclinical atherosclerosis traits (other) | rs3849150 |
| 10 | 48.91101 | Systemic lupus erythematosus | rs1913517 |
| 10 | 49.02159 | Migraine with aura | rs2137920 |
| 10 | 49.24589 | Plasma omega−6 polyunsaturated fatty acid levels (linoleic acid) | rs17009617 |
| 10 | 49.29241 | Oleic acid (18:1n−9) plasma levels | rs17774576 |
| 10 | 49.31570 | Alzheimer disease and age of onset | rs117850870 |
| 10 | 49.35346 | HIV−1 control | rs4838508 |
| 10 | 49.36859 | Inflammatory biomarkers | rs12220898 |
