## Supplemental File 1 for "Genetic Associations with Age at Dementia Onset in the *PSEN1 E280A* Colombian Kindred": chr12_88820371-89820371.pdf

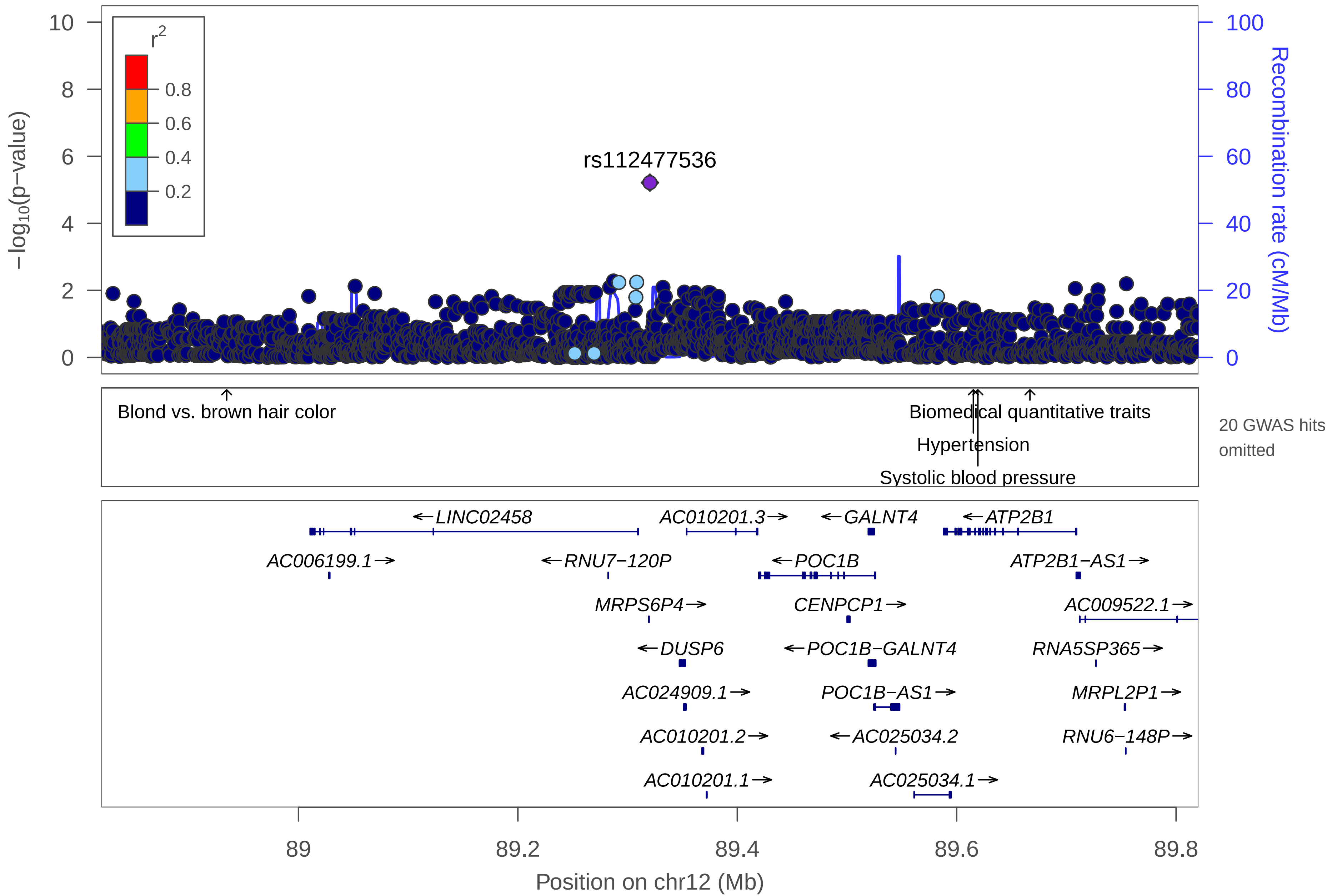

rs112477536
r2
0.8
0.6
0.4
0.2
−log10(p−value)
Recombination rate (cM/Mb)
Blond vs. brown hair color
Biomedical quantitative traits
Hypertension
Systolic blood pressure
20 GWAS hits omitted
←LINC02458
AC010201.3→
←GALNT4
←ATP2B1
AC006199.1→
←RNU7−120P
←POC1B
ATP2B1−AS1→
MRPS6P4→
CENPCP1→
AC009522.1→
←DUSP6
←POC1B−GALNT4
RNA5SP365→
AC024909.1→
POC1B−AS1→
MRPL2P1→
AC010201.2→
←AC025034.2
RNU6−148P→
AC010201.1→
AC025034.1→
Position on chr12 (Mb)

| | |
|---|---|
| date: | Thu Oct 20 22:13:34 2022 |
| build: | hg38 |
| display range: | chr12:88820371−89820371 [88820371−89820371] |
| hilite range: | 0 − 0 [ 0 − 0 ] |
| reference SNP: | chr12:89320371 |
| number of SNPs plotted: | 2375 |
| min P−value: | 6.12E−6 [chr12:89320371] |
| max P−value: | 9.99E−1 [chr12:89234726] |
| omitted GWAS Hits: | NA, NA |
| omitted GWAS Hits: | NA, NA |
| omitted GWAS Hits: | NA, NA |
| omitted GWAS Hits: | NA, NA |
| omitted GWAS Hits: | NA, NA |
| omitted GWAS Hits: | NA, NA |
| omitted GWAS Hits: | NA, NA |

Make more plots at http://csg.sph.umich.edu/locuszoom/

GWAS Catalog SNPs in Region

| chr | pos (Mb) | trait | snp |
|---|---|---|---|
| 12 | 88.89548 | Bone mineral density (femoral neck) | rs191780267 |
| 12 | 88.93456 | Blond vs. brown hair color | rs12821256 |
| 12 | 88.93456 | Hair color | rs12821256 |
| 12 | 88.93456 | Blond vs non−blond hair color | rs12821256 |
| 12 | 88.93456 | Brown vs. non−brown hair color | rs12821256 |
| 12 | 88.93456 | Light vs. dark hair color | rs12821256 |
| 12 | 89.38182 | Night sleep phenotypes | rs61925087 |
| 12 | 89.44709 | Schizophrenia | rs10777165 |
| 12 | 89.50361 | NT−proBNP levels in acute coronary syndrome | rs11105306 |
| 12 | 89.54777 | Blood pressure | rs4842666 |
| 12 | 89.61518 | Hypertension | rs2681472 |
| 12 | 89.61518 | Diastolic blood pressure | rs2681472 |
| 12 | 89.61518 | Coronary artery disease | rs2681472 |
| 12 | 89.61518 | Myocardial infarction | rs2681472 |
| 12 | 89.61518 | Systolic blood pressure | rs2681472 |
| 12 | 89.61931 | Systolic blood pressure | rs2681492 |
| 12 | 89.66681 | Biomedical quantitative traits | rs17249754 |
| 12 | 89.66681 | Blood pressure | rs17249754 |
| 12 | 89.66681 | Systolic blood pressure | rs17249754 |
| 12 | 89.66681 | Diastolic blood pressure | rs17249754 |
| 12 | 89.66681 | Hypertension | rs17249754 |

GWAS Catalog SNPs in Region

| chr | pos (Mb) | trait | snp |
|---|---|---|---|
| 12 | 89.66681 | Mean arterial pressure | rs17249754 |
| 12 | 89.66681 | Pulse pressure | rs17249754 |
| 12 | 89.68741 | Coronary heart disease | rs7136259 |
