## Supplemental File 1 for "Genetic Associations with Age at Dementia Onset in the *PSEN1 E280A* Colombian Kindred": chr13_45614229-46624586.pdf

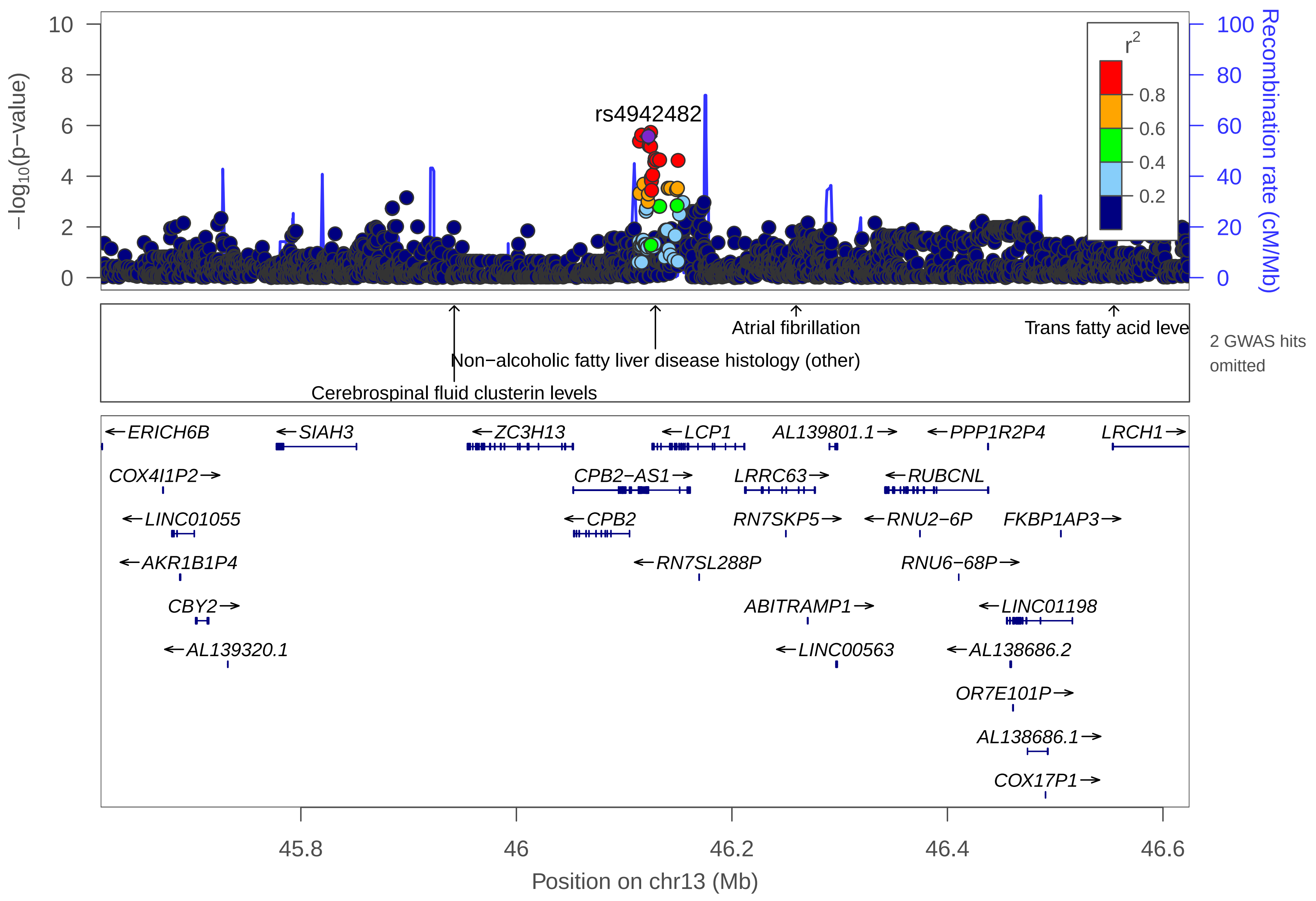

rs4942482
−log10(p−value)
Recombination rate (cM/Mb)
r2
0.8
0.6
0.4
0.2
Atrial fibrillation
Trans fatty acid leve
Non−alcoholic fatty liver disease histology (other)
Cerebrospinal fluid clusterin levels
2 GWAS hits omitted
←ERICH6B
←SIAH3
←ZC3H13
←LCP1
AL139801.1→
←PPP1R2P4
LRCH1→
COX4I1P2→
CPB2−AS1→
LRRC63→
←RUBCNL
←LINC01055
←CPB2
RN7SKP5→
←RNU2−6P
FKBP1AP3→
←AKR1B1P4
←RN7SL288P
RNU6−68P→
CBY2→
ABITRAMP1→
←LINC01198
←AL139320.1
←LINC00563
←AL138686.2
OR7E101P→
AL138686.1→
COX17P1→
45.8
46
46.2
46.4
46.6
Position on chr13 (Mb)

| | |
|---|---|
| date: | Thu Oct 20 22:13:34 2022 |
| build: | hg38 |
| display range: | chr13:45614229−46624586 [45614229−46624586] |
| hilite range: | 0 − 0 [ 0 − 0 ] |
| reference SNP: | chr13:46122525 |
| number of SNPs plotted: | 3276 |
| min P−value: | 1.87E−6 [chr13:46124586] |
| max P−value: | 9.99E−1 [chr13:45925981] |
| omitted GWAS Hits: | NA, NA |

Make more plots at http://csg.sph.umich.edu/locuszoom/

GWAS Catalog SNPs in Region

| chr | pos (Mb) | trait | snp |
|---|---|---|---|
| 13 | 45.83390 | Pneumococcal bacteremia | rs9526114 |
| 13 | 45.94233 | Cerebrospinal fluid clusterin levels | rs741668 |
| 13 | 46.12901 | Non−alcoholic fatty liver disease histology (other) | rs7324845 |
| 13 | 46.21139 | Systemic lupus erythematosus | rs912784 |
| 13 | 46.25958 | Atrial fibrillation | rs958546 |
| 13 | 46.55447 | Trans fatty acid levels | rs7325799 |
