## Supplemental File 1 for "Genetic Associations with Age at Dementia Onset in the *PSEN1 E280A* Colombian Kindred": chr14_74903852-79621238.pdf

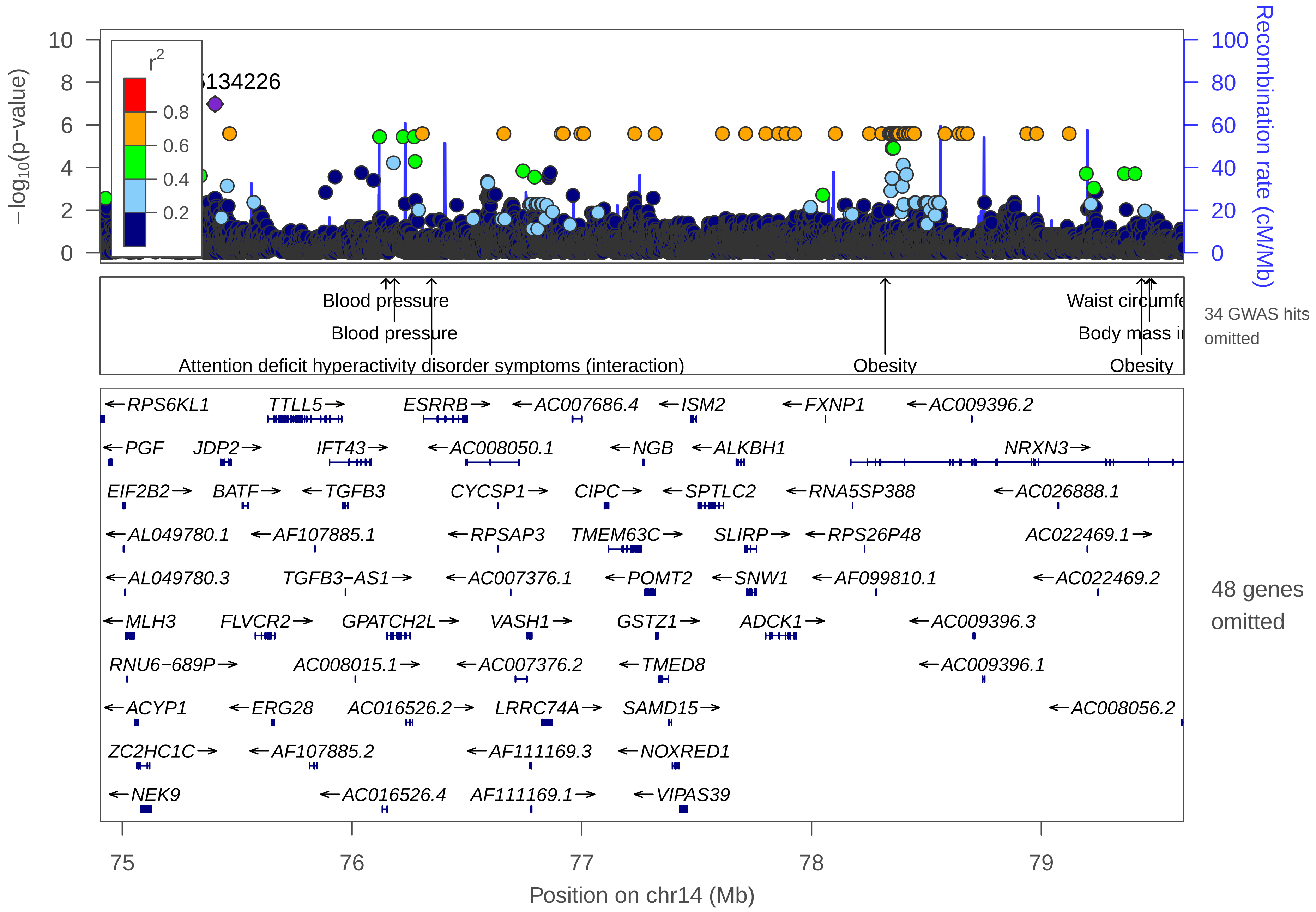

−log10(p−value)
Recombination rate (cM/Mb)
r2
0.8
0.6
0.4
0.2
5134226
10
8
6
4
2
0
100
80
60
40
20
0
Blood pressure
Blood pressure
Attention deficit hyperactivity disorder symptoms (interaction)
Obesity
Waist circumfe
Body mass i
Obesity
34 GWAS hits omitted
←RPS6KL1
TTLL5→
ESRRB→
←AC007686.4
←ISM2
←FXNP1
←AC009396.2
←PGF
JDP2→
IFT43→
←AC008050.1
←NGB
←ALKBH1
NRXN3→
EIF2B2→
BATF→
←TGFB3
CYCSP1→
CIPC→
←SPTLC2
←RNA5SP388
←AC026888.1
←AL049780.1
←AF107885.1
←RPSAP3
TMEM63C→
SLIRP→
←RPS26P48
AC022469.1→
←AL049780.3
TGFB3−AS1→
←AC007376.1
←POMT2
←SNW1
←AF099810.1
←AC022469.2
←MLH3
FLVCR2→
GPATCH2L→
VASH1→
GSTZ1→
ADCK1→
←AC009396.3
RNU6−689P→
AC008015.1→
←AC007376.2
←TMED8
←AC009396.1
←ACYP1
←ERG28
AC016526.2→
LRRC74A→
SAMD15→
←AC008056.2
ZC2HC1C→
←AF107885.2
←AF111169.3
←NOXRED1
←NEK9
←AC016526.4
AF111169.1→
←VIPAS39
48 genes omitted
75
76
77
78
79
Position on chr14 (Mb)

| | |
|---|---|
| date: | Thu Oct 20 22:13:37 2022 |
| build: | hg38 |
| display range: | chr14:74903852−79621238 [74903852−79621238] |
| hilite range: | 0 − 0 [ 0 − 0 ] |
| reference SNP: | chr14:75403852 |
| number of SNPs plotted: | 11574 |
| min P−value: | 1.06E−7 [chr14:75403852] |
| max P−value: | 10E−1 [chr14:75900744] |
| omitted Genes: | HIF1AP1, AL049780.2, TMED10 |
| omitted Genes: | AL691403.2, RNU4ATAC14P, AL691403.1 |
| omitted Genes: | AF111167.1, AF111167.3, FOS |
| omitted Genes: | DPPA5P4, LINC01220, AF111167.2 |
| omitted Genes: | AC007182.1, AC007182.2, RNA5SP387 |
| omitted Genes: | AC007182.3, AC016526.3, AC016526.1 |
| omitted Genes: | VASH1−AS1, ANGEL1, AF111169.4 |
| omitted Genes: | RPL22P2, RN7SKP17, AF111169.2 |
| omitted Genes: | LINC01629, RN7SL356P, AC007686.1 |
| omitted Genes: | IRF2BPL, AC007686.3, LINC02288 |
| omitted Genes: | AC007686.2, LINC02289, AC007686.5 |
| omitted Genes: | AC007375.2, ZDHHC22, AC007375.3 |
| omitted Genes: | FAM204DP, AC007375.1, MIR1260A |
| omitted Genes: | AC007954.1, AHSA1, AF111168.1 |
| omitted Genes: | RN7SL587P, COX6CP11, RPL21P10 |

Make more plots at http://csg.sph.umich.edu/locuszoom/

GWAS Catalog SNPs in Region

| chr | pos (Mb) | trait | snp |
|---|---|---|---|
| 14 | 75.15934 | Height | rs910316 |
| 14 | 75.23452 | Inflammatory bowel disease | rs4899554 |
| 14 | 75.27505 | Crohn's disease | rs1569328 |
| 14 | 75.27505 | Inflammatory bowel disease | rs1569328 |
| 14 | 75.34732 | Periodontal microbiota | rs11621969 |
| 14 | 75.44976 | Obesity−related traits | rs84044 |
| 14 | 75.49419 | Rheumatoid arthritis | rs7155603 |
| 14 | 75.53921 | Multiple sclerosis | rs2300603 |
| 14 | 75.66325 | Height | rs724743 |
| 14 | 75.70752 | Large artery stroke | rs1005224 |
| 14 | 76.03396 | Acute urticaria and angioedema (non−steroidal anti−inflammatory drug−induced) | rs9323624 |
| 14 | 76.14733 | Blood pressure | rs935334 |
| 14 | 76.18442 | Blood pressure | rs2121070 |
| 14 | 76.23701 | Acute lymphoblastic leukemia (childhood) | rs7156960 |
| 14 | 76.34615 | Attention deficit hyperactivity disorder symptoms (interaction) | rs2360997 |
| 14 | 76.34635 | Maximal oxygen uptake response | rs12893597 |
| 14 | 77.03163 | Adverse response to lamotrigine and phenytoin | rs183266 |
| 14 | 77.04750 | Sleep quality | rs1986116 |
| 14 | 77.13548 | Large artery stroke | rs1465330 |
| 14 | 77.21833 | Bipolar disorder | rs4467006 |
| 14 | 77.21850 | Bipolar disorder | rs2287375 |

GWAS Catalog SNPs in Region

| chr | pos (Mb) | trait | snp |
|---|---|---|---|
| 14 | 77.23762 | Obsessive−compulsive symptoms | rs67366981 |
| 14 | 77.56203 | Gut microbiome composition (winter) | rs4903604 |
| 14 | 77.91334 | Resting heart rate | rs1549118 |
| 14 | 77.95471 | Response to anti−retroviral therapy (ddI/d4T) in HIV−1 infection (Grade 1 peripheral neuropathy) | rs4899685 |
| 14 | 78.06856 | Response to platinum−based chemotherapy in non−small−cell lung cancer | rs3850370 |
| 14 | 78.09012 | Objective response to lithium treatment in bipolar disorder | rs809 |
| 14 | 78.22872 | Electrodermal activity | rs10129666 |
| 14 | 78.28464 | Electrodermal activity | rs11159346 |
| 14 | 78.30896 | Brain structure (temporal lobe volume) | rs7155434 |
| 14 | 78.31973 | Obesity | rs11624704 |
| 14 | 78.31973 | Coronary artery aneurysm in Kawasaki disease | rs11624704 |
| 14 | 78.31982 | Cognitive performance | rs6574433 |
| 14 | 78.74307 | Body mass index | rs17835706 |
| 14 | 79.38089 | Underweight status | rs12882679 |
| 14 | 79.43311 | Obesity | rs7141420 |
| 14 | 79.43311 | Body mass index | rs7141420 |
| 14 | 79.43703 | Obesity | rs2370983 |
| 14 | 79.47062 | Body mass index | rs10150332 |
| 14 | 79.47882 | Waist circumference | rs10146997 |
| 14 | 79.53575 | Amyotrophic lateral sclerosis (age of onset) | rs7147705 |
