## Supplemental File 1 for "Genetic Associations with Age at Dementia Onset in the *PSEN1 E280A* Colombian Kindred": chr14_74903852-79621238_rs184135706.pdf

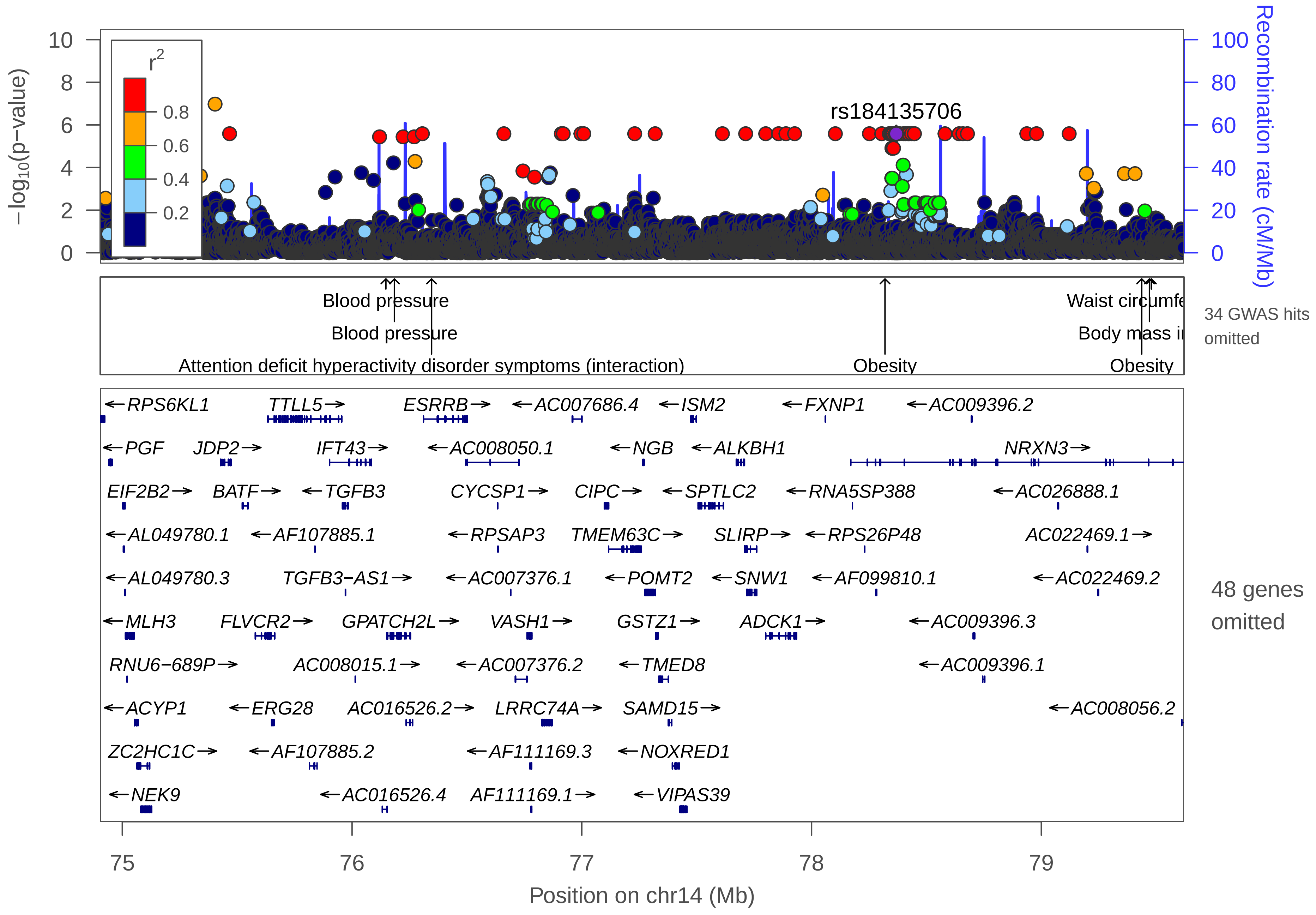

r2
0.8
0.6
0.4
0.2
−log10(p−value)
Recombination rate (cM/Mb)
rs184135706
Blood pressure
Blood pressure
Attention deficit hyperactivity disorder symptoms (interaction)
Obesity
Waist circumfe
Body mass in
Obesity
34 GWAS hits omitted
48 genes omitted
RPS6KL1
TTLL5
ESRRB
AC007686.4
ISM2
FXNP1
AC009396.2
PGF
JDP2
IFT43
AC008050.1
NGB
ALKBH1
NRXN3
EIF2B2
BATF
TGFB3
CYCSP1
CIPC
SPTLC2
RNA5SP388
AC026888.1
AL049780.1
AF107885.1
RPSAP3
TMEM63C
SLIRP
RPS26P48
AC022469.1
AL049780.3
TGFB3−AS1
AC007376.1
POMT2
SNW1
AF099810.1
AC022469.2
MLH3
FLVCR2
GPATCH2L
VASH1
GSTZ1
ADCK1
AC009396.3
RNU6−689P
AC008015.1
AC007376.2
TMED8
AC009396.1
ACYP1
ERG28
AC016526.2
LRRC74A
SAMD15
AC008056.2
ZC2HC1C
AF107885.2
AF111169.3
NOXRED1
NEK9
AC016526.4
AF111169.1
VIPAS39
Position on chr14 (Mb)
