## Supplemental File 1 for "Genetic Associations with Age at Dementia Onset in the *PSEN1 E280A* Colombian Kindred": chr15_71014241-72788879.pdf

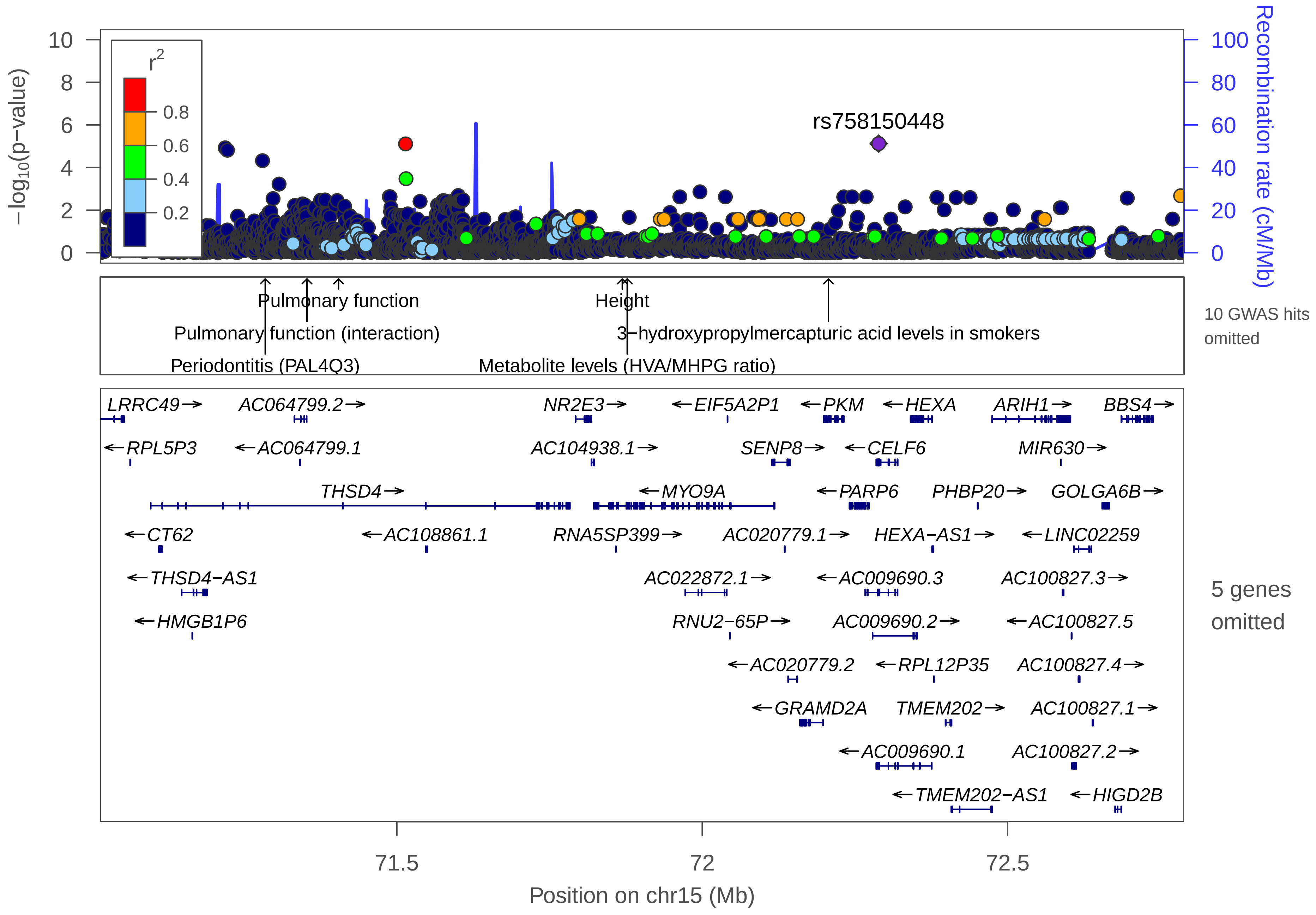

rs758150448
−log10(p−value)
Recombination rate (cM/Mb)
r2
0.8
0.6
0.4
0.2
Pulmonary function
Pulmonary function (interaction)
Periodontitis (PAL4Q3)
Height
3−hydroxypropylmercapturic acid levels in smokers
Metabolite levels (HVA/MHPG ratio)
10 GWAS hits omitted
LRRC49
AC064799.2
NR2E3
EIF5A2P1
PKM
HEXA
ARIH1
BBS4
RPL5P3
AC064799.1
AC104938.1
SENP8
CELF6
MIR630
THSD4
MYO9A
PARP6
PHBP20
GOLGA6B
CT62
AC108861.1
RNA5SP399
AC020779.1
HEXA−AS1
LINC02259
THSD4−AS1
AC022872.1
AC009690.3
AC100827.3
HMGB1P6
RNU2−65P
AC009690.2
AC100827.5
AC020779.2
RPL12P35
AC100827.4
GRAMD2A
TMEM202
AC100827.1
AC009690.1
AC100827.2
TMEM202−AS1
HIGD2B
5 genes omitted
71.5
72
72.5
Position on chr15 (Mb)

| | |
|---|---|
| date: | Thu Oct 20 22:13:34 2022 |
| build: | hg38 |
| display range: | chr15:71014241−72788879 [71014241−72788879] |
| hilite range: | 0 − 0 [ 0 − 0 ] |
| reference SNP: | chr15:72288879 |
| number of SNPs plotted: | 3914 |
| min P−value: | 7.55E−6 [chr15:72288879] |
| max P−value: | 9.99E−1 [chr15:72330977] |
| omitted Genes: | AC079322.1, RN7SL853P, AC009712.1 |
| omitted Genes: | ADPGK, ADPGK−AS1 |
| omitted GWAS Hits: | chr15:71.869062−Height, chr15:72.206596−3−hydroxypropylmercapturic acid levels in smokers |
| omitted GWAS Hits: | NA, NA |
| omitted GWAS Hits: | NA, NA |
| omitted GWAS Hits: | NA |

Make more plots at http://csg.sph.umich.edu/locuszoom/

GWAS Catalog SNPs in Region

| chr | pos (Mb) | trait | snp |
|---|---|---|---|
| 15 | 71.13167 | Thiazide−induced adverse metabolic effects in hypertensive patients | rs12904863 |
| 15 | 71.28061 | Symmetrical dimethylarginine levels | rs76917273 |
| 15 | 71.28246 | Periodontitis (PAL4Q3) | rs9806183 |
| 15 | 71.28442 | Periodontitis (PAL4Q3) | rs1442779 |
| 15 | 71.28442 | Periodontitis (Mean PAL) | rs1442779 |
| 15 | 71.31628 | Antibody status in Tripanosoma cruzi seropositivity | rs17786786 |
| 15 | 71.35278 | Pulmonary function (interaction) | rs12899618 |
| 15 | 71.35278 | Pulmonary function | rs12899618 |
| 15 | 71.38762 | Airflow obstruction | rs2044029 |
| 15 | 71.40455 | Pulmonary function | rs12441227 |
| 15 | 71.44975 | Alzheimer disease and age of onset | rs142200609 |
| 15 | 71.81483 | 3−hydroxypropylmercapturic acid levels in smokers | rs113441626 |
| 15 | 71.86906 | Height | rs12902421 |
| 15 | 71.87713 | Metabolite levels (HVA/MHPG ratio) | rs12050794 |
| 15 | 72.20660 | 3−hydroxypropylmercapturic acid levels in smokers | rs113920924 |
| 15 | 72.51906 | Posterior cortical atrophy and Alzheimer's disease | rs8038734 |
