## Supplemental File 1 for "Genetic Associations with Age at Dementia Onset in the *PSEN1 E280A* Colombian Kindred": chr16_1-701813.pdf

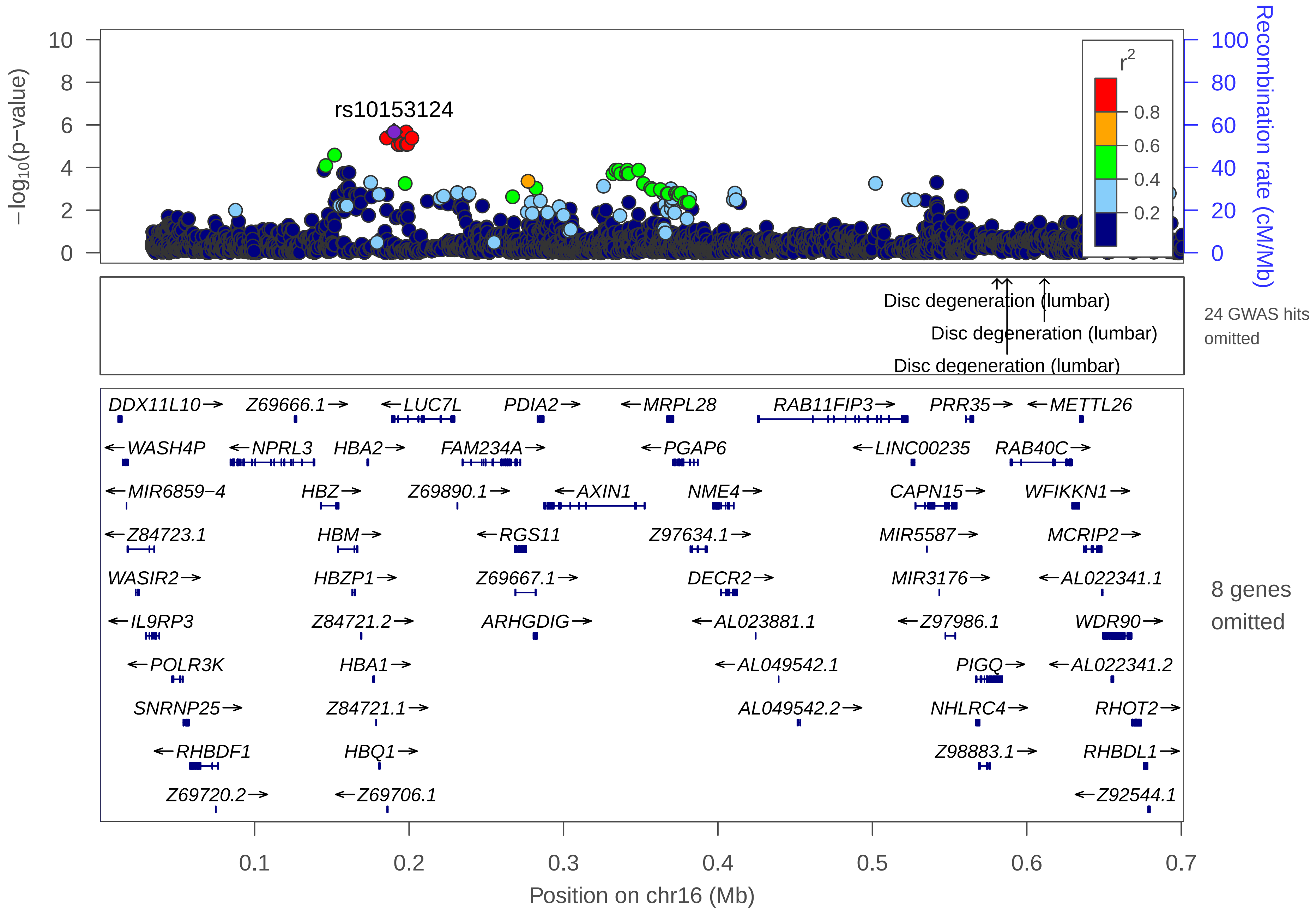

rs10153124
−log10(p−value)
Recombination rate (cM/Mb)
r2
0.8
0.6
0.4
0.2
10
8
6
4
2
0
100
80
60
40
20
0
Disc degeneration (lumbar)
Disc degeneration (lumbar)
Disc degeneration (lumbar)
24 GWAS hits omitted
DDX11L10→
Z69666.1→
←LUC7L
PDIA2→
←MRPL28
RAB11FIP3→
PRR35→
←METTL26
←WASH4P
←NPRL3
HBA2→
FAM234A→
←PGAP6
←LINC00235
RAB40C→
←MIR6859−4
HBZ→
Z69890.1→
←AXIN1
NME4→
CAPN15→
WFIKKN1→
←Z84723.1
HBM→
←RGS11
Z97634.1→
MIR5587→
MCRIP2→
WASIR2→
HBZP1→
Z69667.1→
DECR2→
MIR3176→
←AL022341.1
←IL9RP3
Z84721.2→
ARHGDIG→
←AL023881.1
←Z97986.1
WDR90→
←POLR3K
HBA1→
←AL049542.1
PIGQ→
←AL022341.2
SNRNP25→
Z84721.1→
AL049542.2→
NHLRC4→
RHOT2→
←RHBDF1
HBQ1→
Z98883.1→
RHBDL1→
Z69720.2→
←Z69706.1
←Z92544.1
8 genes omitted
0.1
0.2
0.3
0.4
0.5
0.6
0.7
Position on chr16 (Mb)

| | |
|---|---|
| date: | Thu Oct 20 22:13:35 2022 |
| build: | hg38 |
| display range: | chr16:1−701813 [1−701813] |
| hilite range: | 0 − 0 [ 0 − 0 ] |
| reference SNP: | chr16:190339 |
| number of SNPs plotted: | 2385 |
| min P−value: | 2.15E−6 [chr16:190339] |
| max P−value: | 10E−1 [chr16:347820] |
| omitted Genes: | MPG, Z69720.1, Z92544.3 |
| omitted Genes: | STUB1, JMJD8, WDR24 |
| omitted Genes: | Z92544.2, FBXL16 |
| omitted GWAS Hits: | NA, NA |
| omitted GWAS Hits: | NA, NA |
| omitted GWAS Hits: | NA, NA |
| omitted GWAS Hits: | NA, NA |
| omitted GWAS Hits: | NA, NA |
| omitted GWAS Hits: | NA, NA |
| omitted GWAS Hits: | NA, NA |
| omitted GWAS Hits: | NA, NA |

Make more plots at http://csg.sph.umich.edu/locuszoom/

GWAS Catalog SNPs in Region

| chr | pos (Mb) | trait | snp |
|---|---|---|---|
| 16 | 0.099541 | Hemoglobin levels | rs570013781 |
| 16 | 0.113599 | Red blood cell traits | rs11248850 |
| 16 | 0.134391 | Hematology traits | rs7203560 |
| 16 | 0.134391 | Mean corpuscular volume | rs7203560 |
| 16 | 0.134391 | Mean corpuscular hemoglobin concentration | rs7203560 |
| 16 | 0.134391 | Mean corpuscular hemoglobin | rs7203560 |
| 16 | 0.134391 | Sickle cell anemia (haemolysis) | rs7203560 |
| 16 | 0.175654 | Mean corpuscular hemoglobin | rs2858942 |
| 16 | 0.190281 | Hematology traits | rs1211375 |
| 16 | 0.190281 | Mean corpuscular volume | rs1211375 |
| 16 | 0.190281 | Mean corpuscular hemoglobin | rs1211375 |
| 16 | 0.248589 | Red blood cell traits | rs13339636 |
| 16 | 0.254804 | Mean corpuscular volume | rs7189020 |
| 16 | 0.259156 | Mean corpuscular hemoglobin | rs1122794 |
| 16 | 0.259156 | Red blood cell traits | rs1122794 |
| 16 | 0.260381 | Red blood cell traits | rs13335629 |
| 16 | 0.264781 | Red blood cell traits | rs9924561 |
| 16 | 0.284580 | Morning vs. evening chronotype | rs432925 |
| 16 | 0.297063 | Bone mineral density (spine) | rs117208012 |
| 16 | 0.325782 | Bone mineral density | rs9921222 |
| 16 | 0.325782 | Bone mineral density (spine) | rs9921222 |

GWAS Catalog SNPs in Region

| chr | pos (Mb) | trait | snp |
|---|---|---|---|
| 16 | 0.337867 | Body mass index | rs11866815 |
| 16 | 0.530124 | Mean corpuscular hemoglobin concentration | rs2266928 |
| 16 | 0.580665 | Disc degeneration (lumbar) | rs1981483 |
| 16 | 0.587212 | Disc degeneration (lumbar) | rs2017567 |
| 16 | 0.611335 | Disc degeneration (lumbar) | rs7204439 |
| 16 | 0.625680 | Height | rs763014 |
