## Supplemental File 1 for "Genetic Associations with Age at Dementia Onset in the *PSEN1 E280A* Colombian Kindred": chr16_53696382-54696839.pdf

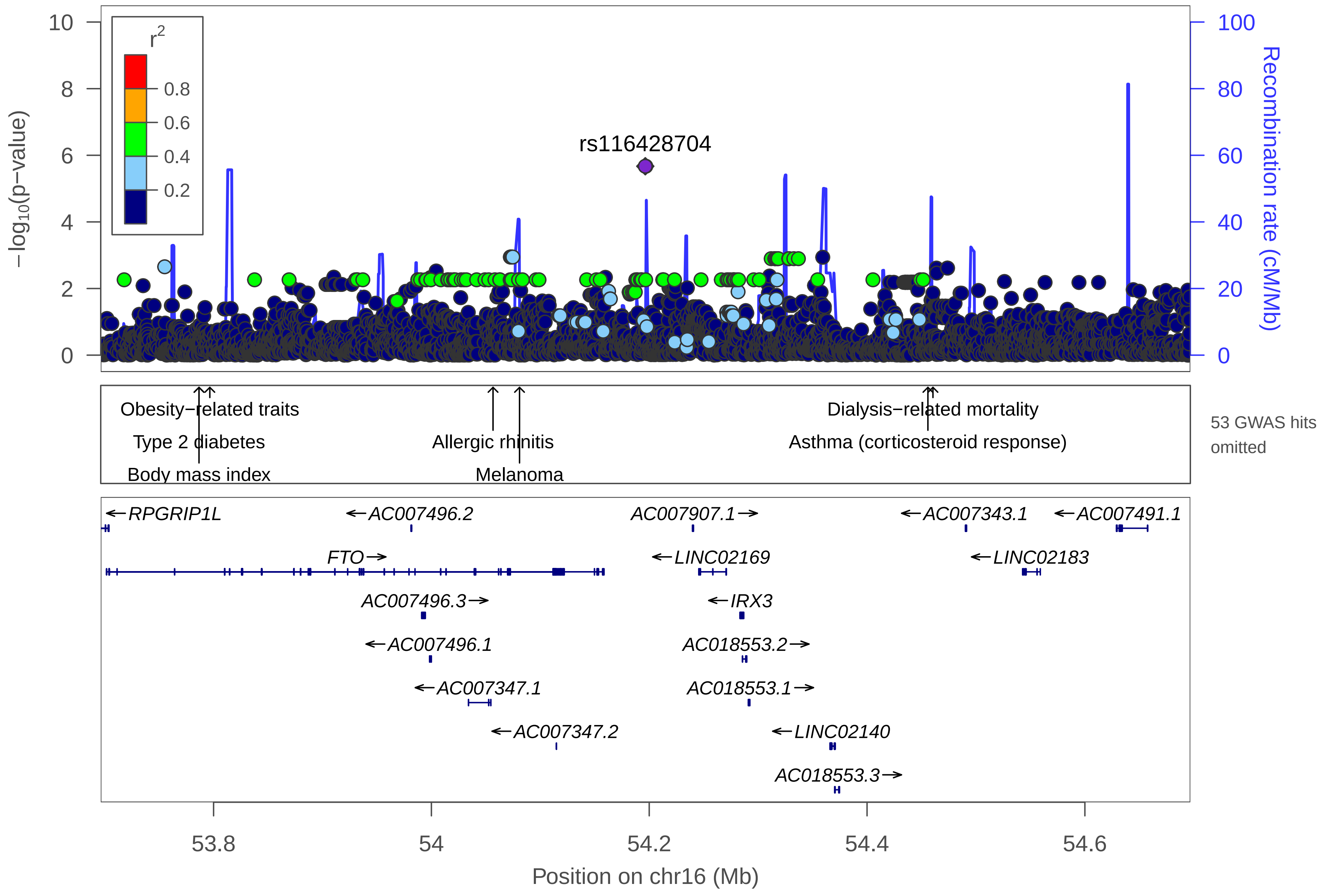

rs116428704
r2
0.8
0.6
0.4
0.2
−log10(p−value)
Recombination rate (cM/Mb)
Obesity−related traits
Type 2 diabetes
Body mass index
Allergic rhinitis
Melanoma
Dialysis−related mortality
Asthma (corticosteroid response)
53 GWAS hits omitted
RPGRIP1L
AC007496.2
AC007907.1
AC007343.1
AC007491.1
FTO
LINC02169
LINC02183
AC007496.3
IRX3
AC007496.1
AC018553.2
AC007347.1
AC018553.1
AC007347.2
LINC02140
AC018553.3
53.8
54
54.2
54.4
54.6
Position on chr16 (Mb)

| | |
|---|---|
| date: | Thu Oct 20 22:13:35 2022 |
| build: | hg38 |
| display range: | chr16:53696382−54696839 [53696382−54696839] |
| hilite range: | 0 − 0 [ 0 − 0 ] |
| reference SNP: | chr16:54196382 |
| number of SNPs plotted: | 4154 |
| min P−value: | 2.13E−6 [chr16:54196382] |
| max P−value: | 9.99E−1 [chr16:54415261] |
| omitted GWAS Hits: | chr16:54.460512−Dialysis−related mortality, chr16:54.056583−Allergic rhinitis |
| omitted GWAS Hits: | chr16:54.080912−Melanoma, NA |
| omitted GWAS Hits: | NA, NA |
| omitted GWAS Hits: | NA, NA |
| omitted GWAS Hits: | NA, NA |
| omitted GWAS Hits: | NA, NA |
| omitted GWAS Hits: | NA, NA |
| omitted GWAS Hits: | NA, NA |
| omitted GWAS Hits: | NA, NA |
| omitted GWAS Hits: | NA, NA |
| omitted GWAS Hits: | NA, NA |
| omitted GWAS Hits: | NA, NA |
| omitted GWAS Hits: | NA, NA |
| omitted GWAS Hits: | NA, NA |
| omitted GWAS Hits: | NA, NA |

Make more plots at http://csg.sph.umich.edu/locuszoom/

GWAS Catalog SNPs in Region

| chr | pos (Mb) | trait | snp |
|---|---|---|---|
| 16 | 53.72383 | Subcutaneous adipose tissue | rs1421084 |
| 16 | 53.73577 | Body mass index | rs6499640 |
| 16 | 53.73577 | Weight | rs6499640 |
| 16 | 53.76684 | Metabolic syndrome | rs9940128 |
| 16 | 53.76684 | Body mass index | rs9940128 |
| 16 | 53.76704 | Dietary macronutrient intake | rs1421085 |
| 16 | 53.76704 | Obesity | rs1421085 |
| 16 | 53.76704 | Obesity (early onset extreme) | rs1421085 |
| 16 | 53.76704 | Childhood body mass index | rs1421085 |
| 16 | 53.76704 | Type 2 diabetes | rs1421085 |
| 16 | 53.76966 | Waist circumference | rs1558902 |
| 16 | 53.76966 | Body mass index | rs1558902 |
| 16 | 53.76966 | Obesity | rs1558902 |
| 16 | 53.76966 | Height adjusted BMI | rs1558902 |
| 16 | 53.76966 | Obesity (early onset extreme) | rs1558902 |
| 16 | 53.76966 | Body mass index (age interaction) | rs1558902 |
| 16 | 53.76966 | Childhood body mass index | rs1558902 |
| 16 | 53.76966 | Body fat percentage | rs1558902 |
| 16 | 53.76966 | C−reactive protein levels or HDL−cholesterol levels (pleiotropy) | rs1558902 |
| 16 | 53.77533 | Body mass index | rs1121980 |
| 16 | 53.77533 | Obesity (early onset extreme) | rs1121980 |

GWAS Catalog SNPs in Region

| chr | pos (Mb) | trait | snp |
|---|---|---|---|
| 16 | 53.77533 | HDL cholesterol | rs1121980 |
| 16 | 53.77533 | Triglycerides | rs1121980 |
| 16 | 53.77677 | Sasang constitutional medicine type (So−Eum) | rs7193144 |
| 16 | 53.77788 | Body mass index | rs62033400 |
| 16 | 53.77945 | Obesity | rs17817449 |
| 16 | 53.77945 | Breast cancer | rs17817449 |
| 16 | 53.77945 | Breast cancer (estrogen−receptor negative) | rs17817449 |
| 16 | 53.77954 | Obesity | rs8043757 |
| 16 | 53.77954 | circulating leptin levels | rs8043757 |
| 16 | 53.78236 | Type 2 diabetes | rs8050136 |
| 16 | 53.78236 | Body mass in chronic obstructive pulmonary disease | rs8050136 |
| 16 | 53.78236 | Adiposity | rs8050136 |
| 16 | 53.78236 | Body mass index | rs8050136 |
| 16 | 53.78236 | Menarche (age at onset) | rs8050136 |
| 16 | 53.78236 | Weight | rs8050136 |
| 16 | 53.78526 | Type 2 diabetes | rs9936385 |
| 16 | 53.78598 | Body mass index | rs11075990 |
| 16 | 53.78661 | Type 2 diabetes | rs9939609 |
| 16 | 53.78661 | Body mass index | rs9939609 |
| 16 | 53.78661 | Biomedical quantitative traits | rs9939609 |
| 16 | 53.78661 | Menarche (age at onset) | rs9939609 |

GWAS Catalog SNPs in Region

| chr | pos (Mb) | trait | snp |
|---|---|---|---|
| 16 | 53.78770 | Body mass index | rs7202116 |
| 16 | 53.78874 | Obesity | rs7185735 |
| 16 | 53.79158 | Obesity (extreme) | rs9941349 |
| 16 | 53.79415 | Body mass index | rs17817964 |
| 16 | 53.79655 | Obesity−related traits | rs9930506 |
| 16 | 53.79786 | Subcutaneous adipose tissue | rs9922619 |
| 16 | 53.80522 | Osteoarthritis | rs8044769 |
| 16 | 53.80900 | Body mass index | rs12149832 |
| 16 | 53.81157 | Type 2 diabetes | rs11642841 |
| 16 | 53.82138 | Breast cancer | rs11075995 |
| 16 | 53.82138 | Breast cancer (estrogen−receptor negative) | rs11075995 |
| 16 | 53.82614 | Response to thiopurine in inflammatory bowel disease (leukopenia) | rs79206939 |
| 16 | 53.85383 | Eyebrow thickness | rs12597422 |
| 16 | 53.99406 | Sex hormone−binding globulin levels | rs12596210 |
| 16 | 54.05658 | Allergic rhinitis | rs7187423 |
| 16 | 54.08091 | Melanoma | rs16953002 |
| 16 | 54.45588 | Asthma (corticosteroid response) | rs2388639 |
| 16 | 54.46051 | Dialysis−related mortality | rs9921518 |
