## Supplemental File 1 for "Genetic Associations with Age at Dementia Onset in the *PSEN1 E280A* Colombian Kindred": chr16_79900141-80926104.pdf

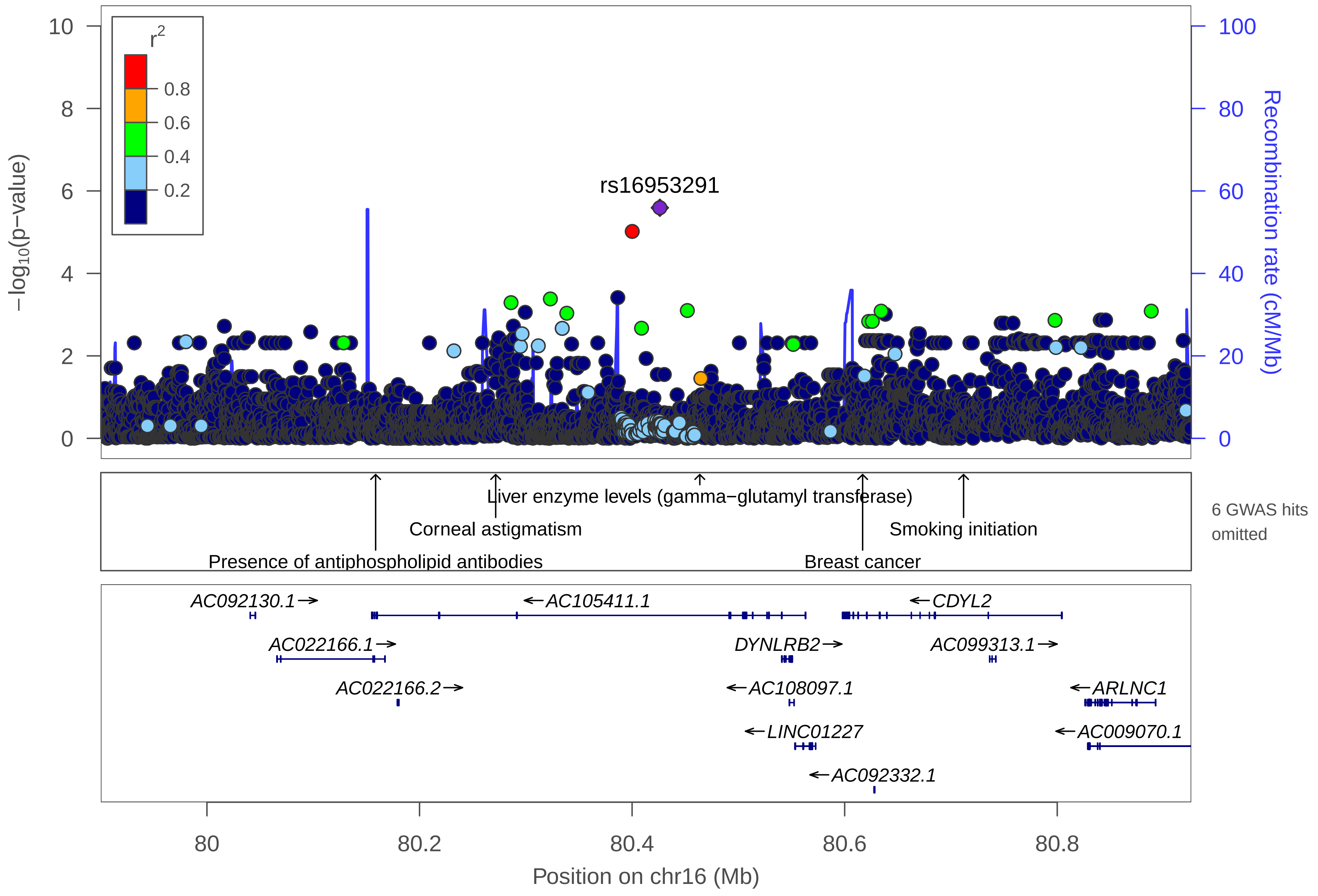

rs16953291
r2
0.8
0.6
0.4
0.2
−log10(p−value)
Recombination rate (cM/Mb)
Liver enzyme levels (gamma−glutamyl transferase)
Corneal astigmatism
Smoking initiation
Presence of antiphospholipid antibodies
Breast cancer
6 GWAS hits omitted
AC092130.1→
←AC105411.1
←CDYL2
AC022166.1→
DYNLRB2→
AC099313.1→
AC022166.2→
←AC108097.1
←ARLNC1
←LINC01227
←AC009070.1
←AC092332.1
80
80.2
80.4
80.6
80.8
Position on chr16 (Mb)

| | |
|---|---|
| date: | Thu Oct 20 22:13:36 2022 |
| build: | hg38 |
| display range: | chr16:79900141−80926104 [79900141−80926104] |
| hilite range: | 0 − 0 [ 0 − 0 ] |
| reference SNP: | chr16:80426104 |
| number of SNPs plotted: | 5085 |
| min P−value: | 2.56E−6 [chr16:80426104] |
| max P−value: | 9.99E−1 [chr16:80707691] |
| omitted GWAS Hits: | NA, NA |
| omitted GWAS Hits: | NA, NA |

Make more plots at http://csg.sph.umich.edu/locuszoom/

GWAS Catalog SNPs in Region

| chr | pos (Mb) | trait | snp |
|---|---|---|---|
| 16 | 79.95780 | Depressive symptoms (SSRI exposure interaction) | rs10514475 |
| 16 | 80.07197 | Bipolar disorder and eating disorder | rs12149074 |
| 16 | 80.07197 | Eating disorder in bipolar disorder | rs12149074 |
| 16 | 80.15871 | Presence of antiphospholipid antibodies | rs8060581 |
| 16 | 80.26465 | Alzheimer disease and age of onset | rs79358781 |
| 16 | 80.27165 | Corneal astigmatism | rs11859036 |
| 16 | 80.43330 | Alzheimer disease and age of onset | rs137967137 |
| 16 | 80.46370 | Liver enzyme levels (gamma−glutamyl transferase) | rs4581712 |
| 16 | 80.61691 | Breast cancer | rs13329835 |
| 16 | 80.71188 | Smoking initiation | rs41472047 |
| 16 | 80.75155 | Inflammatory bowel disease | rs16953946 |
