## Supplemental File 1 for "Genetic Associations with Age at Dementia Onset in the *PSEN1 E280A* Colombian Kindred": chr17_31235486-32235486.pdf

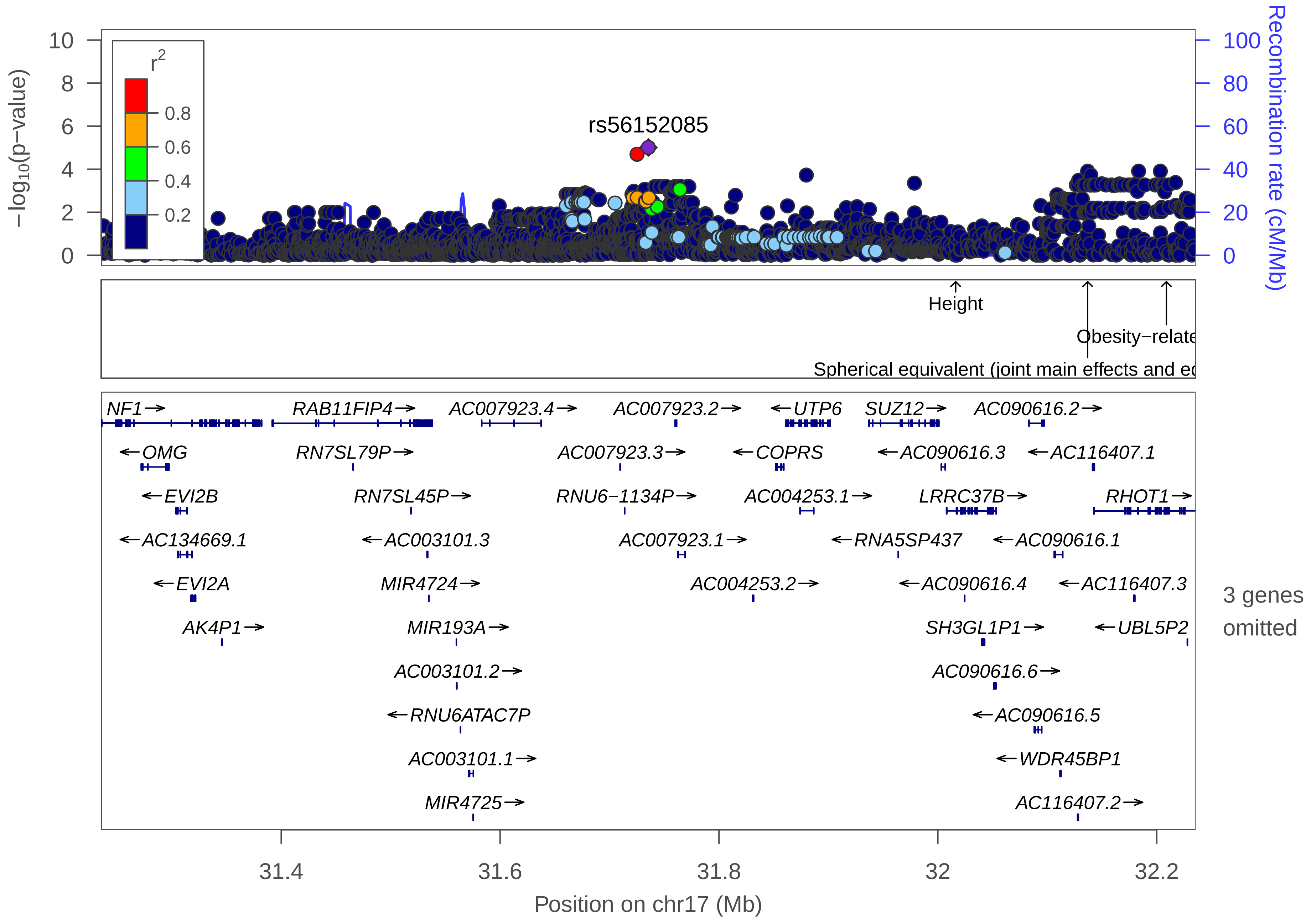
rs56152085
r2
0.8
0.6
0.4
0.2
−log10(p−value)
Recombination rate (cM/Mb)
Height
Obesity−relate
Spherical equivalent (joint main effects and ec
NF1
OMG
EVI2B
AC134669.1
EVI2A
AK4P1
RAB11FIP4
RN7SL79P
RN7SL45P
AC003101.3
MIR4724
MIR193A
AC003101.2
RNU6ATAC7P
AC003101.1
MIR4725
AC007923.4
AC007923.2
AC007923.3
RNU6−1134P
AC007923.1
AC004253.2
UTP6
COPRS
AC004253.1
RNA5SP437
SUZ12
AC090616.3
LRRC37B
AC090616.4
SH3GL1P1
AC090616.6
AC090616.5
WDR45BP1
AC090616.2
AC116407.1
RHOT1
AC090616.1
AC116407.3
UBL5P2
AC116407.2
3 genes omitted
31.4
31.6
31.8
32
32.2
Position on chr17 (Mb)

| | |
|---|---|
| date: | Thu Oct 20 22:13:35 2022 |
| build: | hg38 |
| display range: | chr17:31235486−32235486 [31235486−32235486] |
| hilite range: | 0 − 0 [ 0 − 0 ] |
| reference SNP: | chr17:31735486 |
| number of SNPs plotted: | 2994 |
| min P−value: | 9.84E−6 [chr17:31735486] |
| max P−value: | 9.98E−1 [chr17:31483072] |
| omitted Genes: | MIR365B, ARGFXP2, AC116407.4 |

Make more plots at http://csg.sph.umich.edu/locuszoom/

GWAS Catalog SNPs in Region

| chr | pos (Mb) | trait | snp |
|---|---|---|---|
| 17 | 32.01626 | Height | rs17780086 |
| 17 | 32.13687 | Spherical equivalent (joint main effects and education interaction) | rs72483203 |
| 17 | 32.20884 | Obesity−related traits | rs17780304 |
