## Supplemental File 1 for "Genetic Associations with Age at Dementia Onset in the *PSEN1 E280A* Colombian Kindred": chr18_32057937-33145059.pdf

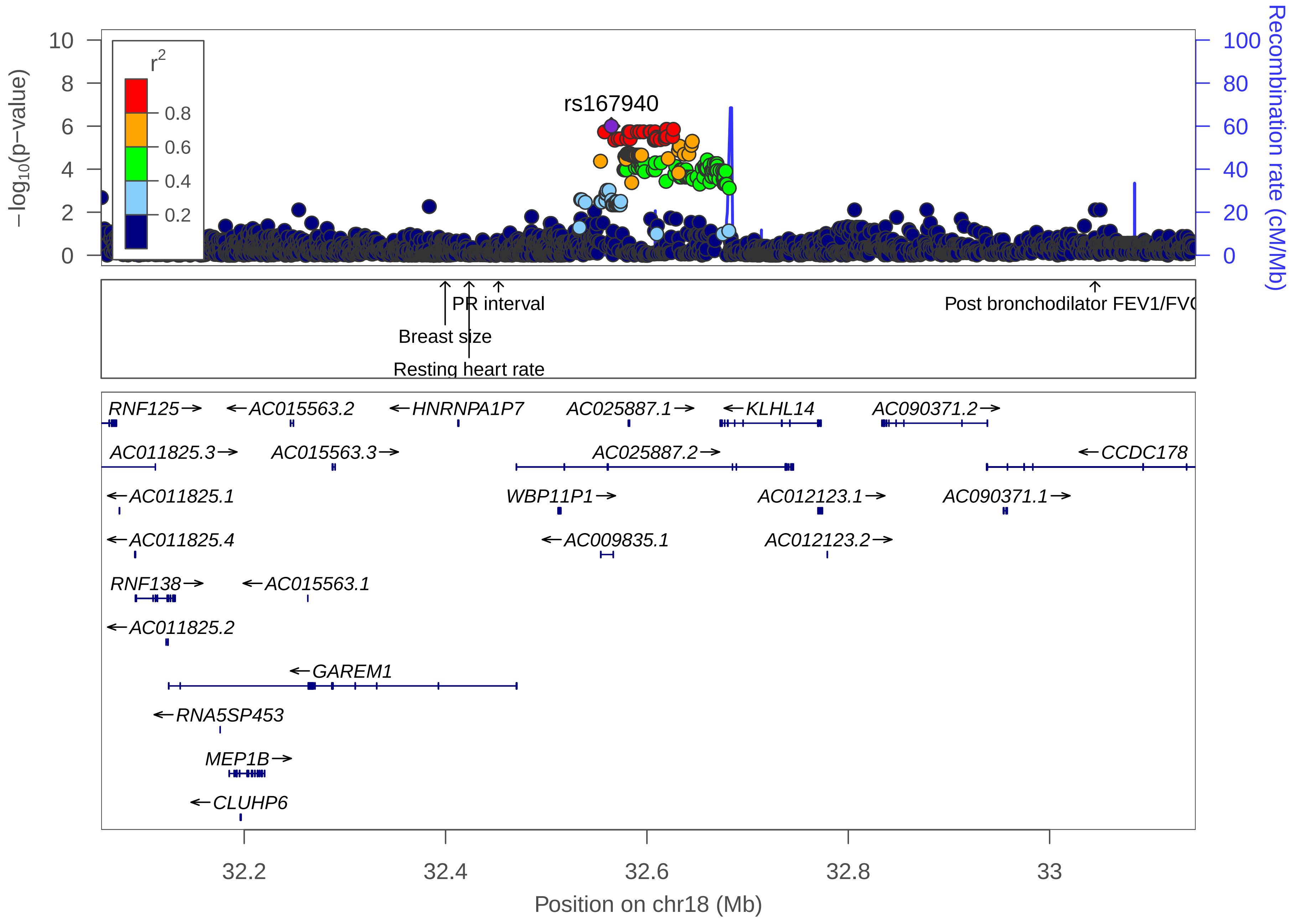

rs167940
r2
0.8
0.6
0.4
0.2
−log10(p−value)
Recombination rate (cM/Mb)
PR interval
Breast size
Resting heart rate
Post bronchodilator FEV1/FV
RNF125
AC015563.2
HNRNPA1P7
AC025887.1
KLHL14
AC090371.2
AC011825.3
AC015563.3
AC025887.2
CCDC178
AC011825.1
WBP11P1
AC012123.1
AC090371.1
AC011825.4
AC009835.1
AC012123.2
RNF138
AC015563.1
AC011825.2
GAREM1
RNA5SP453
MEP1B
CLUHP6
32.2
32.4
32.6
32.8
33
Position on chr18 (Mb)

| | |
|---|---|
| date: | Thu Oct 20 22:13:31 2022 |
| build: | hg38 |
| display range: | chr18:32057937−33145059 [32057937−33145059] |
| hilite range: | 0 − 0 [ 0 − 0 ] |
| reference SNP: | chr18:32564660 |
| number of SNPs plotted: | 2809 |
| min P−value: | 1E−6 [chr18:32564660] |
| max P−value: | 9.99E−1 [chr18:32444036] |

Make more plots at http://csg.sph.umich.edu/locuszoom/

GWAS Catalog SNPs in Region

| chr | pos (Mb) | trait | snp |
|---|---|---|---|
| 18 | 32.39961 | Breast size | rs58263042 |
| 18 | 32.42336 | Resting heart rate | rs11081761 |
| 18 | 32.45256 | PR interval | rs17744182 |
| 18 | 33.04505 | Post bronchodilator FEV1/FVC ratio | rs190650493 |
