## Supplemental File 1 for "Genetic Associations with Age at Dementia Onset in the *PSEN1 E280A* Colombian Kindred": chr18_59418230-60428012.pdf

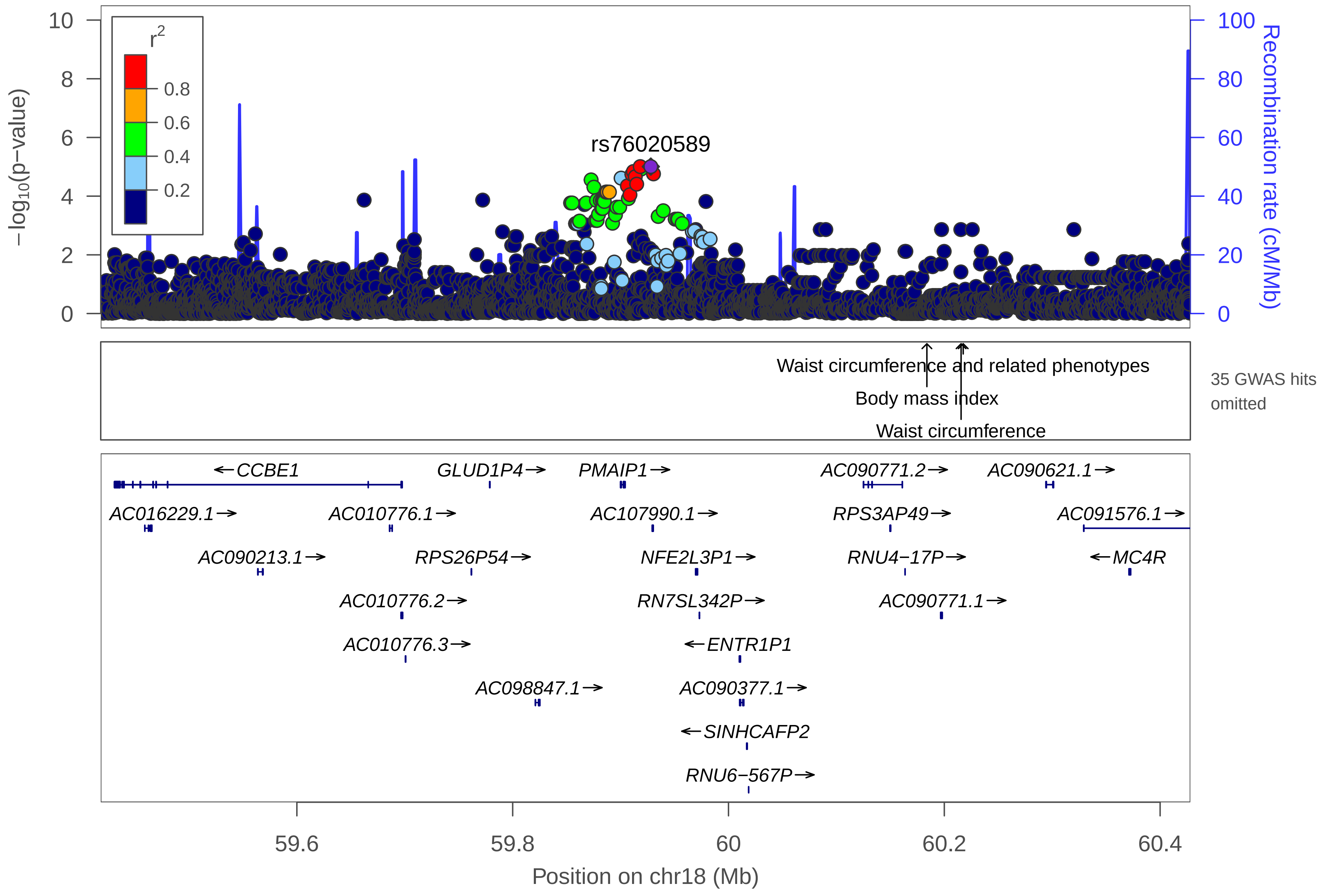

rs76020589
r2
0.8
0.6
0.4
0.2
−log10(p−value)
Recombination rate (cM/Mb)
Waist circumference and related phenotypes
Body mass index
Waist circumference
35 GWAS hits omitted
←CCBE1
GLUD1P4→
PMAIP1→
AC090771.2→
AC090621.1→
AC016229.1→
AC010776.1→
AC107990.1→
RPS3AP49→
AC091576.1→
AC090213.1→
RPS26P54→
NFE2L3P1→
RNU4−17P→
←MC4R
AC010776.2→
RN7SL342P→
AC090771.1→
AC010776.3→
←ENTR1P1
AC098847.1→
AC090377.1→
←SINHCAFP2
RNU6−567P→
59.6
59.8
60
60.2
60.4
Position on chr18 (Mb)

| | |
|---|---|
| date: | Thu Oct 20 22:13:34 2022 |
| build: | hg38 |
| display range: | chr18:59418230−60428012 [59418230−60428012] |
| hilite range: | 0 − 0 [ 0 − 0 ] |
| reference SNP: | chr18:59928012 |
| number of SNPs plotted: | 4182 |
| min P−value: | 9.85E−6 [chr18:59918230] |
| max P−value: | 9.99E−1 [chr18:59721823] |
| omitted GWAS Hits: | NA, NA |
| omitted GWAS Hits: | NA, NA |
| omitted GWAS Hits: | NA, NA |
| omitted GWAS Hits: | NA, NA |
| omitted GWAS Hits: | NA, NA |
| omitted GWAS Hits: | NA, NA |
| omitted GWAS Hits: | NA, NA |
| omitted GWAS Hits: | NA, NA |
| omitted GWAS Hits: | NA, NA |
| omitted GWAS Hits: | NA, NA |
| omitted GWAS Hits: | NA, NA |
| omitted GWAS Hits: | NA, NA |

Make more plots at http://csg.sph.umich.edu/locuszoom/

GWAS Catalog SNPs in Region

| chr | pos (Mb) | trait | snp |
|---|---|---|---|
| 18 | 59.48481 | Age−related hearing impairment | rs12955474 |
| 18 | 59.64758 | Major depressive disorder | rs11152166 |
| 18 | 59.83615 | Response to bleomycin (chromatid breaks) | rs8093763 |
| 18 | 59.91425 | Alzheimer disease and age of onset | rs142176337 |
| 18 | 59.95506 | Chronic lymphocytic leukemia | rs4368253 |
| 18 | 60.00657 | Body mass index | rs12964056 |
| 18 | 60.06762 | Body mass index | rs7234864 |
| 18 | 60.08378 | Metabolic syndrome | rs12957347 |
| 18 | 60.14475 | Obesity (early onset extreme) | rs17700144 |
| 18 | 60.16140 | Body mass index | rs2331841 |
| 18 | 60.16190 | Body mass index | rs6567160 |
| 18 | 60.16190 | Fat body mass | rs6567160 |
| 18 | 60.16190 | Childhood body mass index | rs6567160 |
| 18 | 60.16190 | Body fat percentage | rs6567160 |
| 18 | 60.17117 | Coronary artery disease | rs663129 |
| 18 | 60.17254 | Body mass index | rs571312 |
| 18 | 60.17254 | Body mass index (age interaction) | rs571312 |
| 18 | 60.17254 | C−reactive protein levels or triglyceride levels (pleiotropy) | rs571312 |
| 18 | 60.17436 | Body mass index | rs591166 |
| 18 | 60.18179 | HDL cholesterol | rs12967135 |
| 18 | 60.18179 | C−reactive protein levels or HDL−cholesterol levels (pleiotropy) | rs12967135 |

### GWAS Catalog SNPs in Region

| chr | pos (Mb) | trait | snp |
|---|---|---|---|
| 18 | 60.18319 | Obesity | rs538656 |
| 18 | 60.18386 | Body mass index | rs17782313 |
| 18 | 60.18386 | Height | rs17782313 |
| 18 | 60.18386 | Obesity | rs17782313 |
| 18 | 60.18453 | Obesity | rs10871777 |
| 18 | 60.18535 | Obesity (early onset extreme) | rs476828 |
| 18 | 60.18572 | Obesity | rs11152213 |
| 18 | 60.18572 | Height | rs11152213 |
| 18 | 60.19160 | Body mass index | rs8089364 |
| 18 | 60.20576 | Urate levels | rs12955983 |
| 18 | 60.21555 | Waist circumference | rs489693 |
| 18 | 60.21555 | Antipsychotic drug−induced weight gain | rs489693 |
| 18 | 60.21752 | Waist circumference and related phenotypes | rs12970134 |
| 18 | 60.21752 | Body mass index | rs12970134 |
| 18 | 60.21752 | Type 2 diabetes | rs12970134 |
| 18 | 60.21752 | Weight | rs12970134 |
| 18 | 60.29588 | Obesity−related traits | rs17773430 |
