## Supplemental File 1 for "Genetic Associations with Age at Dementia Onset in the *PSEN1 E280A* Colombian Kindred": chr19_1796860-2796863.pdf

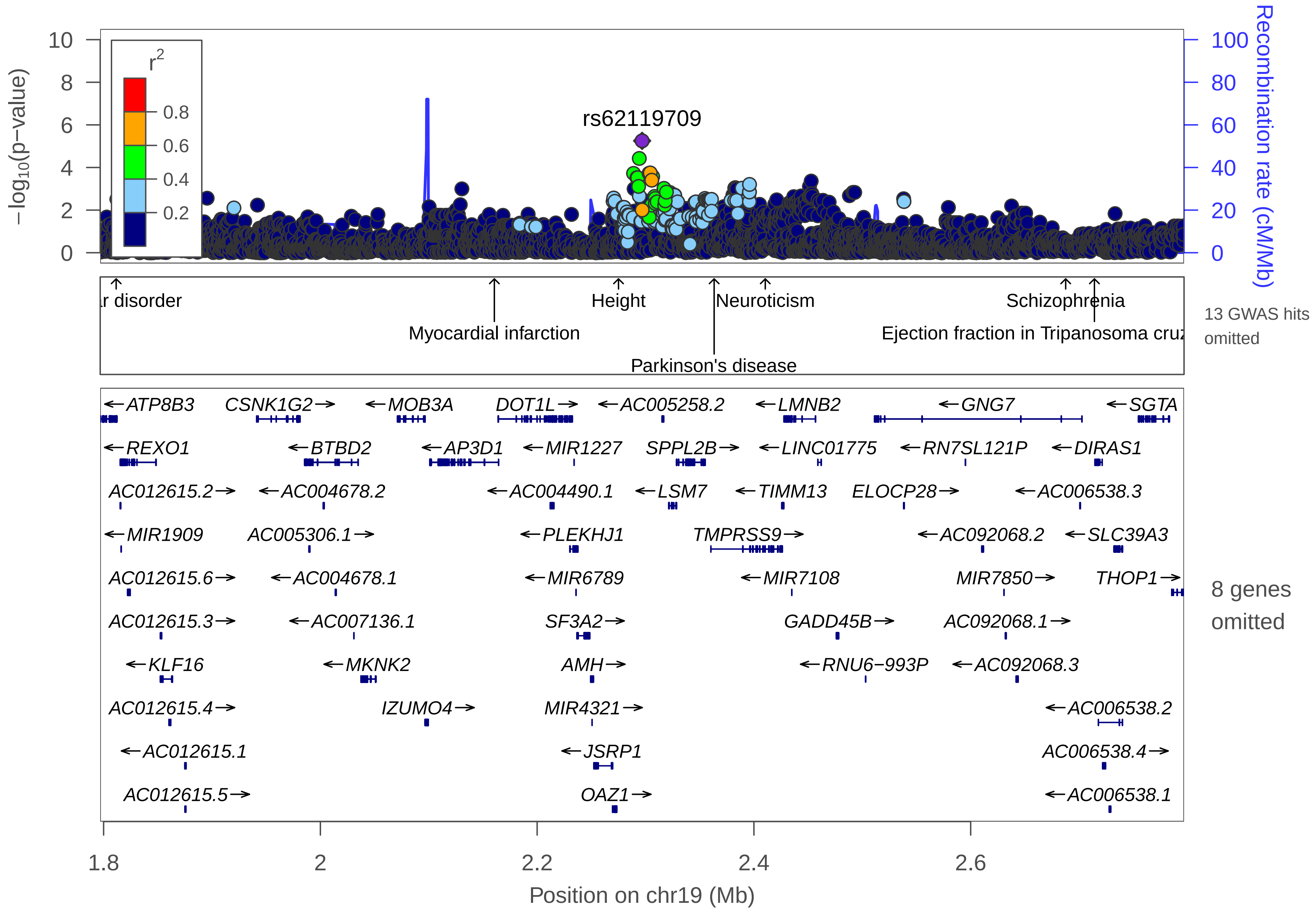

rs62119709
−log10(p−value)
Recombination rate (cM/Mb)
r2
0.8
0.6
0.4
0.2
r disorder
Myocardial infarction
Height
Parkinson's disease
Neuroticism
Schizophrenia
Ejection fraction in Tripanosoma cruz
13 GWAS hits omitted
ATP8B3
CSNK1G2
MOB3A
DOT1L
AC005258.2
LMNB2
GNG7
SGTA
REXO1
BTBD2
AP3D1
MIR1227
SPPL2B
LINC01775
RN7SL121P
DIRAS1
AC012615.2
AC004678.2
AC004490.1
LSM7
TIMM13
ELOCP28
AC006538.3
MIR1909
AC005306.1
PLEKHJ1
TMPRSS9
AC092068.2
SLC39A3
AC012615.6
AC004678.1
MIR6789
MIR7108
MIR7850
THOP1
AC012615.3
AC007136.1
SF3A2
GADD45B
AC092068.1
KLF16
MKNK2
AMH
RNU6−993P
AC092068.3
AC012615.4
IZUMO4
MIR4321
AC006538.2
AC012615.1
JSRP1
AC006538.4
AC012615.5
OAZ1
AC006538.1
8 genes omitted
1.8
2
2.2
2.4
2.6
Position on chr19 (Mb)

| | |
|---|---|
| date: | Thu Oct 20 22:13:36 2022 |
| build: | hg38 |
| display range: | chr19:1796860−2796863 [1796860−2796863] |
| hilite range: | 0 − 0 [ 0 − 0 ] |
| reference SNP: | chr19:2296863 |
| number of SNPs plotted: | 3977 |
| min P−value: | 5.61E−6 [chr19:2296860] |
| max P−value: | 10E−1 [chr19:1833291] |
| omitted Genes: | ABHD17A, SCAMP4, ADAT3 |
| omitted Genes: | CSNK1G2−AS1, AC005258.1, PEAK3 |
| omitted Genes: | LINGO3, AC104530.1 |
| omitted GWAS Hits: | chr19:2.714172−Ejection fraction in Tripanosoma cruzi seropositivity, chr19:2.687653−Schizoph |
| omitted GWAS Hits: | NA, NA |
| omitted GWAS Hits: | NA, NA |
| omitted GWAS Hits: | NA, NA |
| omitted GWAS Hits: | NA |

Make more plots at http://csg.sph.umich.edu/locuszoom/

GWAS Catalog SNPs in Region

| chr | pos (Mb) | trait | snp |
|---|---|---|---|
| 19 | 1.803583 | Obesity−related traits | rs7253430 |
| 19 | 1.811604 | Bipolar disorder | rs7250872 |
| 19 | 1.937194 | Body mass index | rs11672550 |
| 19 | 2.048282 | Subjective well−being | rs12610177 |
| 19 | 2.160530 | Myocardial infarction | rs3803915 |
| 19 | 2.160530 | Body mass index | rs3803915 |
| 19 | 2.170955 | Height | rs12986413 |
| 19 | 2.176404 | Height | rs11880992 |
| 19 | 2.176404 | Hip minimal joint space width | rs11880992 |
| 19 | 2.177194 | Osteoarthritis | rs12982744 |
| 19 | 2.177194 | Height | rs12982744 |
| 19 | 2.214058 | Plasma omega−6 polyunsaturated fatty acid levels (gamma−linolenic acid) | rs2074552 |
| 19 | 2.232222 | Pulse pressure | rs740406 |
| 19 | 2.275080 | Height | rs2523178 |
| 19 | 2.363321 | Parkinson's disease | rs62120679 |
| 19 | 2.410486 | Neuroticism | rs4806846 |
| 19 | 2.522673 | 3−hydroxypropylmercapturic acid levels in smokers | rs74407240 |
| 19 | 2.547017 | Ejection fraction in Tripanosoma cruzi seropositivity | rs6510683 |
| 19 | 2.687653 | Schizophrenia | rs57549656 |
| 19 | 2.714172 | Ejection fraction in Tripanosoma cruzi seropositivity | rs185543003 |
