## Supplemental File 1 for "Genetic Associations with Age at Dementia Onset in the *PSEN1 E280A* Colombian Kindred": chr21_21556609-27678240.pdf

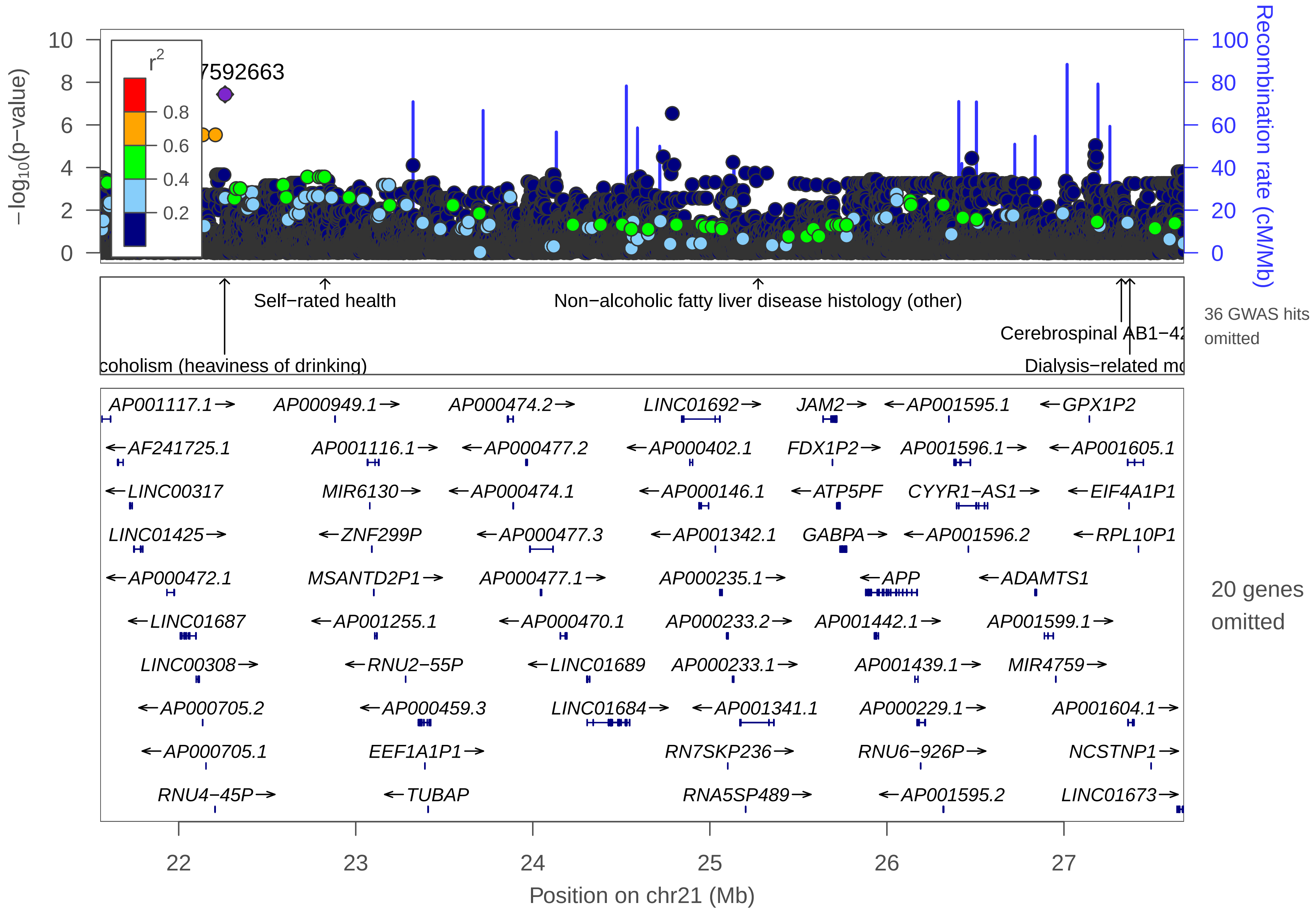

−log10(p−value)
Recombination rate (cM/Mb)
r2
0.8
0.6
0.4
0.2
7592663
Self−rated health
Non−alcoholic fatty liver disease histology (other)
Cerebrospinal AB1−42
coholism (heaviness of drinking)
Dialysis−related mo
36 GWAS hits omitted
AP001117.1→
AP000949.1→
AP000474.2→
LINC01692→
JAM2→
←AP001595.1
←GPX1P2
←AF241725.1
AP001116.1→
←AP000477.2
←AP000402.1
FDX1P2→
AP001596.1→
←AP001605.1
←LINC00317
MIR6130→
←AP000474.1
←AP000146.1
←ATP5PF
CYYR1−AS1→
←EIF4A1P1
LINC01425→
←ZNF299P
←AP000477.3
←AP001342.1
GABPA→
←AP001596.2
←RPL10P1
←AP000472.1
MSANTD2P1→
AP000477.1→
AP000235.1→
←APP
←ADAMTS1
←LINC01687
←AP001255.1
←AP000470.1
AP000233.2→
AP001442.1→
AP001599.1→
LINC00308→
←RNU2−55P
←LINC01689
AP000233.1→
AP001439.1→
MIR4759→
←AP000705.2
←AP000459.3
LINC01684→
←AP001341.1
AP000229.1→
AP001604.1→
←AP000705.1
EEF1A1P1→
RN7SKP236→
RNU6−926P→
NCSTNP1→
RNU4−45P→
←TUBAP
RNA5SP489→
←AP001595.2
LINC01673→
20 genes omitted
Position on chr21 (Mb)

| | |
|---|---|
| date: | Thu Oct 20 22:13:41 2022 |
| build: | hg38 |
| display range: | chr21:21556609−27678240 [21556609−27678240] |
| hilite range: | 0 − 0 [ 0 − 0 ] |
| reference SNP: | chr21:22261900 |
| number of SNPs plotted: | 27061 |
| min P−value: | 3.69E−8 [chr21:22261900] |
| max P−value: | 10E−1 [chr21:21980712] |
| omitted Genes: | AP000561.1, MAPK6P2, AP000959.1 |
| omitted Genes: | AP000459.1, AP000459.2, AP000961.1 |
| omitted Genes: | RPL13AP7, AP001340.1, LINC00158 |
| omitted Genes: | AP000221.1, MIR155HG, MIR155 |
| omitted Genes: | AP000223.1, MRPL39, RNGTTP1 |
| omitted Genes: | AP000224.1, LLPHP2, RNU6−123P |
| omitted Genes: | CYYR1, ADAMTS5 |
| omitted GWAS Hits: | NA, NA |
| omitted GWAS Hits: | NA, NA |
| omitted GWAS Hits: | NA, NA |
| omitted GWAS Hits: | NA, NA |
| omitted GWAS Hits: | NA, NA |
| omitted GWAS Hits: | NA, NA |
| omitted GWAS Hits: | NA, NA |
| omitted GWAS Hits: | NA, NA |

Make more plots at http://csg.sph.umich.edu/locuszoom/

GWAS Catalog SNPs in Region

| chr | pos (Mb) | trait | snp |
|---|---|---|---|
| 21 | 21.66078 | Parental longevity (combined parental age at death) | rs11088872 |
| 21 | 21.76953 | Dysphagia | rs2226441 |
| 21 | 21.77228 | Dysphagia | rs2827025 |
| 21 | 21.77363 | Diisocyanate−induced asthma | rs2014791 |
| 21 | 21.77912 | Asthma | rs7281353 |
| 21 | 21.77912 | Asthma (childhood onset) | rs7281353 |
| 21 | 22.25936 | Alcoholism (heaviness of drinking) | rs2827312 |
| 21 | 22.82649 | Self−rated health | rs7279441 |
| 21 | 23.04818 | Colorectal or endometrial cancer | rs4816890 |
| 21 | 23.73289 | Schizophrenia | rs2828478 |
| 21 | 23.77080 | Major depressive disorder | rs2828520 |
| 21 | 24.06519 | Urinary albumin−to−creatinine ratio | rs2828785 |
| 21 | 24.54093 | Alzheimer's disease in APOE e4+ carriers | rs721146 |
| 21 | 25.27277 | Non−alcoholic fatty liver disease histology (other) | rs9977253 |
| 21 | 25.45420 | Adverse response to chemotherapy (neutropenia/leucopenia) (all antimetabolite drugs) | rs8127977 |
| 21 | 25.45746 | Parasitemia in Tripanosoma cruzi seropositivity | rs12483240 |
| 21 | 25.63006 | Cognitive performance | rs17001239 |
| 21 | 25.69203 | Longitudinal change in brain amyloid plaque burden | rs8129913 |
| 21 | 26.34434 | Airflow obstruction | rs9975851 |
| 21 | 26.49984 | Response to exercise (triglyceride levels) | rs222158 |
| 21 | 26.76949 | Parental extreme longevity (95 years and older) | rs73185805 |

GWAS Catalog SNPs in Region

| chr | pos (Mb) | trait | snp |
|---|---|---|---|
| 21 | 26.77387 | Metabolite levels (HVA) | rs2830487 |
| 21 | 26.77387 | Metabolite levels (HVA−5−HIAA Factor score) | rs2830487 |
| 21 | 27.28563 | Liver enzyme levels (aspartate transaminase) | rs457603 |
| 21 | 27.29303 | Response to citalopram treatment | rs2830840 |
| 21 | 27.32416 | Cerebrospinal AB1−42 levels | rs239713 |
| 21 | 27.36268 | Dialysis−related mortality | rs9977499 |
| 21 | 27.37204 | Dialysis−related mortality | rs1452093 |
| 21 | 27.45794 | Post bronchodilator FEV1/FVC ratio | rs147130662 |
| 21 | 27.46976 | Post bronchodilator FEV1/FVC ratio | rs139916902 |
| 21 | 27.48792 | Post bronchodilator FEV1/FVC ratio | rs142181472 |
| 21 | 27.51337 | Post bronchodilator FEV1 | rs142429672 |
| 21 | 27.51337 | Post bronchodilator FEV1/FVC ratio | rs142429672 |
| 21 | 27.51781 | Daytime sleep phenotypes | rs242364 |
| 21 | 27.51811 | Post bronchodilator FEV1 | rs141838403 |
| 21 | 27.51811 | Post bronchodilator FEV1/FVC ratio | rs141838403 |
| 21 | 27.54562 | Post bronchodilator FEV1 | rs139202427 |
| 21 | 27.54562 | Post bronchodilator FEV1/FVC ratio | rs139202427 |
| 21 | 27.55480 | Post bronchodilator FEV1 | rs142976294 |
| 21 | 27.55480 | Post bronchodilator FEV1/FVC ratio | rs142976294 |
| 21 | 27.57396 | Response to cytidine analogues (gemcitabine) | rs1598848 |
