## Supplemental File 1 for "Genetic Associations with Age at Dementia Onset in the *PSEN1 E280A* Colombian Kindred": chr21_43792407-44792407.pdf

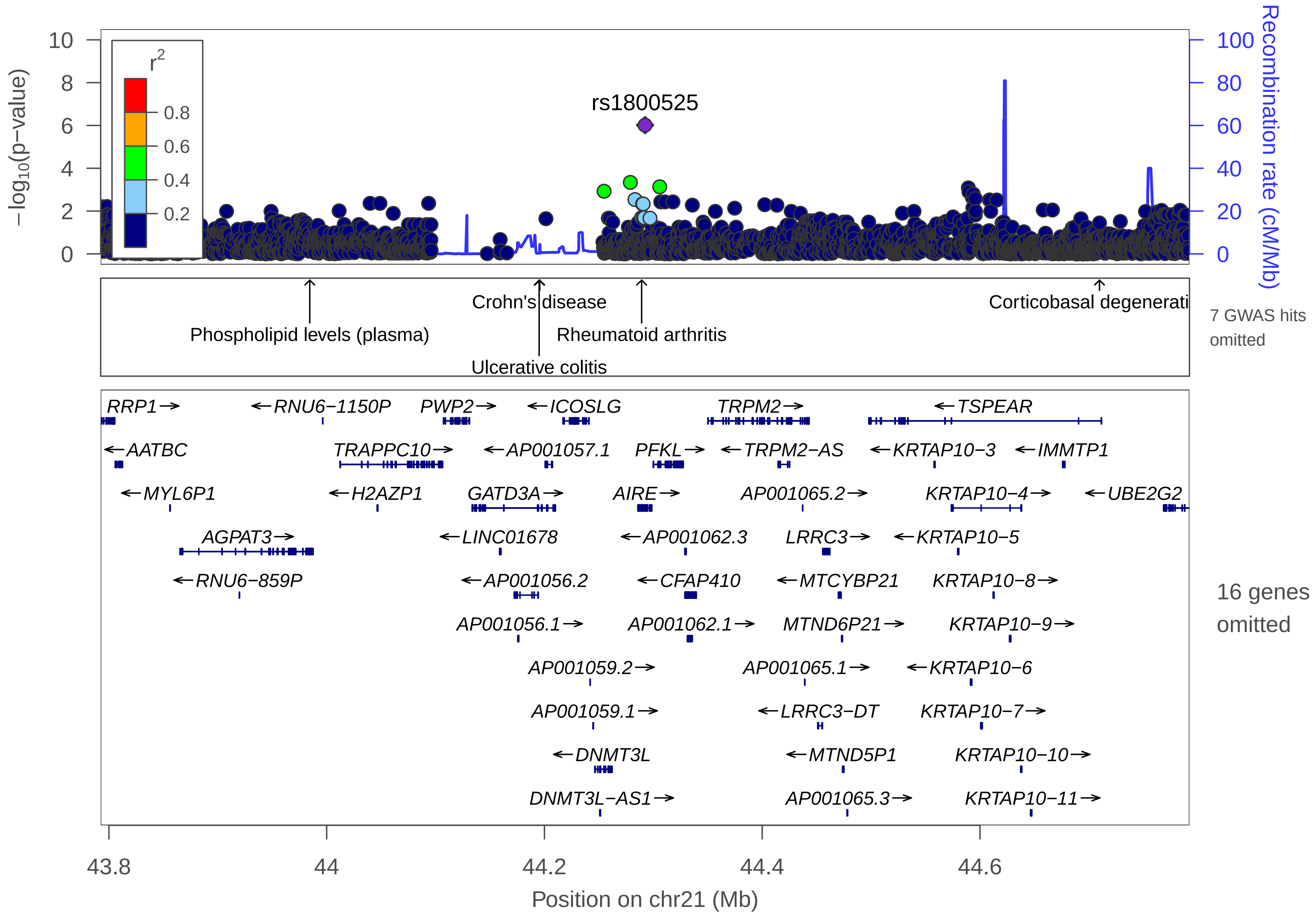

rs1800525
r2
0.8
0.6
0.4
0.2
−log10(p−value)
Recombination rate (cM/Mb)
Phospholipid levels (plasma)
Crohn's disease
Ulcerative colitis
Rheumatoid arthritis
Corticobasal degenerati
7 GWAS hits omitted
RRP1 RNU6−1150P PWP2 ICOSLG TRPM2 TSPEAR
AATBC TRAPPC10 AP001057.1 PFKL TRPM2−AS KRTAP10−3 IMMTP1
MYL6P1 H2AZP1 GATD3A AIRE AP001065.2 KRTAP10−4 UBE2G2
AGPAT3 LINC01678 AP001062.3 LRRC3 KRTAP10−5
RNU6−859P AP001056.2 CFAP410 MTCYBP21 KRTAP10−8
AP001056.1 AP001062.1 MTND6P21 KRTAP10−9
AP001059.2 AP001065.1 KRTAP10−6
AP001059.1 LRRC3−DT KRTAP10−7
DNMT3L MTND5P1 KRTAP10−10
DNMT3L−AS1 AP001065.3 KRTAP10−11
16 genes omitted
43.8 44 44.2 44.4 44.6
Position on chr21 (Mb)

| | |
|---|---|
| date: | Thu Oct 20 22:13:35 2022 |
| build: | hg38 |
| display range: | chr21:43792407−44792407 [43792407−44792407] |
| hilite range: | 0 − 0 [ 0 − 0 ] |
| reference SNP: | chr21:44292407 |
| number of SNPs plotted: | 3120 |
| min P−value: | 9.68E−7 [chr21:44292407] |
| max P−value: | 10E−1 [chr21:44689277] |
| omitted Genes: | AP001062.2, AP001063.1, LINC02575 |
| omitted Genes: | AP001065.4, TSPEAR−AS1, TSPEAR−AS2 |
| omitted Genes: | AP001066.1, KRTAP10−1, KRTAP10−2 |
| omitted Genes: | KRTAP12−4, KRTAP12−3, KRTAP12−2 |
| omitted Genes: | KRTAP12−1, KRTAP12−6P, KRTAP10−12 |
| omitted Genes: | KRTAP10−13P |
| omitted GWAS Hits: | chr21:44.709771−Corticobasal degeneration, NA |
| omitted GWAS Hits: | NA, NA |
| omitted GWAS Hits: | NA |

Make more plots at http://csg.sph.umich.edu/locuszoom/

GWAS Catalog SNPs in Region

| chr | pos (Mb) | trait | snp |
|---|---|---|---|
| 21 | 43.98446 | Phospholipid levels (plasma) | rs7435 |
| 21 | 44.19498 | Inflammatory bowel disease | rs8127691 |
| 21 | 44.19514 | Ulcerative colitis | rs2838519 |
| 21 | 44.19514 | Crohn's disease | rs2838519 |
| 21 | 44.19568 | Crohn's disease | rs762421 |
| 21 | 44.19586 | Ulcerative colitis | rs7282490 |
| 21 | 44.19586 | Crohn's disease | rs7282490 |
| 21 | 44.19586 | Inflammatory bowel disease | rs7282490 |
| 21 | 44.22480 | Left ventricular QRS voltage | rs8131653 |
| 21 | 44.22754 | Celiac disease | rs4819388 |
| 21 | 44.28927 | Rheumatoid arthritis | rs2075876 |
| 21 | 44.70977 | Corticobasal degeneration | rs875125 |
