## Supplemental File 1 for "Genetic Associations with Age at Dementia Onset in the *PSEN1 E280A* Colombian Kindred": chr22_47987809-48987809.pdf

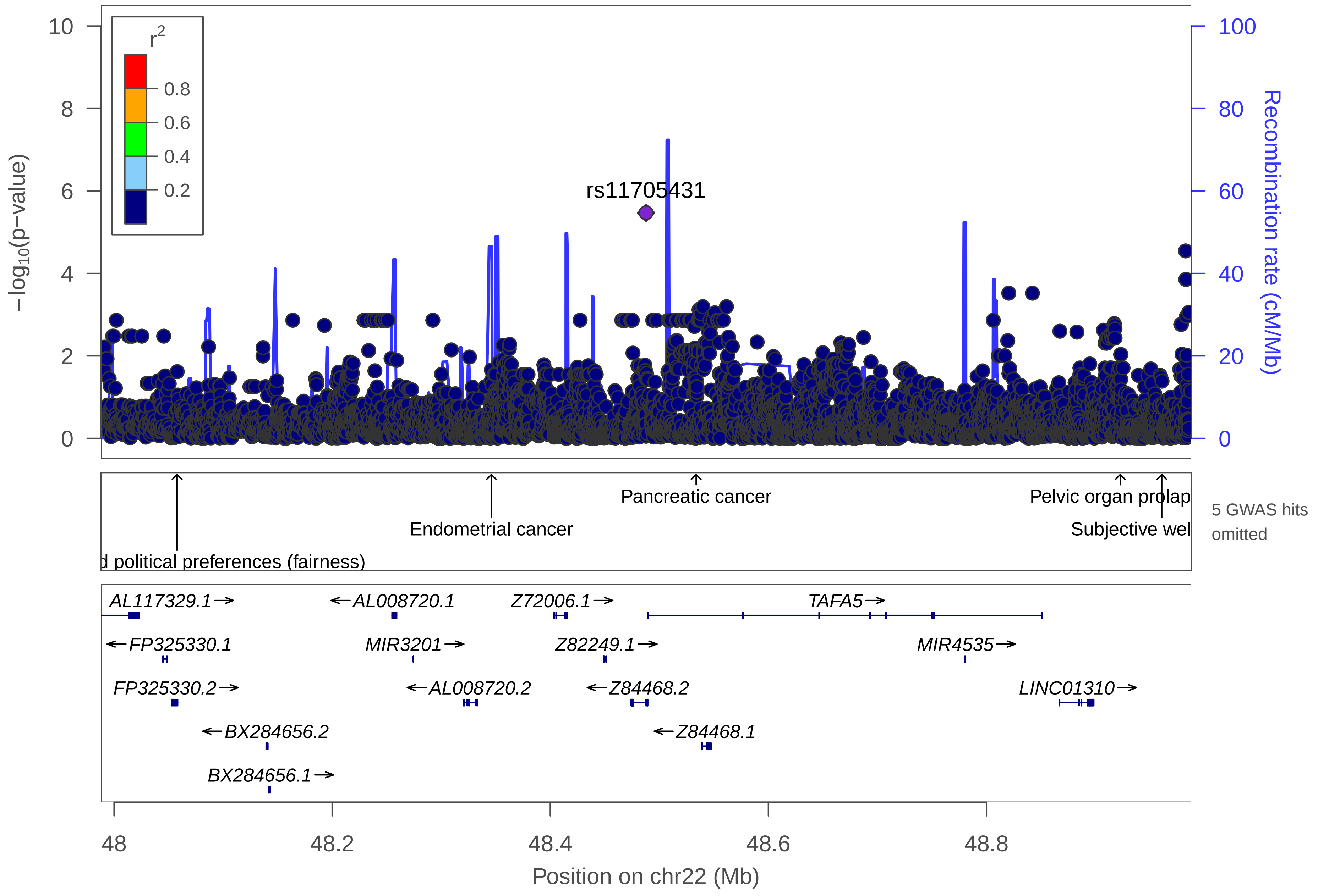

r2
0.8
0.6
0.4
0.2
−log10(p−value)
Recombination rate (cM/Mb)
rs11705431
Pancreatic cancer
Pelvic organ prolap
Endometrial cancer
Subjective wel
d political preferences (fairness)
5 GWAS hits omitted
AL117329.1→
←AL008720.1
Z72006.1→
TAFA5→
←FP325330.1
MIR3201→
Z82249.1→
MIR4535→
FP325330.2→
←AL008720.2
←Z84468.2
LINC01310→
←BX284656.2
←Z84468.1
BX284656.1→
48
48.2
48.4
48.6
48.8
Position on chr22 (Mb)

| | |
|---|---|
| date: | Thu Oct 20 22:13:36 2022 |
| build: | hg38 |
| display range: | chr22:47987809−48987809 [47987809−48987809] |
| hilite range: | 0 − 0 [ 0 − 0 ] |
| reference SNP: | chr22:48487809 |
| number of SNPs plotted: | 5946 |
| min P−value: | 3.38E−6 [chr22:48487809] |
| max P−value: | 10E−1 [chr22:48705385] |
| omitted GWAS Hits: | chr22:48.922714−Pelvic organ prolapse, chr22:48.960751−Subjective well−being |
| omitted GWAS Hits: | NA, NA |

Make more plots at http://csg.sph.umich.edu/locuszoom/

GWAS Catalog SNPs in Region

| chr | pos (Mb) | trait | snp |
|---|---|---|---|
| 22 | 48.05777 | Economic and political preferences (fairness) | rs117294 |
| 22 | 48.09884 | Alzheimer disease and age of onset | rs150927461 |
| 22 | 48.23437 | Obesity−related traits | rs13054085 |
| 22 | 48.25619 | DNA methylation (variation) | rs1004689 |
| 22 | 48.34599 | Endometrial cancer | rs4407 |
| 22 | 48.49244 | Late−onset Alzheimer's disease | rs1034435 |
| 22 | 48.52765 | Bipolar disorder (body mass index interaction) | rs80088139 |
| 22 | 48.53376 | Pancreatic cancer | rs5768709 |
| 22 | 48.92271 | Pelvic organ prolapse | rs7290192 |
| 22 | 48.96075 | Subjective well−being | rs113959751 |
